# Living alone and mental health: parallel analyses in longitudinal population surveys and electronic health records prior to and during the COVID-19 pandemic

**DOI:** 10.1101/2023.03.15.23287292

**Authors:** Eoin McElroy, Emily Herrett, Kishan Patel, Dominik M Piehlmaier, Giorgio Di Gessa, Charlotte Huggins, Michael J Green, Alex Kwong, Ellen J. Thompson, Jingmin Zhu, Kathryn E Mansfield, Richard J Silverwood, Rosie Mansfield, Jane Maddock, Rohini Mathur, Ruth E Costello, Anthony Matthews, John Tazare, Alasdair Henderson, Kevin Wing, Lucy Bridges, Sebastian Bacon, Amir Mehrkar, OpenSafely Collaborative, Richard J Shaw, Jacques Wels, Srinivasa Vittal Katikireddi, Nish Chaturvedi, Laurie Tomlinson, Praveetha Patalay, the Longitudinal Health and Wellbeing Collaborative

## Abstract

**Objectives:** To describe the mental health gap between those who live alone and those who live with others, and to examine whether the COVID-19 pandemic had an impact on this gap.

**Design:** Ten population based prospective cohort studies, and a retrospective descriptive cohort study based on electronic health records (EHRs).

**Setting:** UK Longitudinal population-based surveys (LPS), and primary and secondary care records within the OpenSAFELY-TPP database.

**Participants:** Participants from the LPS were included if they had information on living status in early 2020, valid data on mental ill-health at the closest pre-pandemic assessment and at least once during the pandemic, and valid data on a key minimum set of covariates. The EHR dataset included 16 million adults registered with primary care practices in England using TPP SystmOne software on 1st February 2020, with at least three months of registration, valid address data, and living in households of <16 people.

**Main outcome measures:** In the LPS, self-reported survey measures of psychological distress and life satisfaction were assessed in the nearest pre-pandemic sweep and three periods during the pandemic: April-June 2020, July-October 2020, and November 2020-March 2021. In the EHR analyses, outcomes were morbidity codes recorded in primary or secondary care between March 2018 and January 2022 reflecting the diagnoses of depression, self-harm, anxiety, obsessive compulsive disorder, eating disorders, and severe mental illnesses.

**Results:** The LPS consisted of 37,544 participants (15.2% living alone) and we found greater psychological distress (SMD: 0.09 (95% CI: 0.04, 0.14) and lower life satisfaction (SMD: -0.22 (95% CI: -0.30, -0.15) in those living alone pre-pandemic, and the gap between the two groups stayed similar after the onset of the pandemic. In the EHR analysis of almost 16 million records (21.4% living alone), codes indicating mental health conditions were more common in those who lived alone compared to those who lived with others (e.g., depression 26 and severe mental illness 58 cases more per 100,000). Recording of mental health conditions fell during the pandemic for common mental health disorders and the gap between the two groups narrowed.

**Conclusions:** Multiple sources of data indicate that those who live alone experience greater levels of common and severe mental illnesses, and lower life satisfaction. During the pandemic this gap in need remained, however, there was a narrowing of the gap in service use, suggesting greater barriers to healthcare access for those who live alone.

**Summary Box:** *What is already known on the topic?:* Households with one individual are an increasing demographic, comprising over a quarter of all households in the UK in 2021. However, the mental health gap between those who live alone compared to those who live with others is not well described and even less is known about the relative gaps in need and healthcare-seeking and access. The pandemic and associated restrictive measures further increased the likelihood of isolation for this group, which may have impacted mental health.

*What this study adds?:* We present comprehensive evidence from both population-based surveys and electronic health records regarding the greater levels of mental health symptoms and in recorded diagnoses for common (anxiety, depression) and less common (OCD, eating disorders, SMIs) mental health conditions for people living alone compared to those living with others. Our analyses indicate that mental health conditions are more common among those who live alone compared to those who live with others. Although levels of reported distress increased for both groups during the pandemic, healthcare-seeking dropped in both groups, and the rates of healthcare-seeking among those who live alone converged with those who live with others for common mental health conditions. This suggests greater barriers for treatment access among those that live alone. The findings have implications for mental health service planning and efforts to reduce barriers to treatment access, especially for individuals who live on their own.

## Introduction

More people than ever are living alone, and this increase is projected to continue (1). For instance, recent estimates from the UK suggest that the proportions of lone households are 25.8% in London, and 36.0% in Scotland (2). Studies consistently demonstrate that people who live alone are more likely to experience common (3,4) and severe mental illness (5,6), along with increased self-harm and suicide rates (7).

The COVID-19 pandemic, at least temporarily, changed the context of living alone. Attempts to curb the spread of the virus via lockdowns led to extended periods of greatly reduced in-person social interactions. People living alone during lockdown saw the greatest declines in face-to-face contact (8), and research suggests that alternative means of social interaction (e.g., telephone, video chat) may not protect mental health to the same degree as in-person interaction (9,10). Consequently, those living alone may have experienced a disproportionate increase in mental health difficulties as a result of enforced isolation during the pandemic. However, studies to date, using both convenience and longitudinal samples, have reported mixed findings, with some suggesting no widening of the distress gap between the two groups (11–13), and others noting a steeper rise in psychological distress for people who lived alone in the first months of the pandemic (14–18).

The existing evidence, however, suffers from several key limitations. First, the majority of studies have focussed only on the pandemic and the short-term impact of the pandemic, and few UK studies have evidenced the extent of the gap pre-pandemic and whether the mental health gap between people living alone and living with others changed as the pandemic became prolonged. Second, most studies have only considered a narrow range of mental health outcomes, particularly high-prevalence disorders such as depression and anxiety. To our knowledge, little attention has been given to more severe mental health outcomes, such as eating disorders, obsessive compulsive disorder (OCD), psychoses, and self-harm. Finally, the majority of studies used convenience samples, that may not generalise to the UK population. The combination of limited mental health outcomes studied and convenience sampling makes it difficult to assess if living alone continued to be associated with poor mental health during the COVID-19 pandemic.

Establishing the impact of COVID-19 on population mental health requires rich longitudinal data, with assessments for periods both pre-COVID-19 and during the pandemic. In the UK, such data are available in ongoing population-based longitudinal studies and electronic health records (EHRs). The longitudinal population studies (LPS) include regular assessments of the same individuals and have a wealth of information on living arrangements and mental health prior to and during the pandemic.

However, they are limited to relatively small samples, focusing on common mental health symptoms, and may contain only a limited number of assessments (19). EHRs overcome many of these limitations, in this case allowing us to study the association between living status and mental health outcomes at scale, providing the statistical power and precision to explore the impact of the pandemic on more serious but lower prevalence mental illnesses (e.g., psychosis, OCD) (20). However, EHRs have other limitations as they do not capture psychosocial phenomena such as loneliness and social support, and they capture information only for those who seek healthcare and report symptoms. This may be problematic given that there are many inequalities in who accesses health services for mental health difficulties (21). While EHRs give an indication of the impact of mental health on service use, relying solely on this data source is likely to underestimate mental health problems due to underreporting of common mental health problems, especially for certain subgroups of the population, and particularly during the pandemic, due to closure or reduction of services.

Consequently, this study aimed to provide high-quality evidence on the association between living alone and mental health outcomes before and during the COVID-19 pandemic by estimating these associations in EHRs and 10 UK longitudinal population-based studies, thus adding robustness and information on both need and healthcare utilisation, while balancing the strengths and weaknesses of each data source. Our specific objectives were as follows:

### Objective 1

To describe the gap between those who live alone versus those who live with others, in a range of mental health outcomes in both population-based surveys and recorded within EHRs

### Objective 2

To examine whether there was an effect of the pandemic on the mental health gap between those who live alone and those who live with others by examining pre- and during-pandemic levels of distress in the population and frequency of healthcare contacts due to mental ill-health. We also examine whether the association between living alone and mental health pre and during the pandemic differed by socio-demographic subgroups and according to feelings of loneliness.

## METHODS

### Participants

The population of interest was community-dwelling adults living in private residences.

#### Longitudinal Population Surveys (LPS)

Data were drawn from 10 ongoing longitudinal population studies in the UK that had data available prior to and during the COVID-19 pandemic. The details of each study (design, sample frames, current age range, timing of the most recent pre-pandemic and COVID-19 surveys, response rates and analytical sample size) are presented in Table 1. Five of these studies were age-homogenous birth cohorts and the remaining five covered different age ranges. Ethics statements and data availability information for each study are available in supplementary file 1, and funding statements are available in supplementary file 2. Further details on the cohorts are provided in supplementary file 3.

**Table 1.** Details of each Longitudinal Population Survey including design, timepoints, response rates, measures used and % living alone.

| Study Population | Design and Sample Frame | 2020 Age Range in years | Most Recent Pre-Pandemic Survey | Details of COVID-19 Surveys (Response Rate) | Psychological Distress Measure Used | Analytic N | % Living alone |
| --- | --- | --- | --- | --- | --- | --- | --- |
| NS: Next Steps, formerly known as Longitudinal Study of Young People in England | Sample recruited via secondary schools in England at around age 13 with regular follow-up surveys thereafter. | 29-31 | 2015 | Three surveys: May (20.3%); Sep-Oct (31.8%); Feb-Mar (29%) | General Health Questionnaire 12 (GHQ-12) | 1,262 | 19.3 |
| BCS70: British Cohort Study 1970 | Cohort of all children born in Great Britain (i.e. England, Wales & Scotland) in one week in 1970, with regular follow-up surveys from birth. | 50 | 2016 | Three surveys: May (40.4%); Sep-Oct (43.9%); Feb-Mar (40%) | 9-item Malaise inventory | 2,793 | 8.8 |
| NCDS: National Child Development Study | Cohort of all children born in Great Britain (i.e. England, Wales & Scotland) in one week in 1958, with regular follow-up surveys from birth. | 62 | 2013 | Three surveys: May (57.9%); Sep-Oct (53.9%); Feb-Mar (52%) | 9-item Malaise inventory | 3,772 | 9.8 |
| NSHD: National Survey of Health and Development | Cohort of all children born in Great Britain (i.e. England, Wales & Scotland) in one week in 1946, with regular follow-up surveys from birth. | 74 | 2015 | Three surveys: May (68.2%); Sep-Oct (61.5%); Feb-Mar (90%) | General Health Questionnaire 12 (GHQ-12) | 1,640 | 21.7 |
| ALSPAC G1: Avon Longitudinal Study of Parents and Children- Generation 1 | Cohort of children born in the South-West of England between April 1991 and Dec 1992, with regular follow-up questionnaires from birth. | 27-29 | 2017-2018 | Three surveys: April 2020 (19%); June 2020 (17.4%); December 2020 (26.4%) | Short Mood and Feelings Questionnaire (SMFQ) | 2,252 | 6.2 |
| USoc: Understanding Society: the UK Household Longitudinal Survey | A nationally representative longitudinal household panel study, based on a clustered-stratified probability sample of UK households, with all adults aged 16+ in chosen households surveyed annually. | 16-96 | 2018-2019 | Eight surveys: April (40.3%); May (33.6%); Jun (32.0%); July (31.2%); Sep (29.2%); Nov (27.3%); Jan 2021 (27.2%); Mar 2021 (28.8%) | General Health Questionnaire 12 (GHQ-12) | 12,270 | 18.3 |
| ELSA: English Longitudinal Study of Ageing | A nationally-representative population study of individuals aged 50+ living in England, with biennial surveys and periodic refreshing of the sample to maintain representativeness. | 52-90+ | 2018-2019 | Two surveys: Jun-July (75%); Nov-Dec (73%) | Centre for Epidemiological Studies – Depression (CES-D) | 5,471 | 21.2 |
| GS: Generation Scotland: the Scottish Family Health Study | A family-structured, population-based Scottish cohort, with participants aged 18-99 recruited between 2006-2011. | 27-100 | 2006-2011 | Three surveys: April-Jun 2020 (21.3%); Jul-Aug 2020 (15.4%); Feb 2021 (14.3%) | Patient Health Questionnaire 9 or 8 (PHQ-9 or 8) | 2,984 | 9.1 |
| TwinsUK: the UK Adult Twin Registry | A cohort of volunteer adult TwinsUK (55% monozygotic and 43% dizygotic) from around the United Kingdom who were sampled between 18-101 years of age. | 22-96 | 2017-2018 | Three surveys: April (64.3%); July (77.6%); November (76.1%) | Hospital and Anxiety Depression Scale (HADS) | 2,327 | 20.6 |
| ALSPAC G0: Avon Longitudinal Study of Parents and Children- Generation 0 (Parents) | Parents of the ALSPAC(G1) cohort described above, treated as a separate age-heterogenous study population (original parents). | 45-81 | 2011-2013 | Three questionnaires: April 2020 (12.4%); June 2020 (12.2%); December 2020 (14.3%) | Short Mood and Feelings Questionnaire (SMFQ) | 2,773 | 7.4 |

To be included in the analytic sample, participants were required to have: i) information on living status (alone versus not alone) from early 2020 (the first COVID survey); ii) valid data on our primary outcome (psychological distress) pre-pandemic and during at least one COVID data sweep; and iii) valid data on a key minimum set of covariates (sex, age, ethnicity, education, UK nation, urbanicity, occupational class, housing tenure, chronic illness, see online supplement for details of covariates in each study). Where possible, studies were weighted to be representative of their target population, accounting for sampling design and differential non-response.

#### Electronic Health Records (EHRs)

EHRs managed by the GP software provider TPP were accessed through OpenSAFELY (https://www.opensafely.org/). The dataset covers approximately 24 million people currently registered with primary care practices in England using TPP SystmOne software. Primary care records included coded diagnoses, prescriptions, and physiological measures recorded as part of routine care. Free text data are not available in OpenSAFELY. Data for this study were extracted each month between 1st March 2018 and 31st January 2022 for all adults (aged 18 or older) who were registered with a GP using TPP SystmOne, had at least three months of continuous registration prior to that month, were registered with a TPP practice as of 1st February 2020, and a valid address or postcode. Individuals who met the inclusion criteria were included until the first of: death, de-registration from primary care practice, or the end of the study period. Those living in households with more than 15 individuals or with missing age, sex, Sustainability and Transformation Partnership (STP) region (a geographical area used in NHS administration), or Index of Multiple Deprivation (IMD) were excluded to ensure high data quality without including institutionalised living facilities (e.g., care homes) and omit any institutional effects.

Study size was based on all participants in each cohort for LPS meeting inclusion criteria of data availability (Table 2), and the number of individuals registered with TPP and meeting the inclusion criteria for EHRs.

**Table 2.** Characteristics of patients in OpenSAFELY-TPP in January 2020.

|  | Total population |  | Not living alone |  | Living alone |  |
| --- | --- | --- | --- | --- | --- | --- |
|  | N | (%) | N | (%) | N | (%) |
| **Total population in 2020** | 15,983,045 |  | 12,560,414 |  | 3,422,631 |  |
| **Age group** |  |  |  |  |  |  |
| 18 - 40 years | 5,759,423 | (36.0) | 4,825,262 | (38.4) | 934,161 | (27.3) |
| 41 - 60 years | 5,272,785 | (33.0) | 4,325,383 | (34.4) | 947,402 | (27.7) |
| 61 - 80 years | 3,966,060 | (24.8) | 2,905,601 | (23.1) | 1,060,459 | (31.0) |
| >80 years | 984,777 | (6.20) | 504,168 | (4.0) | 480,609 | (14.0) |
| **Sex** |  |  |  |  |  |  |
| Female | 8,092,393 | (50.6) | 6,347,467 | (50.5) | 1,744,926 | (51.0) |
| Male | 7,890,652 | (49.4) | 6,212,947 | (49.5) | 1,677,705 | (49.0) |
| **Living status** |  |  |  |  |  |  |
| Living alone | 3,422,631 | (21.4) | - |  | 3,422,631 | (100) |
| Not living alone | 12,560,414 | (78.6) | 12,560,414 | (100) | - |  |
| **Ethnicity** |  |  |  |  |  |  |
| White | 11,094,879 | (85.6) | 8,601,261 | (84.7) | 2,493,618 | (89.0) |
| Mixed | 177,748 | (1.4) | 139,583 | (1.4) | 38,165 | (1.4) |
| Asian | 996,754 | (7.7) | 882,723 | (8.7) | 114,031 | (4.1) |
| Black | 351,631 | (2.7) | 276,676 | (2.7) | 74,955 | (2.7) |
| Other | 333,836 | (2.6) | 251,884 | (2.5) | 81,952 | (2.9) |
| Missing | 3,028,197 |  | 2,408,287 |  | 619,910 |  |
| **IMD quintile** |  |  |  |  |  |  |
| 1 (least deprived) | 3,161,810 | (19.8) | 2,455,384 | (19.5) | 706,426 | (20.6) |
| 2 | 3,220,734 | (20.2) | 2,485,660 | (19.8) | 735,074 | (21.5) |
| 3 | 3,410,618 | (21.3) | 2,650,307 | (21.1) | 760,311 | (22.2) |
| 4 | 3,276,903 | (20.5) | 2,596,043 | (20.7) | 680,860 | (19.9) |
| 5 (most deprived) | 2,912,980 | (18.2) | 2,373,020 | (18.9) | 539,960 | (15.8) |
| **Region** |  |  |  |  |  |  |
| East | 3,681,521 | (23.0) | 2,998,303 | (23.9) | 683,218 | (20.0) |
| East Midlands | 2,809,793 | (17.6) | 2,258,073 | (18) | 551,720 | (16.1) |
| London | 1,195,246 | (7.5) | 869,491 | (6.9) | 325,755 | (9.5) |
| North East | 775,155 | (4.8) | 608,067 | (4.8) | 167,088 | (4.9) |
| North West | 1,391,571 | (8.7) | 1,082,407 | (8.6) | 309,164 | (9.0) |
| South East | 1,099,976 | (6.9) | 839,697 | (6.7) | 260,279 | (7.6) |
| South West | 2,196,573 | (13.7) | 1,685,418 | (13.4) | 511,155 | (14.9) |
| West Midlands | 631,974 | (4.0) | 495,883 | (3.9) | 136,091 | (4.0) |
| Yorkshire and The Humber | 2,201,236 | (13.8) | 1,723,075 | (13.7) | 478,161 | (14.0) |
| **Household size** |  |  |  |  |  |  |
| >10 | 94,169 | (0.6) | 94,169 | (0.7) | - |  |
| <=10 | 15,888,876 | (99.4) | 12,466,245 | (99.3) | 3,422,631 | (100) |
| **Household members** |  |  |  |  |  |  |
| All at a TPP practice | 13,773,345 | (86.2) | 10,856,742 | (86.4) | 2,916,603 | (85.2) |
| Not all at a TPP practice | 2,209,700 | (13.8) | 1,703,672 | (13.6) | 506,028 | (14.8) |
| **Advised to shield** | 1,359,711 | (8.5) | 952,960 | (7.6) | 406,751 | (11.9) |
| **Previous mental illness** | 2,193,540 | (13.7) | 1,661,537 | (13.2) | 532,003 | (15.5) |

### Exposure: Living alone or not

Our primary exposure was living status derived in the longitudinal studies at the start of the pandemic between April-June 2020 (this timepoint was used as it was consistent across LPSs and housing status changes later in the pandemic might be affected by mental health) and in EHRs just before the start of the pandemic on 1st February 2020 (identification of the number of individuals in each household was done by TPP independently of the present study, methods are described by Wing et al (22).

Participants were defined as living alone if they had a household size of 1, and were defined as not living alone if they had any value > 1.

### Mental health outcomes

In the LPS, our primary outcomes were measures of psychological distress (i.e., symptoms of common mental health conditions like depression and anxiety) and life satisfaction, which were assessed in a pre-pandemic sweep (T0) and three periods during the pandemic: 1) a period roughly corresponding to the first lockdown (April-June 2020, T1); 2) a period between July-October 2020, when initial restrictions were eased (T2); and 3) a final period (T3) covered November 2020-March 2021, when infection rates rose again, necessitating a second national lockdown. The continuous scales were standardised across the assessment waves within each cohort. We also derived binary outcome variables based on established cut-offs reflecting probable disorder (see supplementary file 4 for criteria). Our secondary outcome, life satisfaction, was measured using questions such as, “Overall, how satisfied are you with your life nowadays?”, with responses indicated on a scale ranging from not at all to completely. Full details of the measures used in each study are available in supplementary file 4.

In EHRs mental health outcomes were considered as any morbidity code recorded in primary or secondary care for diagnoses of depression, self-harm, anxiety, obsessive compulsive disorder, eating disorders, or severe mental illnesses (schizophrenia, bipolar disorder or other psychosis) as defined in table S1. Prescriptions issued to treat these conditions were not included as outcomes in the main analysis. Within each condition separately, a maximum of one event per individual per month was included.

### Analytic Strategy

To answer objective 1 (estimate the mental health gap between people living alone and living with others), we compare the levels of distress and the rates of various mental health conditions in those living alone and those living with others.

To answer objective 2 (whether the gap changed during the pandemic), we use a multi-level modelling approach in the longitudinal studies with a time x living alone interaction to estimate whether there was a change in the difference during the pandemic. For EHRs, we used a quasi-experimental approach with an interrupted time series to estimate monthly period prevalence prior to and during the pandemic while accounting for seasonality, heteroskedasticity, and autocorrelation.

The analysis steps in each data source are described below in greater detail.

#### LPS

First, within each study, we estimated the association between living alone and our mental health outcomes at each timepoint cross-sectionally, using linear regression for continuous outcomes, and logistic regression for binary caseness outcomes. We tested for effect modification by shielding eligibility, loneliness, sex, age-groups, and prior mental health status (caseness at nearest pre-pandemic sweep).

To allow pooling and comparisons of effect sizes across studies, we standardised continuous psychological distress score across all timepoints within each cohort so estimates are interpretable on the same scale. We then estimated longitudinal multi-level models within each cohort, with a time x living alone interaction term to test any changes in the gap during the pandemic.

Both cross-sectional and longitudinal models were estimated unadjusted, and then adjusted for the following covariates as available in each cohort: sex (male; female), age (coded in groups: 16-24, 25-34, 35-44, 45-54, 55-64, 65-74, 75+), education (degree vs. no degree; parental education used for the MCS cohort), ethnicity (white; non-white), UK nation (England, Scotland, Wales or Northern Ireland), area (urban/rural), occupational class (manual; non-manual), home ownership (owned or mortgaged; other), disability (yes; no), prior chronic conditions/illness (yes; no). Full details of modifiers and covariates are presented in supplementary file 5. Models were weighted to be representative of their target population, and account for sampling design and differential non-response, except for TwinsUK, Generation Scotland, and ALSPAC, where no weights were available.

Results from each study were pooled using a random effects meta-analysis with restricted maximum likelihood. Interaction coefficients between time and each of the modifiers were also meta-analysed to inform subsequent stratification. Heterogeneity was explored using the I^2^ statistic. In sensitivity analyses, random-effects meta-regression was conducted to investigate whether between-study heterogeneity could be explained by individual studies’ mental health measurement, time between the pre-pandemic and first pandemic measurement, and representativeness of their target population. All meta-analyses and meta-regressions were conducted in Stata 17.

#### EHRS

We calculated the monthly period prevalence of each mental health outcome by dividing the number of people with the outcome by the total adult population in that month. This was done each month between March 2018 and January 2022. The period prevalence was stratified by those living alone vs living with others. The change in period prevalence was estimated using a linear interrupted time series analysis with Newey-West robust standard errors to account for heteroskedasticity. The interruption was defined to be a binary variable comparing pre vs mid-pandemic periods (i.e., after the start of the COVID-19 pandemic in England on 23^rd^ March 2020). We accounted for seasonal differences in period prevalence by including a categorical variable for spring (March, April, May), summer (June, July, August), autumn (September, October, November), and winter (December, January, February) with spring serving as our baseline. Lastly, we included a lag of 1 to account for autocorrelation between two consecutive months.

Sub-group differences in our results were tested using the same approach while introducing additional stratifying variables for age (using broad groups 18-39, 40-59, 60-79, 80+), sex, STP region, quintile of IMD, ethnicity (White, Mixed, Asian, Black, Other, Unknown), eligibility for shielding, and a binary indicator for history of mental health problems in the five years prior to February 2020. In sensitivity analysis, we performed analyses restricting our outcomes to: (i) those recorded in primary care; and (ii) those recorded in secondary care to explore whether there were differences between primary and secondary care due to the reduction in primary care services during the pandemic (see supplementary table 1). For all variables in the analysis except ethnicity, recording is based on the presence or absence of codes and therefore there are no missing data. Data analysis was carried out in Stata MP 17. Our code with full version control, disorder code lists, and protocol are publicly available on GitHub (https://github.com/opensafely/lone_households).

## RESULTS

Of the 37,544 participants in the LPS, 5,716 lived alone (15.2%), and this figure was 21.4% for patients in the EHR (3,422,631 of 15,983,045). Descriptive statistics for each LPS are presented in Table 1 and Table S2, and for EHRs patient characteristics are given in Table 2.

### Mental health gap between those who live alone and those who live with others

In the LPS, Figure 1 shows modelling estimates for each time-point for both continuous and binary coding of psychological distress (see supplementary tables S3-S8 for full details). At all time points and measures, symptoms were higher for those who lived alone. Before the pandemic, we observed greater mean psychological distress (standardised mean difference [SMD]: 0.09 (95% CI: 0.04, 0.14), higher risk of scoring above the cut-offs reflecting probable disorder (relative risk [RR]: 1.25 (95% CI: 1.12, 1.39) and lower mean life satisfaction (SMD: -0.22 (95% CI: -0.30, -0.15) in those living alone. Full results of the meta-analyses are presented in Figures S1 and table s9.

**Figure 1.**
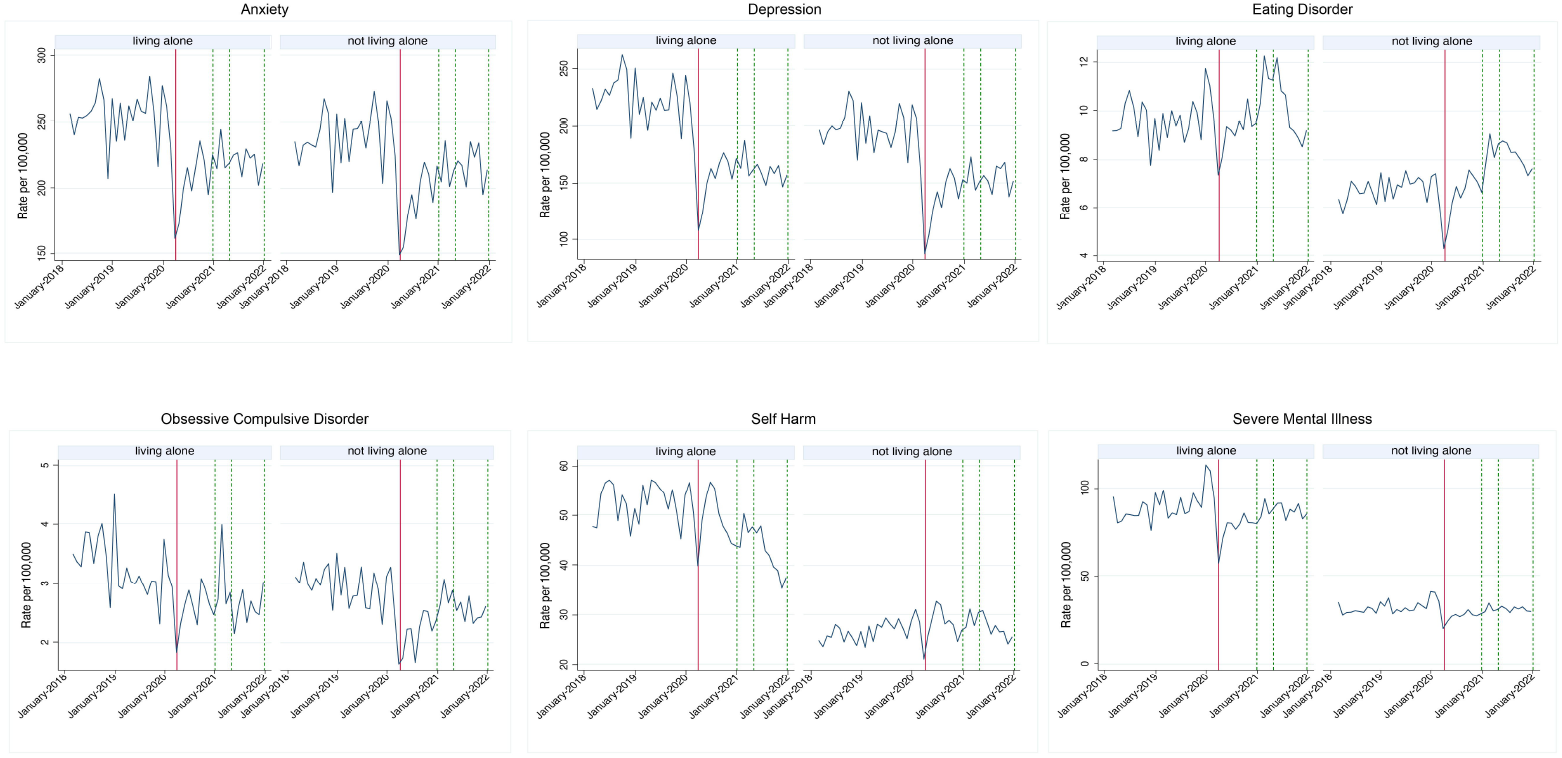
Regression estimates comparing those living alone with those living with others at each timepoint for each longitudinal study and the pooled estimate for the continuous standardised distress scores (left panel) and the binary score (right panel)

In EHRs the pre-pandemic prevalence of mental health outcomes per 100,000 was higher among those living alone compared to those living with others. For example, depression (by 26 cases per 100,000/month, 95% CI 18-33), anxiety (15 cases per 100,000/month, 95% CI 6-21) and self-harm (26 cases per 100,000/month, 95% CI 24-28) (table s10). The difference in eating disorders was on average 3 cases more among those living alone per 100,000 (95% CI 2-3). There was a small average difference of -0.31 cases per 100,000 patients per month (95% CI 0-1) for OCD although statistically significant. For severe mental illness, those living with others had approximately 58 fewer cases of severe mental illness per 100,000 individuals per month than those living alone (95% CI 54-62) (table s10).

### Effect of the pandemic on the mental health gap between those who live alone and those who live with others

As seen in Figure 1, the mental health gaps between groups were similar in magnitude both before and during the pandemic. For the majority of LPS, overall psychological distress and life satisfaction worsened over the course of the pandemic, however there wasn’t consistent evidence of a change in the gap between those living alone and with others (see table S8 for time x living alone interactions).

In the EHRs (Figure 2 and 3), records indicating anxiety and depression across people from all included households dropped during the pandemic, and there was no longer a difference in the gap in prevalence between those living alone and those living with others during the pandemic. For self-harm, while there was a reduction of 8 recorded cases per 100,000 (95% CI 5-11) individuals per month during the pandemic (rSE = 1.44, p<0.001), the difference between people living alone compared to living with others persisted (table s10).

**Figure 2:**
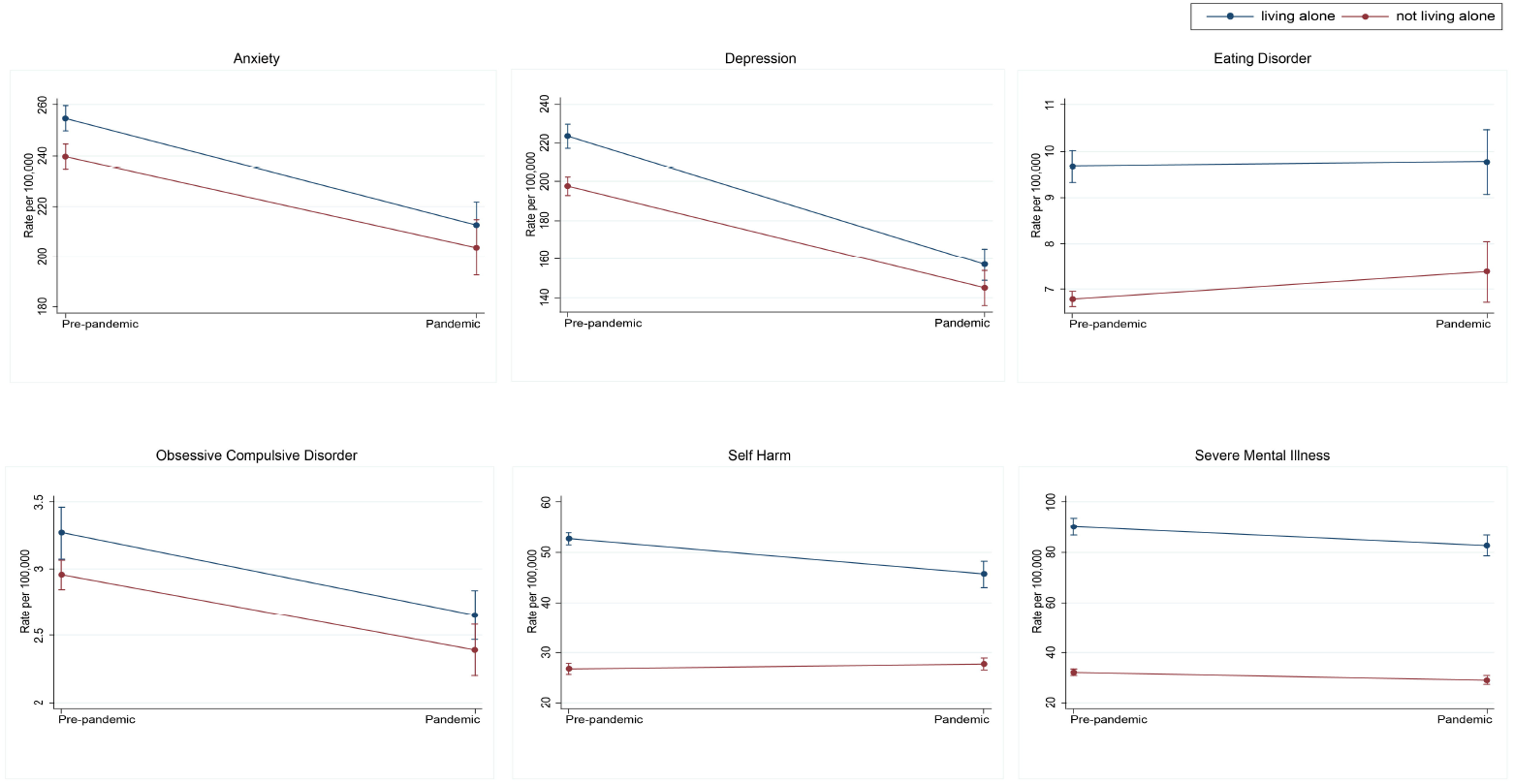
Period prevalence per 100,000 patients by mental health outcome from March 2018 to January 2022 in OpenSAFELY-TPP. Solid red lines indicate the introduction of the first lockdown in England in March 2020. Dotted green lines represent subsequent restrictive measures. All regression estimates can be found under “Main Effects” in Supplementary File 3

**Figure 3:**
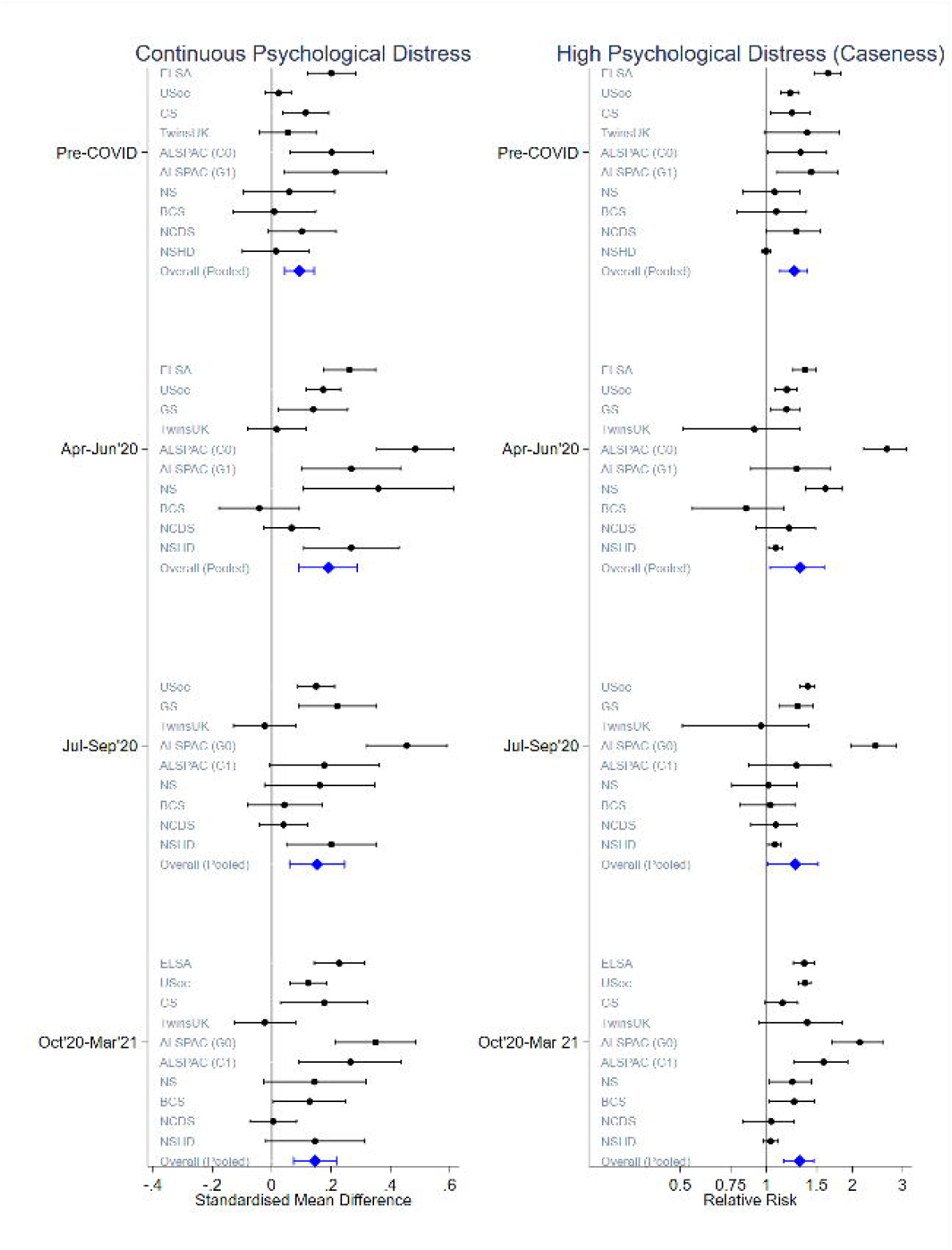
Interrupted time series marginal effects of pre-pandemic vs pandemic periods on outcome measures per 100,000 patients in OpenSAFELY-TPP. Blue lines represent people living alone while red lines are people living with others. Whiskers illustrate 95% confidence intervals

Contrary to anxiety, depression and self-harm, there was no evidence that the pandemic had an effect on monthly prevalence of eating disorder cases recorded in primary or secondary care (table s10). The pre-pandemic difference observed between groups continued after the onset of the pandemic. Notably, the distributions of cases of eating disorder of those living alone versus living with others are dissimilar. While those living with others saw little variance in monthly prevalence prior to the pandemic, dispersion increased during the pandemic. Variation among those living alone was historically more pronounced but more closely resembled the variance of eating disorder in those living with others during pandemic periods.

Period prevalence of OCD presents a similar picture to eating disorders with lower variance in cases among those living with others prior to the pandemic, followed by a notable increase in dispersion during the pandemic. After the onset of the pandemic, there was no detectable difference in OCD between those living alone and those living with others. Furthermore, people from all household sizes saw a drop in recorded monthly OCD prevalence after the start of the pandemic. There was no noticeable pandemic effect on reported severe mental illness among people living with others.

In the LPS, meta-analysis of study specific interaction terms between each modifier of interest (age, sex, shielding status or reported levels of loneliness) and time period indicated that the gap in psychological distress and life satisfaction between those living alone and those living with others did not vary by any of these modifiers. Stratified estimates are displayed in table S6.

In the EHRs, we did observe some differences between subgroups for some mental health outcomes (tables S11-S17). These must be interpreted with caution due to multiple testing and very large sample sizes, leading to small p-values but potentially limited clinical significance.

### Sensitivity analyses

Whilst individual estimates for both high psychological distress and life satisfaction varied between the LPS, meta-regression analysis found that heterogeneity was somewhat explained by the type of mental health measure used, but could not be explained by time between pre- and during-pandemic measures or the representativeness of the studies for their target population (table s18). Leave-one-out meta-analysis found that no individual study significantly skewed the pooled estimates (table s19).

## DISCUSSION

Our study aimed to describe the disparity across a range of mental health outcomes between those who live alone versus with others, and whether the pandemic impacted on this disparity. Using data from longitudinal studies and electronic health records, we found consistent evidence of poorer mental health in people who lived alone prior to the onset of the COVID-19 pandemic. Results from the longitudinal studies showed that the gap for psychological distress persisted through the pandemic. The EHR analyses for depression and anxiety, on the other hand, indicated a narrowing of the mental health gap in recorded cases after the beginning of the pandemic, highlighting the possibility that those living alone, albeit not having reduced need, showed greater reductions in healthcare-seeking and access.

Results from the longitudinal population surveys suggested that common mental health problems, such as symptoms of depression and anxiety, were greater in people who lived alone both prior to and during the pandemic. These symptoms increased for both groups during the pandemic, however, the size of the gap did not vary significantly across the pandemic. Indeed, living alone is unlikely to be a direct risk for mental illness – rather those who live alone may already be more vulnerable to poor mental health, receive less social support and experience higher levels of loneliness (which in turn may lead to greater mental ill-health) (4,23,24). Therefore, the negative impact of living alone on mental health may be mitigated by strong social ties (4,25). A similar gap between lone and non-lone households was identified for life satisfaction, and this difference remained consistent prior to and during the pandemic.

The initial increase in common mental health symptoms did not translate to increased access to mental health care in people who lived alone. Our analyses of EHR records indicated that, while there was a small difference in service use between the two groups pre-pandemic for depression and anxiety, this gap narrowed during the pandemic, while rates of diagnosis plunged for both people who lived alone and with others. The reduction in the gap suggests that there might have been more barriers to help-seeking for those who lived alone compared to those living with others during the pandemic. The overall drop is consistent with other EHR studies that noted a sharp decline in primary care contacts across almost all physical and mental health conditions after the onset of the pandemic (26) and is likely due to barriers to access that were an unintended consequence of the nationwide social restrictions introduced to combat the spread of the virus. It is noteworthy that the rates of diagnosis in EHRs had still not come back to pre-pandemic levels by 2022, albeit their being an increased need as indicated by higher distress levels in surveys.

Looking at lower-prevalence outcomes, results from the EHR analyses indicated that there were higher rates of eating disorders, self-harm, SMI and OCD in people who lived alone prior to the pandemic. While the pandemic saw an overall drop in the number of healthcare contacts for self-harm, SMI and OCD, the relative gaps between the two housing groups remained largely unchanged. However, eating disorders did not see a fall in records during the pandemic. Lastly, EHR analysis revealed a notable increase in variance in rates of anxiety, depression, and eating disorder, seemingly independent of a person’s living status. While period prevalence for these mental health outcomes illustrated high degree of precision prior to the COVID-19 outbreak, this is no longer the case during pandemic, indicating that some individuals continued to consult, while others likely avoided using healthcare. The finding has implications for future healthcare planning, impact, and forecasting studies as estimates may be less reliable, and it is important to understand who was likely to avoid accessing healthcare and why.

The use of different data sources, with their different strengths and sources of bias, permits an examination of the levels of need and healthcare-seeking behaviours, highlighting contrasts in these during the pandemic. Evidence from both data sources demonstrates that those living alone experience greater distress and rates of all examined conditions. However, during the pandemic, there is some indication from the survey data that need might have stayed the same or increased more in those living alone, but this did not translate to more healthcare-seeking behaviours in this group (as reflected in rates in EHRs), and instead for common disorders, the gap narrowed, suggesting potentially greater barriers to healthcare access for this group.

Our study had a number of strengths. By drawing on two distinct forms of data (EHR and longitudinal surveys), we add robustness to our findings by balancing the strengths and weaknesses of each data source. Both had rich data before and during the pandemic. Most of the longitudinal cohorts included were weighted to be representative of their target population and accounted for sampling design and differential non-response. Furthermore, our harmonization strategy across the 10 longitudinal cohorts allowed us to develop comparable exposures, outcomes, and covariates, and pool estimates for similar time periods. However, despite this, the between-study heterogeneity of estimates was large, further highlighting the need to triangulate results from multiple sources, rather than relying on a single data source, when informing policy and health planning. Further benefits unique to our EHR analysis include the statistical power to study serious but low-prevalence mental illness, and provide an indication of service use, which is important information for service planning.

However, the findings of our study should also be considered in light of the following limitations. In both data sources, the proportions of individuals living alone was lower than national estimates (which suggest over a quarter of households are lone households), suggesting people in this group are more likely to drop out from surveys and are potentially missing from formal healthcare or misclassified in the EHRs. This might have led to underestimation of the mental health gap in both data sources. In the LPS, the timing of the assessments, both pre-COVID-19 and during the pandemic, were different between the longitudinal cohorts, although we tried to mitigate this by grouping assessments together into broad timeframes that corresponded with key milestones in the pandemic (e.g., first national lockdown; easing of restrictions; second national lockdown). The survey measures of psychological distress varied across the longitudinal cohorts, and although scores were standardised within each cohort for our analyses, the slightly different symptoms captured by different scales might contribute to heterogeneity in estimates. EHR specific limitations are that records reflect healthcare use which, in turn, depends on several factors including the availability of healthcare, healthcare seeking behaviour, and, for mental health issues in particular, trust in the healthcare system. Differential access to services among people living alone compared to those who don’t could also introduce ascertainment bias in either direction. We only included people who were registered (and therefore with household status recorded) on 1^st^ February 2020, so we did not capture people who moved house during the pandemic, and who could have moved house because they lived alone or were experiencing a mental health problem. Finally, the EHR analysis only had access to the coded GP and hospital records, and some important symptoms of mental health conditions may be recorded as free text (electronic health records also reflect clinician’s diagnostic beliefs and coding practices.), leading to under-ascertainment. However, this GP coding behaviour is unlikely to be differential by household status, although as our findings suggest there still might be important differences in healthcare seeking behaviour by household status.

### Implications and conclusions

Our analyses offer some of the most robust evidence to-date that people who live alone are at increased risk of both common and severe mental illness compared with people who live with others. As living alone continues to increase in many high-income countries, evidence-based interventions which might mitigate its potential adverse mental health effects should be further developed and rolled out. However, we found little evidence to suggest that the medium-term mental health consequences of the pandemic were more keenly felt by those who live alone. Our findings highlight that accounting for household composition and lone households is an important consideration in allocation of services for mental health, especially given the much higher recording of severe mental illnesses in this group. Understanding the drivers and barriers to healthcare-seeking, especially as some of these might be different for those who live alone, will help inform practices to support this vulnerable group better.

## Data Availability

Data for MCS (SN 8682), NS (SN 5545), BCS70 (SN 8547), NCDS (SN 6137) and all four COVID-19 surveys (SN 8658) are available through the UK Data Service. NSHD data are available on request to the NSHD Data Sharing Committee. Interested researchers can apply to access the NSHD data via a standard application procedure. Data requests should be submitted to; further details can be found at http://www.nshd.mrc.ac.uk/data.aspx. doi:10.5522/NSHD/Q101; doi:10.5522/NSHD/Q10.

ALSPAC data is available to researchers through an online proposal system. Information regarding access can be found on the ALSPAC website (http://www.bristol.ac.uk/media-library/sites/alspac/documents/researchers/data-access/ALSPAC_Access_Policy.pdf).

Understanding Society (USoc) data are available through the UK Data Service (SN 6614 and SN 8644).

English Longitudinal Study of Aging (ELSA) data are available through the UK Data Service (SN 8688 and 5050).

Access to Generation Scotland data is approved by the Generation Scotland Access Committee. See https://www.ed.ac.uk/generation-scotland/for-researchers/access or for further details.

The TwinsUK Resource Executive Committee (TREC) oversees management, data sharing and collaborations involving the TwinsUK registry (for further details see https://twinsuk.ac.uk/resources-for-researchers/access-our-data/).

All EHR data were linked, stored and analysed securely within the OpenSAFELY platform https://opensafely.org/. Data include pseudonymized data such as coded diagnoses, medications and physiological parameters. No free text data are included. All code is shared openly for review and re-use under MIT open license ([https://github.com/opensafely/lone_households]). Detailed pseudonymised patient data is potentially re-identifiable and therefore not shared. We rapidly delivered the OpenSAFELY data analysis platform without prior funding to deliver timely analyses on urgent research questions in the context of the global Covid-19 health emergency: now that the platform is established we are developing a formal process for external users to request access in collaboration with NHS England, details of this process are available at OpenSAFELY.org.

## DATA SHARING

Details of data sharing for both the EHRs and LPS used in this study are provided in the supplementary materials.

## SOFTWARE AND REPRODUCIBILITY

For EHRs data management and analysis performed using the OpenSAFELY software libraries and Python, both implemented using Python 3. Code for data management and analysis as well as codelists archived online [https://github.com/opensafely/lone_households]. All iterations of the pre-specified study protocol are archived with version control. Further details are supplementary file 6.

For LPSs datasheets and code are available archived online and available at: https://osf.io/wthpg/?view_only=a3a1b0837bea4e9ea1ace90079821a90

## FUNDING ACKNOWLEDGEMENTS

This work was funded by UK Research and Innovation (UKRI) (COV0076;MR/V015737/1), the Longitudinal Health and Wellbeing strand of the National Core Studies programme (MC_PC_20030: MC_PC_20059: COV-LT-0009).

EH was funded by an NIHR post-doctoral fellowship (PDF-2016-09-029). DMP was funded by an MRC fellowship (MR/W02148X/1). MG, RJSh and SVK acknowledge funding from the Medical Research Council (MC_UU_00022/2) and the Scottish Government Chief Scientist Office (SPHSU17). RJSh additional acknowledges funding from Health Data Research UK (SS005). SVK additionally acknowledges funding from a NRS Senior Clinical Fellowship (SCAF/15/02). RM is supported by Barts Charity (MGU0504). This research will also use data assets made available as part of the Data and Connectivity National Core Study, led by Health Data Research UK in partnership with the Office for National Statistics and funded by UK Research and Innovation (grant ref MC_PC_20058). In addition, the OpenSAFELY Platform is supported by grants from the Wellcome Trust (222097/Z/20/Z); MRC (MR/V015757/1, MC_PC-20059, MR/W016729/1); NIHR (NIHR135559), and Health Data Research UK (HDRUK2021.000, 2021.0157).

## AUTHOR CONTRIBUTIONS

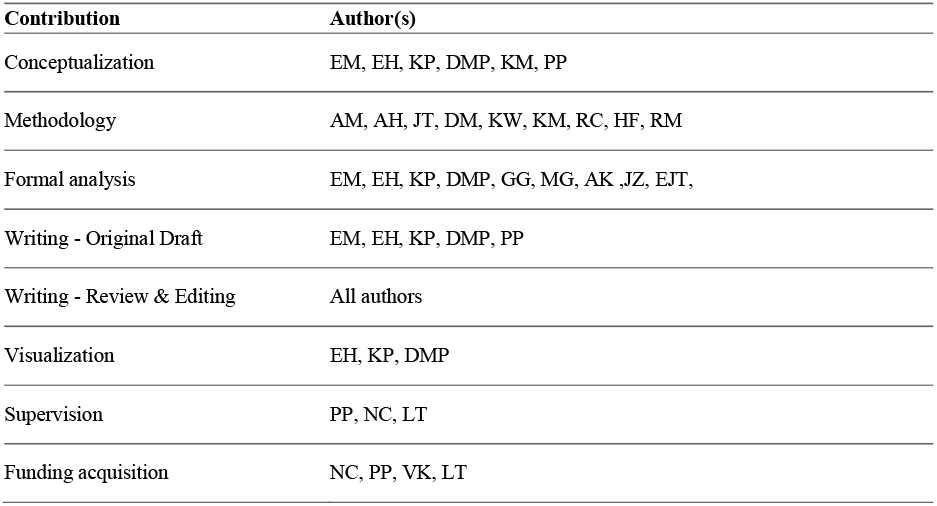

