## supplementary for "Living alone and mental health: parallel analyses in longitudinal population surveys and electronic health records prior to and during the COVID-19 pandemic"

Supplementary materials

### Supplementary file 1. Ethics and data availability statements

The most recent sweeps of the Millennium Cohort Study (MCS), Next Steps (NS), the British Cohort Study 1970 (BCS70), the National Child Development Study (NCDS) and the National Survey of Health and Development (NSHD) and have all been granted ethical approval by the National Health Service (NHS) Research Ethics Committee and all participants have given informed consent. Data for MCS (SN 8682), NS (SN 5545), BCS70 (SN 8547), NCDS (SN 6137) and all four COVID-19 surveys (SN 8658) are available through the UK Data Service. NSHD data are available on request to the NSHD Data Sharing Committee. Interested researchers can apply to access the NSHD data via a standard application procedure. Data requests should be submitted to; further details can be found at http://www.nshd.mrc.ac.uk/data.aspx. doi:10.5522/NSHD/Q101; doi:10.5522/NSHD/Q10.

Ethical approval was obtained from the Avon Longitudinal Study of Parents and Children (ALSPAC) Ethics and Law Committee and the Local Research Ethics Committees. The study website contains details of all the data that is available through a fully searchable data dictionary and variable search tool: http://www.bristol.ac.uk/alspac/researchers/our-data. ALSPAC data is available to researchers through an online proposal system. Information regarding access can be found on the ALSPAC website (http://www.bristol.ac.uk/media-library/sites/alspac/documents/researchers/data-access/ALSPAC_Access_Policy.pdf).

The University of Essex Ethics Committee has approved all data collection for the Understanding Society (USoc) main study and COVID-19 waves. No additional ethical approval was necessary for this secondary data analysis. All data are available through the UK Data Service (SN 6614 and SN 8644).

Waves 1-9 of the English Longitudinal Study of Aging (ELSA) were approved through the National Research Ethics Service, while the COVID-19 Sub-study was approved by the UCL Research Ethics Committee. All participants provided informed consent. All data are available through the UK Data Service (SN 8688 and 5050).

Generation Scotland (GS) obtained ethical approval from the East of Scotland Committee on Medical Research Ethics (on behalf of the National Health Service). Reference number 20/ES/0021. Access to data is approved by the Generation Scotland Access Committee. See https://www.ed.ac.uk/generation-scotland/for-researchers/access or for further details.

All waves of TwinsUK (TwinsUK) have received ethical approval associated with TwinsUK Biobank (19/NW/0187), TwinsUK (EC04/015) or Healthy Ageing Twin Study (H.A.T.S) (07/H0802/84) studies from NHS Research Ethics Committees at the Department of Twin Research and Genetic Epidemiology, King's College London. The TwinsUK Resource Executive Committee (TREC) oversees management, data sharing and collaborations involving the TwinsUK registry (for further details see https://twinsuk.ac.uk/resources-for-researchers/access-our-data/).

### Supplementary file 2. Funding statements for longitudinal population surveys

**ALSPAC:** The UK Medical Research Council and Wellcome (Grant Ref: 217065/Z/19/Z) and the University of Bristol provide core support for ALSPAC. A comprehensive list of grants funding is available on the ALSPAC website (http://www.bristol.ac.uk/alspac/external/documents/grant-acknowledgements.pdf). We are extremely grateful to all the families who took part in this study, the midwives for their help in recruiting them, and the whole ALSPAC team, which includes interviewers, computer and laboratory technicians, clerical workers, research scientists, volunteers, managers, receptionists and nurses. Part of this data was collected using REDCap, see the REDCap website for details: https://projectredcap.org/resources/citations/). This work was supported by Wellcome through the Wellcome Longitudinal Population Studies COVID-19 Secretariat and Steering Group (UK LPS COVID co-ordination, Grant Ref: 221574/Z/20/Z) and supported by the Elizabeth Blackwell Institute, University of Bristol, Wellcome Trust Institutional Strategic Support Fund and Rosetrees Trust (Grant Ref: 204813/Z/16/Z; R105121). ASFK is funded by an Economics and Social Research Council (ESRC) Postdoctoral Fellowship (ES/V011650/1).

**USOC**: Understanding Society is an initiative funded by the Economic and Social Research Council and various Government Departments, with scientific leadership by the Institute for Social and Economic Research, University of Essex, and survey delivery by NatCen Social Research and Kantar Public. The Understanding Society COVID-19 study is funded by the Economic and Social Research Council (ES/K005146/1) and the Health Foundation (2076161). The research data are distributed by the UK Data Service.

**MCS, NS, BCS, NCDS, NSHD**: The Millennium Cohort Study, Next Steps, 1970 British Cohort Study and 1958 National Child Development Study are supported by the Centre for Longitudinal Studies, Resource Centre 2015-20 grant (ES/M001660/1) and a host of other co-funders. The 1946 NSHD cohort is hosted by the the MRC Unit for Lifelong Health and Ageing funded by the Medical Research Council (MC_UU_00019/1Theme 1: Cohorts and Data Collection). The COVID-19 data collections in these five cohorts were funded by the UKRI grant Understanding the economic, social and health impacts of COVID-19 using lifetime data: evidence from 5 nationally representative UK cohorts (ES/V012789/1)

**ELSA**: The English Longitudinal Study of Ageing was developed by a team of researchers based at University College London, NatCen Social Research, the Institute for Fiscal Studies, the University of Manchester and the University of East Anglia. The data were collected by NatCen Social Research. The funding is currently provided by the National Institute on Aging in the US, and a consortium of UK government departments coordinated by the National Institute for Health Research. Funding has also been received by the Economic and Social Research Council. The English Longitudinal Study of Ageing Covid-19 Substudy was supported by the UK Economic and Social Research Grant (ESRC) ES/V003941/1.

**GS**: Generation Scotland received core support from the Chief Scientist Office of the Scottish Government Health Directorates [CZD/16/6] and the Scottish Funding Council [HR03006]. Genotyping of the GS:SFHS samples was carried out by the Genetics Core Laboratory at the Wellcome Trust Clinical Research Facility, Edinburgh, Scotland and was funded by the Medical Research Council UK and the Wellcome Trust (Wellcome Trust Strategic Award “STratifying Resilience and Depression Longitudinally” (STRADL)

Reference 104036/Z/14/Z). Generation Scotland is funded by the Wellcome Trust (216767/Z/19/Z).

**TwinsUK**: TwinsUK receives funding from the Wellcome Trust (WT212904/Z/18/Z), the National Institute for Health Research (NIHR) Biomedical Research Centre based at Guy's and St Thomas' NHS Foundation Trust and King's College London. The TwinsUK COVID-19 personal experience study was funded by the King's Together Rapid COVID-19 Call award, under the projects original title ‘Keeping together through coronavirus: The physical and mental health implications of self-isolation due to the Covid-19 TwinsUK is also supported by the Chronic Disease Research Foundation and Zoe Global Ltd. The funders had no role in study design, data collection and analysis, decision to publish, or preparation of the manuscript.

### Supplementary file 3. Further study information - longitudinal population surveys (as required by the host institutions)

**ALSPAC**: Pregnant women resident in Avon, UK with expected dates of delivery 1st April 1991 to 31st December 1992 were invited to take part in the study. The initial number of pregnancies enrolled is 14,541 (for these at least one questionnaire has been returned or a “Children in Focus” clinic had been attended by 19/07/99). Of these initial pregnancies, there was a total of 14,676 foetuses, resulting in 14,062 live births and 13,988 children who were alive at 1 year of age. When the oldest children were approximately 7 years of age, an attempt was made to bolster the initial sample with eligible cases who had failed to join the study originally. As a result, when considering variables collected from the age of seven onwards (and potentially abstracted from obstetric notes) there are data available for more than the 14,541 pregnancies mentioned above. The number of new pregnancies not in the initial sample (known as Phase I enrolment) that are currently represented on the built files and reflecting enrolment status at the age of 24 is 913 (456, 262 and 195 recruited during Phases II, III and IV respectively), resulting in an additional 913 children being enrolled. The phases of enrolment are described in more detail in the cohort profile paper and its update (see footnote 4 below). The total sample size for analyses using any data collected after the age of seven is therefore 15,454 pregnancies, resulting in 15,589 foetuses. Of these 14,901 were alive at 1 year of age. A 10% sample of the ALSPAC cohort, known as the Children in Focus (CiF) group, attended clinics at the University of Bristol at various time intervals between 4 to 61 months of age. The CiF group were chosen at random from the last 6 months of ALSPAC births (1432 families attended at least one clinic). Excluded were those mothers who had moved out of the area or were lost to follow-up, and those partaking in another study of infant development in Avon.

### Supplementary file 4. Details of measures used in longitudinal population surveys

**MCS:** The K-61 is a 6-item measure of psychological distress (i.e., general anxiety and depression). Responses are rated on a 5-point Likert-type scale, and capture distress over a period of four weeks prior to administration of the scale. Scores range from 0 to 24, with a conservative cut-off of 13+ applied to indicate probable psychological distress.

**ALSPAC:** Self-reported depressive symptoms were measured using the short mood and feelings questionnaire (SMFQ).5 The SMFQ is a 13-item questionnaire that measures the presence of depression symptoms in the previous two weeks and was administered via postal questionnaire or in research clinics. Each item is scored between 0-2, resulting in a summed score between 0-26. Scores of ≥11 on the SMFQ have good specificity and sensitivity for probable depression. In ALSPAC-G0, pre-pandemic depressive symptoms were measured using the Edinburgh Postnatal Depression Scale or EPDS, between 2011-2013. The EPDS consists of 10 items probing depressive symptoms in the last two weeks with scores ranging between 0-30 and higher scores indicating greater depression. Scores ≥ 12 have been validated against probable major depression. The SMFQ was used for pandemic measures of depressive symptoms in ALSPAC-G0.

**NS:** The 12-item General Health Questionnaire (GHQ)3 was used to detect symptoms of psychological distress in NS and USoc. The GHQ is a screening instrument designed to detect symptoms of psychological distress (i.e., general anxiety and depression). Each item is scored 0-3 resulting in scores ranging from 0-36. For classifying participants into high/low psychological distress, we used the common approach of scoring each item as 0-0-1-1, summing the results, and considering scores of 4+ as above the threshold for high psychological distress.

**BCS70, NCDS:** The 9-item version of the Malaise Inventory2 was used to assess general psychological distress. Items are scored using a simple 'Yes/No' response, meaning continuous scores range from 0-9. Scores of four or more are indicative of probable psychiatric distress.

**NSHD:** The General Health Questionnaire (GHQ)3 was also used to detect symptoms of psychological distress in NSHD. For the pre-pandemic sweep, the 28-item GHQ was used, though the 12-item GHQ was used for the subsequent 3 sweeps. A common set of 6 GHQ questions were identified and used in theses analyses. For continuous analyses, each item is scored 0-3 resulting in scores ranging from 0-18. For binary analyses, each item is scored as 0-0-1-1, resulting in scores ranging from 0-6 with a threshold of 2. Sensitivity analyses were conducted to confirm the validity of the 6-item GHQ.

**USoc:** The 12-item General Health Questionnaire (GHQ)3 was used, as described for NS.

**ELSA:** Depressive symptoms were measured using an abbreviated 8-item version of the validated Center for Epidemiologic Studies Depression Scale (CES-D).8 Respondents were asked whether they had experienced any depressive symptoms, such as feeling sad or having restless sleep, in the week prior to interview. For the binary classification, we considered respondents who reported four or more depressive symptoms on the CES-D scale as having elevated depressive symptoms.

**TwinsUK:** The Hospital Anxiety and Depression Scale (HADS)9 is a 14-item scale used to measure levels of psychiatric distress in non-psychiatric patient populations. Responses are indicated on a 4-point ordinal Likert scale.

**GS:** Depression and anxiety were assessed during the pandemic sweeps using the GAD-7 and the Patient Health Questionnaire (PHQ-9)4. The PHQ-9 is a nine-item, validated tool for the assessment of depressive symptoms experienced in the previous two weeks. Participants are asked to indicate how often they have been bothered by problems such as “Little interest or pleasure in doing things?” on a four-point Likert scale from 0 “Not at all” to 3 “Nearly every day”. The score is the sum of the nine items, to a total of 27. A score of 10 or more indicates major depression. The 28-item GHQ3 was used to assess pre-pandemic psychological distress, and so a comparable composite measure was created from the GAD-7 and the PHQ-9 scales to enable evaluation of change over time. Correlations between each GHQ-28 item and items of the GAD-7 and PHQ-9 scales were examined, and the text of the most highly correlated (r > 0.25) questions was then reviewed and sense-checked; matched items are presented in table S1.2.T1 below. Cut-off scores were determined using ROC curves, which found a general cut-off of >4, a depression cut-off of >5 and an anxiety cut-off of >2. The latter matched the general “proportion” of PHQ-9/GAD-7 scores required for a cut-off.

| **Pre-pandemic** | | | | **Pandemic** | | | |
|---|---|---|---|---|---|---|---|
| **Measure** | **Subscale** | **Item** | **Question** | **Measure** | **Subscale** | **Item** | **Question** |
| GHQ-28 | Somatic | 3 | Been feeling run down and out of sorts | PHQ-9 | - | 4 | Feeling tired or having little energy |
| GHQ-28 | Anxiety | 2 | Had difficulty in staying asleep once you are off | PHQ-9 | - | 3 | Trouble falling or staying asleep, or sleeping too much |
| GHQ-28 | Anxiety | 4 | Been getting edgy and bad-tempered | GAD-7 | - | 6 | Becoming easily annoyed or irritable |
| GHQ-28 | Anxiety | 5 | Been getting scared or panicky for no good reason | GAD-7 | - | 1 | Feeling nervous, anxious or on edge |
| GHQ-28 | Depression | 1 | Been thinking of yourself as a worthless person | PHQ-9 | - | 6 | Feeling bad about yourself - or that you are a failure or have let yourself or your family down |
| GHQ-28 | Depression | 4 | Thought of the possibility that you might make away with yourself | PHQ-9 | - | 9 | Thoughts that you would be better off dead or of hurting yourself in some way |
| GHQ-28 | Depression | 5 | Found at times you couldn't do anything because your nerves were too bad | PHQ-9 | - | 7 | Trouble concentrating on things, such as reading the newspaper or watching television |

### Supplementary file 5. Variable coding

| *Variables* | *Study* | *Question(s)* | *Recoding if needed* | *Notes* |
|---|---|---|---|---|
| *** Lone household * 0=Non-lone household; 1=Lives alone** | | | | *** reference period** |
| | **NS<br>BCS 70<br>NCDS<br>NSHD** | How many people do you currently live with? Please include yourself. | 1= 1 (lives alone); 0 = 2+ (non-lone household) | Reference period: T1 = Apr'20 |
| | **USOC** | Thinking about the people who live in your household. Without counting yourself, how many are…? Aged 0-4; Aged 5-15; Aged 16-18; Aged 19-69; Aged 70+ | 0= 1 (lives alone)<br>1+ =0 (lives with others) | Used first valid data from either April, May or June Surveys. |
| | **ELSA** | [If living at usual home or at someone else's home] How many people (including you) are currently living in the residence you are staying in? | 1= 1 (lives alone); 0 = 2+ (non-lone household) | Current situation in Jun/July 2020 |
| | **GS** | 1. Do you currently live alone? 2. Including yourself, how many people live in your household? | 1. Yes = 1 (lives alone)<br>No = 0 (does not live alone)<br>Prefer not to answer = NA (excluded)<br>2. 1 - 12 plus (2-10 = 0; 11+ = excluded) | |
| | **TWINS UK** | Who do you live with? | o I live with other people 1==0<br>o I live by myself 2==1 | |
| | **ALSPAC** | Number of people the participant lives with? | 0 = Live alone; 1 = Live with at least one person | |
| *** Loneliness * 0=Not lonely; 1=Lonely** | | | | |
| | **NS<br>BCS 70<br>NCDS<br>NSHD<br>USOC<br>ELSA** | How often do you feel lonely? 1= Hardly ever; 2= Some of the time; 3= Often | 1+2=0; 3=1 | |

| | | | |
|---|---|---|---|
| | **GS** | How often have you felt lonely during the past week? 1 = None, or almost none of the time; 2 = Some of the time; 3 = Most of the time; 4 = All, or almost all of the time; 99 = Don't know; 98 = Prefer not to answer | 1+2=0; 3+4=1, 98+99 = NA |
| | **TWINS UK** | How often do you feel like this NOW? How often do you feel alone? 1= Hardly ever; 2= Some of the time; 3= Often | 1+2=0; 3=1 |
| | **ALSPAC** | Not Available | |
| *** Sex * 0=Male; 1=Female** | | | |
| | **All** | 0=Male; 1=Female | |
| *** Ethnicity * 0=White; 1=Ethnic Minority** | | Note: Ethnicity is self-reported in these studies | |
| | **NS** | 1=White; 2=Mixed; 3=Indian; 4=Pakistani; 5=Bangladeshi; 6=Black Caribbean; 7=Black African; 8=Other | 1=0; 2/8=1 |
| | **BCS70** | Not Available | |
| | **NCDS** | Not Available | |
| | **NSHD** | Not Available | |
| | **ALSPAC** | G0 (Parents) 1=White; 2=Black carribean; 3=Black african; 4=Other black; 5=Indian; 6=Pakistani; 7=Bangladeshi; 8=Chinese; 9=Other<br>G1 (Children) 1=White; 2=Mixed/Multiple Ethnic group; 3=Asian; 4=Black/African/Caribbean/Black British; 5=Arab or Other | 1=0; 2/9=1 |
| | **USOC** | 1=White British; 2=Irish (White); 3=Gypsy or Irish Traveller (white); 4=Any other white background; 5=White and black caribbean (mixed); 6=White and black african (mixed); 7=White and Asian (mixed); 8=Any other mixed background; 9=Indian (Asian or Asian British); 10=Pakistani (Asian or Asian British); 11=Bangladeshi (Asian or Asian British); 12=Chinese (Asian or Asian British); 13=Any other Asian background (Asian or Asian British); 14=Caribbean (Black or Black British); 15=African (Black or Black British); 16=Any other Black background (Black or Black British); 17=Arab (other Ethnic group); 97=Any other ethnic group | 1-4=0; 5-97=1 |
| | **ELSA** | 1.White; 2=Mixed ethnic group; 3=Black; 4=Black British; 5=Asian; 6=Asian British | 1=0; 2/6=1 |

| | | | |
|---|---|---|---|
| | GS | 1=White Scottish; 2=White English; 3=White Welsh; 4=White N. Irish; 5=White Irish; 6=White Gypsy/Irish traveller; 7=White Polish; 8=Any other white; 9=Asian/British Asian - Indian; 10=Asian/British Asian - Pakistani; 11=Asian/British Asian - Bangladeshi; 12=Asian/British Asian - Chinese; 13=Any other Asian background; 14=Black or Black British - African; 15=Black or Black British - Carribean; 16=Any other Black/African/Caribbean background; 17=Arab or Arab British; 18=Mixed - White and Black Caribbean; 19=Mixed - White and Black African; 20=Mixed - White and Asian; 21=Any other Mixed/Multiple ethnic background; 22=Any other ethnic group | 1-8=0; 9-22=1 |
| | TWINS UK | 1=White- English, Welsh, Scottish, Northern Irish, Irish; 2=White- Other white background; 3=Mixed/multiple ethnic groups - White and Black Caribbean; 4=Mixed/multiple ethnic groups - White and Black African; 5=Mixed/multiple ethnic groups - White and Asian; 6=Mixed/multiple ethnic groups - Other mixed/ multiple ethnic background; 7=Asian/Asian British- Indian; 8=Asian/Asian British - Pakistani; 9=Asian/Asian British - Bangladeshi; 10=Asian/Asian British - Chinese; 11=Asian/Asian British - Other Asian background; 12=Black/Black British - African; 13=Black/Black British - Caribbean; 14=Black/Black British - Other Black Background; 15=Middle-Eastern; 16=Other ethnic group | 1/2=0; 3/16=1 |
| | | Note: Ethnicity is self-reported in these studies | |
| *** Education * 0=No Degree; 1=Degree** | | | |
| | **NS BCS 70 NCDS** | 0=None; 1=Nvq1; 2=Nvq2; 3=Nvq3; 4=Nvq4; 5=Nvq5 *parent's education for MCS | 1/3=0; 4/5=1 |
| | **NSHD** | 0=None attempted; 1.=Vocational course, proficiency only; 2=Sub GCE or sub Burnham C; 3=GCE 'O' level or Burnham C; 4=GCE 'A' Level or Burnham B; 5=Burnham A2; 6= 1st Degree or graduate equivalent; 7= Higher degree, Masters; 8= Higher degree, doctorate; 9=Unknown | 0/5 OR 9=0; 6/8=1 |
| | **ALSPAC** | 1=Degree; 2=A levels/AS levels or equivalent; 3=O levels; 4=Vocational; 5=CSE *parent's education for G1 (Children) | 1=1; 2/5=0 |

| | | | |
|---|---|---|---|
| | **USOC** | 1.Higher degree 2. 1st degree or equivalent 3. Diploma in Higher Education 4. Teaching qualification (not PGCE) 5. Nursing or other medical qualification 6. Other higher degree 7. A-Level 8. Welsh baccalaureate 9. International baccalaureate 10. AS Level 11. Scottish Highers 12. Certificate of 6th year studies 13. GCSE/O-Level 14. Certificate of secondary education 15. Standard or lower 16. Other school certificate 96. No qualifications | 1/6=1; 7/96=0 |
| | **ELSA** | 1=Nvq4/nvq5/degree or equivalent; 2=Higher Education below degree; 3=Nvq3/GCE A level equivalent; 4=Nvq2/GCE O level equivalent; 5=Nvq1/CSE other grade equivalent; 6=Foreign/other; 7=No qualification | 1=1; 2/7=0 |
| | **GS** | 1=No qualifications; 2=Other (please specify); 3=School leavers certificate; 4=CSEs or equivalent; 5=Standard grade, National 4 or 5, O levels, GCSEs or equivalent; 6=Higher grade, A levels, AS levels or equivalent; 7=NVQ or HND or HNC or equivalent; 8=Other professional or technical qualification; 9=Undergraduate degree; 10=Postgraduate degree | 1/8=0; 9 OR 10=1 |
| | **TWINS UK** | 1=No qualification; 2=NVQ1/SVQ1; 3=O-level/GCSE/NVQ2/SVQ2/Scottish intermediate; 4=Scottish Higher, NVQ3, City and Guilds, Pitman; 5=A-level, Scottish Advanced Higher; 6=Higher vocational training (e.g. Diploma, NVQ4, SVQ4); 7=Undergraduate degree; 8=Postgraduate degree (e.g. Masters or PhD), NVQ5, SVQ5 | 1/6=0; 7 OR 8=1 |
| *** Occupational Social Class *<br>1=Managerial/Admin/Professional; 2=Intermediate; 3=Manual/routine; 4=Other/Missing** | | | |
| **NS-SEC: National Statistics Socioeconomic Classification.<br>RGSC: Registrat General's Social Class<br>ONS SOC: Office of National Statistics Standard Occupational Classification** | | | |

| | | |
|---|---|---|
| **NS BCS 70 NCDS** | [NS-SEC] 1=Higher managerial and professional; 2=Lower managerial and rofessional; 3=Intermediate occupations; 4=Small employers and own account workers; 5=Lower supervisory and technical; 6=Semi-routine occupations; 7=Routine occupations; 8=Never worked and long-term unemployed *parent's occupational social class for MCS | 2=1; 3-4=2; 5-7=3; 8=4 |
| **NSHD** | [RGSC] 1=I Professional; 2=II Managerial and Technical; 3=IIINM Skilled non-manual; 4=IIIM Skilled manual; 5=IV Partly skilled; 6=V Unskilled; 7=Missing | 2=1; 3/5=2; 6=3; 7=4 |
| **ALSPAC** | [RGSC] 1=I Professional; 2=II Managerial and Technical; 3=IIINM Skilled non-manual; 4=IIIM Skilled manual; 5=IV Partly skilled; 6=V Unskilled; 7=Armed Forces *parent's occupational social class for G1 (Children) | 2=1; 3/5=2; 6=3; 7=4 |
| **USOC** | [NS-SEC] 1=Higher managerial and professional; 2=Lower managerial and rofessional; 3=Intermediate occupations; 4=Small employers and own account workers; 5=Lower supervisory and technical; 6=Semi-routine occupations; 7=Routine occupations; 8=Never worked or long-term unemployed | 2=1; 3-4=2; 5-7=3; 8=4 |
| **ELSA** | [NS-SEC] 1=Higher managerial and professional; 2=Lower managerial and rofessional; 3=Intermediate occupations; 4=Small employers and own account workers; 5=Lower supervisory and technical; 6=Semi-routine occupations; 7=Routine occupations; 8=Never worked or long-term unemployed | 2=1; 3-4=2; 5-7=3; 8=4 |
| **GS** | [ONS SOC] 1=Managers, directors, senior officials; 2=Associate professional and technical occupations; 3=Administrative and secretarial occupations; 4=Skilled trades occupations; 5=Sales and customer service occupations; 6=Process, plant and machine operatives; 7=Elementary (unskilled) occupations; 8=Never worked | 1/3=1; 4/5=2; 6/7=3; 8=4 |
| **TWINS UK** | Not Available | |

| | |
|---|---|
| *** Shielding Status * 1=Shielding; 0=Not shielding** | |
| | **All studies except ALSPAC and MCS (given their age)** |

| | | | |
|---|---|---|---|
| | **NS**<br>**BCS 70**<br>**NCDS**<br>**NSHD** | [SHIELD] Did you at any time receive a letter or text message from the NHS or Chief Medical Officer saying that you have been identified as someone at risk of severe illness if you catch Coronavirus, because you have an underlying disease or health condition 1. Yes 2. No | 2=0 |
| | **USOC** | Have you received a letter from the NHS or Chief Medical Officer saying that you have been identified as someone at risk of severe illness if you catch coronavirus, because you have an underlying disease or health condition? 1=Yes, 2=No | 2=0 |
| | **ELSA** | “Have you been contacted by the NHS or your GP and advised that you are vulnerable and at risk of severe illness if you catch coronavirus (Covid-19), and should stay at home at all times and avoid any face-to-face contact?” 1=Yes, 2=No | 2=0 |
| | **GS** | Have you been contacted by letter or text message to say you are<br>at severe risk from COVID-19 due to an underlying health condition and should be shielding? 1 = Yes; 2= No. | 2=0 |
| | **TWINS UK** | Wave 2 (July/August 2020) Have you received a letter or text message over the past few months to say you are at high risk from COVID-19 due to an underlying health condition, and should be ‘shielding’? 1 = Yes; 2= No<br>Wave 4 (March/April 2021): Were you contacted by letter or text message to say you are at severe risk from COVID-19 due to an underlying health condition and should be shielding? 1 = Yes; 2= No | |
| | **ALSPAC** | Not Available | |
| *** Home ownership * 1=Home Owner; 0=Not Home Owner** | | | |
| | **NS**<br>**BCS 70**<br>**NCDS**<br>**NSHD** | TENURE \| Do you (or your household) own or rent your home or have some other arrangement? 1. Own - outright 2. Own - buying with help of a mortgage / loan 3. Pay part rent and part mortgage (shared / equity ownership) 4. Rent it 5. Live here rent-free, including rent-free in relative's / friend's / employer’s property; exclude squatting 6. Squatting 7. Other arrangement | 1/2=1; 3/7=0 |

| | | | |
|---|---|---|---|
| | **USOC** | Derived from a series of questions: 1=Owned outright; 2=Owned with mortgage; 3=Local authority rent; 4=Housing association rent; 5=Rented from employer; 6=Rented private unfurnished; 7=Rented private furnished; 8=Other | 1/2=1; 3/8=0 |
|  | **ELSA** | In which of these ways do you occupy this accommodation? 1=Own it outright; 2=Buying it with the help of a mortgage or loan; 3=Pay part rent and part mortgage shared ownership; 4=Rent it; 5=Live here rent free (including rent free in relative's / friend's property; excluding squatting); 6=Squatting | 1/3=1; 4/6=0 |
| | **ALSPAC** | Not Available | |
| | **GS** | What is the status of the accommodation in which you and your household live? 1=Own outright; 2=Own with mortgage; 3=Rent from local authority/housing association; 4=Rent from private landlord or agency; 5=Pay part rent and part mortgage;6=Live rent free; 7=Other; 8=Don't know; 9=Prefer not to answer | 1/2=1; 3/9=0 |
|  | **TWINS UK** | Which describes the home you live in? Owned outright (1); Owned with the help of a mortgage (2); Shared ownership (part owned, part rented) (3); Rented (4); Living rent free (5); Other (Please specify below) (6) | 1:3 = home owner; 4, 5 = non-home owner |
| *** Pre-Pandemic Physical Chronic Condition/Illness *** | | | |
| | **NCDS** | Whether CM has physical/mental health conditions/illnesses expected to last 12+mths. 1=Yes; 2=No | 2=0 |
| | **BCS70** | Any physical/mental health conditions lasting or expected to last 12 months. 1=Yes; 2=No | 2=0 |
| | **NS** | Has longstanding illness. 1=Yes; 2=No | 2=0 |
| | **NSHD** | | |
| | **USOC** | Do you have any long-standing physical or mental impairment, illness or disability? By 'long-standing' I mean anything that has troubled you over a period of at least 12 months or that is likely to trouble you over a period of at least 12 months. 1=Yes, 2=No<br><br>***I also have a variable derived from a list of physical chronic coniditions, let me know if you'd rather I used that*** | 2=0 |

| | | | |
|---|---|---|---|
| | **ELSA** | Q1: Do you have any long-standing illness, disability or infirmity? By long-standing I mean anything that has troubled you over a period of time, or that is likely to affect you over a period of time. 1=Yes; 2=No<br>Q2: (If Yes in Q1) (Does this / Do these) illness(es) or disability(ies) limit your activities in any way? 1=Yes; 2=No | If Q1=1 & Q2=1 |
| | **ALSPAC** | Participant has a history of diabetes, heart disease, hypertension, is overweight, kidney disease, liver disease, anaemia, other lung conditions, cancer, condition affecting the brain and nerves or weakened immune system | 0 = None; 1 = at least 1 |
| | **GS** | Not Available | |
| | **TWINS UK** | Not Available | |
| *** Pre-Pandemic Disability * 1=If respondent disabled/has disability that limits their activity/work; 0=No Disabilities** | | | |
| | **All studies except GS and ALSPAC** | | |
| | **NCDS BCS70 NS** | (Derived) Disability classification Equality act (2010). 0 = Not disabled; 1 = Disabled | |
| | **NSHD** | Do you have any long-term illness, health problem or disability that limits the activities/work you can do? | 0 = No; 1 = Yes |
| | **USOC** | How much difficulty do you have with [separate items for: Mobility; Lifting, carrying or moving objects; Manual dexterity; Continence; Hearing; Sight; Physical Coordination]? Would you say… 1=Some difficulty; 2=A lot of difficulty; 3=Unable to do this | 2/3 on any item=1; else=0 |
| | **ELSA** | Please tell me if you have any difficulty with these everyday activities because of a physical, mental, emotional or memory problem. Exclude any difficulties you expect to last less than three months. 1-15=List of activities; 96=None of the everyday activities listed | 1 if !=96 |
| | **ALSPAC** | Not Available | |
| | **GS** | Not Available | |

| | | |
|---|---|---|
| **TWINS UK** | Do you currently have a long-term disability that seriously restricts your activities? 0 = no;<br>1 = yes | |
| *** Urban/rural * 1=rural; 0=urban** | | |
| **Prioritising T1** | | |
| **NCDS**<br>**BCS70**<br>**NS** | Not Available | |
| **NSHD** | Not Available | |
| **USOC** | Binary indicator classifying the address as falling into an (1) urban or (2) rural area. This is derived from the Office for National Statistics Rural and Urban Classification of Output Areas 2001 (UKDS Study Number 7454). The indicator assumes a value of (1) if the address falls within urban settlements with a population of 10,000 or more, or (2) otherwise. | |
| **ELSA** | Binary indicator classifying the address as falling into an (1) urban or (2) rural area. This is derived from the Office for National Statistics Rural and Urban Classification of Output Areas 2011. The indicator assumes a value of (1) if the address falls within urban settlements with a population of 10,000 or more, or (2) otherwise. | 2=0 |
| **ALSPAC** | Not Available | |
| **GS** | Based on the Scottish Government Urban-Rural Classification 2016. 1= urban; 2 = rural | 2=0 |
| **TWINS UK** | Binary indicator classifying the address as falling into an (1) urban or (2) rural area. This is derived from the Office for National Statistics Rural and Urban Classification of Output Areas 2011. The indicator assumes a value of (1) if the address falls within urban settlements with a population of 10,000 or more, or (2) otherwise. | |

### Supplementary file 6. Further details on Open SAFELY

#### DATA SHARING

All data were linked, stored and analysed securely within the OpenSAFELY platform https://opensafely.org/. Data include pseudonymized data such as coded diagnoses, medications and physiological parameters. No free text data are included. All code is shared openly for review and re-use under MIT open license ([https://github.com/opensafely/lone_households]). Detailed pseudonymised patient data is potentially re-identifiable and therefore not shared. We rapidly delivered the OpenSAFELY data analysis platform without prior funding to deliver timely analyses on urgent research questions in the context of the global Covid-19 health emergency: now that the platform is established we are developing a formal process for external users to request access in collaboration with NHS England; details of this process are available at OpenSAFELY.org.

#### SOFTWARE AND REPRODUCIBILITY

Data management and analysis was performed using the OpenSAFELY software libraries and Python, both implemented using Python 3. This analysis was delivered using federated analysis through the OpenSAFELY platform: codelists and code for data management and data analysis were specified once using the OpenSAFELY tools; then transmitted securely to the OpenSAFELY-TPP platform within TPP's secure environment, and separately to the OpenSAFELY-EMIS platform within EMIS's secure environment, where they were each executed separately against local patient data; summary results were then reviewed for disclosiveness, released, and combined for the final outputs. All code for the OpenSAFELY platform for data management, analysis and secure code execution is shared for review and re-use under open licenses at GitHub.com/OpenSAFELY. Data management was performed using Python 3.8, with analysis carried out using Stata 17. Code for data management and analysis as well as codelists archived online [https://github.com/opensafely/lone_households]. All iterations of the pre-specified study protocol are archived with version control.

#### ETHICS STATEMENT

We received ethics approval to conduct the data linkage and analyses by the London - City & East Research Ethics Committee on the 2nd of April 2020 (REC reference: 20/LO/0651) and LSHTM Ethics Board (ref 21863). No further ethical or research governance approval was required by the University of Oxford but copies of the approval documents were reviewed and held on record.

#### INFORMATION GOVERNANCE

NHS England is the data controller; TPP is the data processor; and the key researchers on OpenSAFELY are acting on behalf of NHS England. This implementation of OpenSAFELY is hosted within the TPP environment, which is accredited to the ISO 27001 information security standard and is NHS IG Toolkit compliant 52,53; patient data have been pseudonymized for analysis and linkage using industry standard cryptographic hashing techniques; all pseudonymized datasets transmitted for linkage onto OpenSAFELY are encrypted; access to the platform is through a virtual private network (VPN) connection;  the researchers hold contracts with NHS England and only access the platform to initiate database queries and statistical models; all database activity is logged; and only aggregate statistical outputs leave the platform environment following best practice for anonymization of results such as statistical disclosure control for low cell counts54. The OpenSAFELY research platform adheres to the data protection principles of the UK Data Protection Act

2018 and the EU General Data Protection Regulation (GDPR) 2016. In March 2020, the Secretary of State for Health and Social Care used powers under the UK Health Service (Control of Patient Information) Regulations 2002 (COPI) to require organizations to process confidential patient information for the purposes of protecting public health, providing healthcare services to the public and monitoring and managing the COVID-19 outbreak and incidents of exposure55. Together, these provide the legal bases to link patient datasets on the OpenSAFELY platform. GP practices, from which the primary care data are obtained, are required to share relevant health information to support the public health response to the pandemic and have been informed of the OpenSAFELY analytics platform.

#### Table S1. Mental health outcomes used in EHR study

| Outcome | Definition | Source |
|---|---|---|
| Depression | Any record of major depressive disorder, dysthymia, mixed anxiety and depression, and adjustment disorders with depressed mood; we will also include codes for depressive symptoms | Primary care |
| Self harm/suicide | Records that indicate explicit or undetermined intention to self-harm, non-suicidal or suicidal self-harm (including overdoses with drugs commonly implicated in suicide, such as paracetamol) | Primary care, hospital admission, emergency care attendance, death records |
| Anxiety | Any record of symptoms or diagnoses of social phobia, agoraphobia, panic, generalised anxiety disorder, and mixed anxiety and depression | Primary care |
| Obsessive compulsive disorder | Codes for hypochondriasis, hoarding disorder, and body focused repetitive behaviour disorders | Primary care |
| Eating disorders | Anorexia nervosa, bulimia nervosa, body dysmorphic disorders, and other specified feeding and eating disorders | Primary care, hospital admission, emergency care attendance, death records |
| Serious mental illness | Diagnoses of schizophrenia and other psychotic disorders, and bipolar disorders | Primary care, hospital admission, emergency care attendance |

#### Table S2. Descriptive statistics for outcome measures in LPS by cohort, time and living status

| | | | *Pre-Pandemic* | | | *Peri-Pandemic 1* | | | *Peri-Pandemic 2* | | | *Peri-Pandemic 3* | | |
|---|---|---|---|---|---|---|---|---|---|---|---|---|---|---|
| **Longitudinal Study** | **Mental Health Outcome** | **Living Alone Status** | **N** | **Mean Score (SD)** | **High Symptoms (%)** | **N** | **Mean Score (SD)** | **High Symptoms (%)** | **N** | **Mean Score (SD)** | **High Symptoms (%)** | **N** | **Mean Score (SD)** | **High Symptoms (%)** |
| **NS** | Psychological Distress | *Living Alone* | 249 | 11.6 (6.2) | 25.9 | 156 | 14.4 (6.6) | 47.8 | 144 | 14.1 (5.8) | 32.3 | 161 | 14.0 (5.9) | 41.3 |
| | | *Living With Others* | 1013 | 11.4 (5.5) | 23.9 | 1104 | 12.8 (5.5) | 30.9 | 1038 | 13.0 (5.4) | 30.6 | 1087 | 13.5 (5.4) | 34.2 |
| | Low Life Satisfaction | *Living Alone* | 249 | 2.2 (0.8) | 23.9 | 161 | 6.4 (2.2) | 40.1 | 145 | 6.8 (1.8) | 33.7 | 160 | 6.4 (1.9) | 42.1 |
| | | *Living With Others* | 1013 | 2.0 (0.8) | 19.8 | 1128 | 7.2 (1.9) | 28.9 | 1042 | 7.2 (1.8) | 26.4 | 1093 | 6.6 (2.1) | 39.9 |
| **BCS** | Psychological Distress | *Living Alone* | 229 | 1.7 (2.0) | 20.2 | 377 | 1.7 (1.9) | 15.8 | 347 | 2.0 (2.3) | 21.2 | 339 | 2.0 (2.2) | 22.6 |
| | | *Living With Others* | 2564 | 1.5 (1.9) | 14.9 | 2558 | 1.7 (2.0) | 16.5 | 2326 | 1.9 (2.0) | 19.2 | 2386 | 1.7 (1.9) | 17.4 |
| | Low Life Satisfaction | *Living Alone* | 229 | 7.6 (1.8) | 35.2 | 382 | 6.8 (2.0) | 17.0 | 348 | 6.6 (2.1) | 39.2 | 341 | 6.5 (2.1) | 42.4 |
| | | *Living With Others* | 2559 | 8.7 (1.6) | 16.5 | 2578 | 7.3 (1.9) | 25.0 | 2330 | 7.3 (1.8) | 24.4 | 2390 | 6.9 (1.9) | 33.5 |
| **NCDS** | Psychological Distress | *Living Alone* | 369 | 1.5 (1.8) | 14.7 | 841 | 1.4 (1.8) | 12.9 | 815 | 1.5 (1.9) | 14.5 | 829 | 1.4 (1.8) | 13.5 |
| | | *Living With Others* | 3403 | 1.8 (1.7) | 10.4 | 2936 | 1.3 (1.8) | 9.1 | 2878 | 1.3 (1.8) | 11.9 | 2895 | 1.3 (1.7) | 11.4 |
| | Low Life Satisfaction | *Living Alone* | 369 | 6.6 1.8) | 39.5 | 860 | 6.9 (2.2) | 32.7 | 815 | 6.9 (2.0) | 34.0 | 834 | 6.6 (2.2) | 41.7 |
| | | *Living With Others* | 3403 | 7.7 (1.5) | 16.4 | 2974 | 7.6 (1.9) | 20.6 | 2882 | 7.6 (1.8) | 20.9 | 2898 | 7.1 (1.9) | 32.1 |
| **NSHD** | Psychological Distress | *Living Alone* | 304 | 4.4 (1.8) | 20.7 | 353 | 6.8 (2.5) | 69.1 | 357 | 6.0 (2.2) | 55.5 | 251 | 5.5 (2.1) | 43.0 |
| | | *Living With Others* | 1127 | 4.2 (1.8) | 16.2 | 1266 | 6.2 (2.2) | 62.2 | 1170 | 5.6 (2.1) | 44.5 | 862 | 5.0 (1.9) | 32.1 |
| | Low Life Satisfaction | *Living Alone* | 264 | 7.7 (2.2) | 15.9 | 252 | 6.9 (2.2) | 23.8 | 363 | 6.9 (2.0) | 25.9 | 251 | 6.4 (2.0) | 32.7 |
| | | *Living With Others* | 998 | 7.7 (1.8) | 12.2 | 859 | 7.7 (1.8) | 12.2 | 1170 | 7.5 (1.9) | 14.0 | 863 | 7.2 (1.9) | 19.2 |
| **ALSPAC-G1** | Psychological Distress | *Living Alone* | 141 | 8.2 (6.4) | 33.3 | 161 | 7.9 (6.5) | 24.2 | 127 | 7.6 (6.0) | 27.6 | 145 | 8.2 (7.0) | 31.7 |
| | | *Living With Others* | 2111 | 6.7 (6.2) | 22.6 | 2632 | 6.1 (5.0) | 17.7 | 1946 | 6.4 (5.7) | 21.1 | 2323 | 6.4 (6.0) | 20.7 |
| | Low Life Satisfaction | *Living Alone* | - | - | - | - | - | - | - | - | - | - | - | - |
| | | *Living With Others* | - | - | - | - | - | - | - | - | - | - | - | - |
| **USOC** | Psychological Distress | *Living Alone* | 3328 | 11.7 (5.4) | 20.7 | 1148 | 13.7 (6.4) | 31.8 | 1089 | 13.3 (6.3) | 26.7 | 1066 | 13.6 (6.3) | 28.2 |
| | | *Living With Others* | 18859 | 11.3 (5.6) | 19.5 | 8508 | 12.4 (6.2) | 24.6 | 7645 | 11.8 (5.7) | 20.0 | 7670 | 12.5 (6.2) | 24.3 |
| | Low Life Satisfaction | *Living Alone* | 3328 | 5.0 (1.4) | 30.5 | 1190 | 4.4 (1.4) | 49.1 | 1093 | 4.5 (1.4) | 45.4 | 1067 | 4.5 (1.4) | 43.7 |
| | | *Living With Others* | 18859 | 5.1 (1.4) | 26.9 | 8942 | 4.8 (1.6) | 35.2 | 7661 | 4.9 (1.5) | 32.0 | 7964 | 4.9 (1.5) | 31.2 |
| **ELSA** | Psychological Distress | *Living Alone* | 1348 | 1.8 (2.3) | 4.1 | 1451 | 2.6 (2.5) | 7.8 | - | - | - | 1303 | 2.9 (2.7) | 9.3 |
| | | *Living With Others* | 4123 | 1.8 (1.6) | 7.2 | 4020 | 1.9 (2.0) | 14.4 | - | - | - | 3742 | 2.1 (2.3) | 18.1 |
| | Low Life Satisfaction | *Living Alone* | 1214 | 6.9 (2.6) | 7.6 | 1447 | 6.7 (2.5) | 10.2 | - | - | - | 1300 | 6.5 (2.5) | 10.9 |
| | | *Living With Others* | 3876 | 7.6 (2.0) | 18.1 | 4004 | 7.1 (2.1) | 24.4 | - | - | - | 3737 | 7.0 (2.2) | 26.2 |
| **GS** | Psychological Distress | *Living Alone* | 452 | 3.0 (3.0) | 21.9 | 433 | 4.1 (4.5) | 24.9 | 334 | 4.1 (4.2) | 32.9 | 304 | 4.9 (4.6) | 40.8 |
| | | *Living With Others* | 2530 | 2.5 (2.4) | 16.9 | 2447 | 3.6 (3.8) | 30.6 | 1771 | 3.2 (3.8) | 26.0 | 1628 | 4.2 (4.1) | 35.1 |
| | Low Life Satisfaction | *Living Alone* | 452 | 7.7 (1.8) | 17.7 | 452 | 5.8 (2.3) | 57.8 | 347 | 6.6 (2.2) | 38.2 | 324 | 5.7 (2.3) | 56.5 |
| | | *Living With Others* | 2528 | 8.2 (1.6) | 11.1 | 2528 | 6.5 (2.2) | 42.7 | 1836 | 7.2 (1.9) | 28.5 | 1700 | 6.3 (2.2) | 47.3 |
| **TWINSUK** | Psychological Distress | *Living Alone* | 480 | 7.5 (5.9) | 7.1 | 480 | 8.5 (6.5) | 6.5 | 480 | 9.76 (5.3) | 5.2 | 480 | 10 (5.7) | 7.1 |
| | | *Living With Others* | 1847 | 7.2 (5.9) | 5.7 | 1847 | 8.6 (6.5) | 8.1 | 1847 | 10 (5.5) | 6 | 1847 | 10.4 (5.7) | 6.7 |
| | Low Life Satisfaction | *Living Alone* | - | - | - | 480 | 6.8 (2.2) | 36.7 | 480 | 7.0 (2.03) | 30 | 480 | 7.2 (2.0) | 25.8 |
| | | *Living With Others* | - | - | - | 1847 | 7.0 (2.0) | 34.2 | 1847 | 7.4 (1.9) | 22.4 | 1847 | 7.5 (1.9) | 23.8 |
| **ALSPAC-G0** | Psychological Distress | *Living Alone* | 205 | 7.4 (6.4) | 23.4 | 237 | 4.9 (4.9) | 11.4 | 213 | 5.0 (5.1) | 12.7 | 219 | 4.7 (5.0) | 12.3 |
| | | *Living With Others* | 2568 | 6.3 (5.2) | 17.3 | 3047 | 2.8 (3.5) | 3.8 | 2614 | 3.0 (3.7) | 4.9 | 2667 | 3.2 (3.9) | 5.8 |
| | Low Life Satisfaction | *Living Alone* | - | - | - | - | - | - | - | - | - | - | - | - |
| | | *Living With Others* | - | - | - | - | - | - | - | - | - | - | - | - |

### Table S3. Regression coefficients from cross-sectional models with continuous outcomes

| cohort | outcome (Z) | TP | exposure | coef | se | l_ci | u_ci | n | Adjusted for |
|---|---|---|---|---|---|---|---|---|---|
| **NCDS** | Psych distress | T0 | T0 living alone | 0.17 | 0.06 | 0.06 | 0.29 | 3772 | -- |
| **NCDS** | Psych distress | T0 | T0 living alone | 0.10 | 0.06 | -0.01 | 0.22 | 3551 | sex, education, UK nation, occupation, housing tenure, disability, chronic illness |
| **NCDS** | Psych distress | T1 | T1 living alone | 0.17 | 0.06 | 0.06 | 0.28 | 3777 | -- |
| **NCDS** | Psych distress | T1 | T1 living alone | 0.07 | 0.05 | -0.03 | 0.16 | 3567 | sex, education, UK nation, occupation, housing tenure, disability, chronic illness |
| **NCDS** | Psych distress | T2 | T1 living alone | 0.11 | 0.04 | 0.02 | 0.19 | 3693 | -- |
| **NCDS** | Psych distress | T2 | T1 living alone | 0.04 | 0.04 | -0.04 | 0.12 | 3692 | sex, education, UK nation, occupation, housing tenure, disability, chronic illness |
| **NCDS** | Psych distress | T3 | T1 living alone | 0.07 | 0.04 | -0.01 | 0.16 | 3724 | -- |
| **NCDS** | Psych distress | T3 | T1 living alone | 0.01 | 0.04 | -0.07 | 0.08 | 3512 | sex, education, UK nation, occupation, housing tenure, disability, chronic illness |
| **NCDS** | Life satisfaction | T0 | T0 living alone | -0.56 | 0.05 | -0.66 | -0.45 | 3772 | -- |
| **NCDS** | Life satisfaction | T0 | T0 living alone | -0.50 | 0.05 | -0.61 | -0.39 | 3551 | sex, education, UK nation, occupation, housing tenure, disability, chronic illness |
| **NCDS** | Life satisfaction | T1 | T1 living alone | -0.37 | 0.07 | -0.50 | -0.24 | 3834 | -- |
| **NCDS** | Life satisfaction | T1 | T1 living alone | -0.32 | 0.06 | -0.43 | -0.21 | 3623 | sex, education, UK nation, occupation, housing tenure, disability, chronic illness |
| **NCDS** | Life satisfaction | T2 | T1 living alone | -0.35 | 0.04 | -0.43 | -0.26 | 3697 | -- |
| **NCDS** | Life satisfaction | T2 | T1 living alone | -0.31 | 0.04 | -0.39 | -0.22 | 3696 | sex, education, UK nation, occupation, housing tenure, disability, chronic illness |
| **NCDS** | Life satisfaction | T3 | T1 living alone | -0.29 | 0.05 | -0.38 | -0.19 | 3732 | -- |
| **NCDS** | Life satisfaction | T3 | T1 living alone | -0.26 | 0.05 | -0.35 | -0.17 | 3520 | sex, education, UK nation, occupation, housing tenure, disability, chronic illness |
| **NSHD** | Psych distress | T0 | T0 living alone | 0.08 | 0.06 | -0.03 | 0.19 | 1431 | -- |
| **NSHD** | Psych distress | T0 | T0 living alone | 0.01 | 0.06 | -0.10 | 0.13 | 1300 | sex, education, UK nation, tenure, disability |
| **NSHD** | Psych distress | T1 | T1 living alone | 0.36 | 0.08 | 0.21 | 0.51 | 1618 | -- |
| **NSHD** | Psych distress | T1 | T1 living alone | 0.27 | 0.08 | 0.11 | 0.43 | 1439 | sex, education, UK nation, tenure, disability |
| **NSHD** | Psych distress | T2 | T1 living alone | 0.32 | 0.07 | 0.18 | 0.47 | 1526 | -- |
| **NSHD** | Psych distress | T2 | T1 living alone | 0.20 | 0.08 | 0.05 | 0.35 | 1445 | sex, education, UK nation, tenure, disability |
| **NSHD** | Psych distress | T3 | T1 living alone | 0.23 | 0.08 | 0.08 | 0.39 | 1112 | -- |
| **NSHD** | Psych distress | T3 | T1 living alone | 0.15 | 0.08 | -0.02 | 0.31 | 1053 | sex, education, UK nation, tenure, disability |
| **GS** | Psych distress | T0 | Live Alone | 0.12 | 0.04 | 0.06 | 0.19 | 2984 | -- |
| **GS** | Psych distress | T1 | Live Alone | 0.13 | 0.06 | 0.02 | 0.24 | 2882 | -- |
| **GS** | Psych distress | T2 | Live Alone | 0.23 | 0.07 | 0.11 | 0.35 | 2106 | -- |
| **GS** | Psych distress | T3 | Live Alone | 0.22 | 0.08 | 0.08 | 0.36 | 1932 | -- |
| **GS** | Life satisfaction | T0 | Live Alone | -- | -- | -- | -- | -- | *No T0 LifeSat score |

| | | | | | | | | | |
|---|---|---|---|---|---|---|---|---|---|
| **GS** | Life satisfaction | T1 | Live Alone | -0.33 | 0.05 | -0.44 | -0.23 | 2982 | -- |
| **GS** | Life satisfaction | T2 | Live Alone | -0.26 | 0.06 | -0.36 | -0.15 | 2184 | -- |
| **GS** | Life satisfaction | T3 | Live Alone | -0.28 | 0.06 | -0.40 | -0.16 | 2024 | -- |
| **GS** | Psych distress | T0 | Live Alone | 0.11 | 0.04 | 0.05 | 0.18 | 2984 | sex, age, education, ethnicity, tenure, occupation, urbanicity |
| **GS** | Psych distress | T1 | Live Alone | 0.14 | 0.06 | 0.03 | 0.25 | 2882 | sex, age, education, ethnicity, tenure, occupation, urbanicity |
| **GS** | Psych distress | T2 | Live Alone | 0.22 | 0.07 | 0.10 | 0.34 | 2106 | sex, age, education, ethnicity, tenure, occupation, urbanicity |
| **GS** | Psych distress | T3 | Live Alone | 0.18 | 0.07 | 0.04 | 0.31 | 1932 | sex, age, education, ethnicity, tenure, occupation, urbanicity |
| **GS** | Life satisfaction | T0 | Live Alone | -- | -- | -- | -- | -- | *No T0 LifeSat score |
| **GS** | Life satisfaction | T1 | Live Alone | -0.33 | 0.05 | -0.43 | -0.23 | 2982 | sex, age, education, ethnicity, tenure, occupation, urbanicity |
| **GS** | Life satisfaction | T2 | Live Alone | -0.27 | 0.06 | -0.37 | -0.16 | 2184 | sex, age, education, ethnicity, tenure, occupation, urbanicity |
| **GS** | Life satisfaction | T3 | Live Alone | -0.25 | 0.06 | -0.37 | -0.13 | 2024 | sex, age, education, ethnicity, tenure, occupation, urbanicity |
| **USOC** | Psych distress | T0 | T0 living alone | 0.05 | 0.02 | 0.01 | 0.10 | 22187 | -- |
| **USOC** | Psych distress | T0 | T0 living alone | 0.02 | 0.02 | -0.02 | 0.07 | 22187 | sex, age, education, ethnicity, UK nation, rural, social class, tenure, disability, illness |
| **USOC** | Psych distress | T1 | T1 living alone | 0.15 | 0.03 | 0.09 | 0.21 | 12088 | -- |
| **USOC** | Psych distress | T1 | T1 living alone | 0.17 | 0.03 | 0.12 | 0.23 | 12088 | sex, age, education, ethnicity, UK nation, rural, social class, tenure, disability, illness |
| **USOC** | Psych distress | T2 | T1 living alone | 0.15 | 0.03 | 0.09 | 0.22 | 9905 | -- |
| **USOC** | Psych distress | T2 | T1 living alone | 0.15 | 0.03 | 0.09 | 0.21 | 9905 | sex, age, education, ethnicity, UK nation, rural, social class, tenure, disability, illness |
| **USOC** | Psych distress | T3 | T1 living alone | 0.12 | 0.03 | 0.06 | 0.18 | 9913 | -- |
| **USOC** | Psych distress | T3 | T1 living alone | 0.12 | 0.03 | 0.06 | 0.18 | 9913 | sex, age, education, ethnicity, UK nation, rural, social class, tenure, disability, illness |
| **USOC** | Life satisfaction | T0 | T0 living alone | -0.09 | 0.02 | -0.14 | -0.05 | 22187 | -- |
| **USOC** | Life satisfaction | T0 | T0 living alone | -0.06 | 0.02 | -0.11 | -0.02 | 22187 | sex, age, education, ethnicity, UK nation, rural, social class, tenure, disability, illness |
| **USOC** | Life satisfaction | T1 | T1 living alone | -0.29 | 0.04 | -0.37 | -0.21 | 10132 | -- |
| **USOC** | Life satisfaction | T1 | T1 living alone | -0.26 | 0.04 | -0.35 | -0.18 | 10132 | sex, age, education, ethnicity, UK nation, rural, social class, tenure, disability, illness |
| **USOC** | Life satisfaction | T2 | T1 living alone | -0.21 | 0.03 | -0.27 | -0.15 | 9905 | -- |
| **USOC** | Life satisfaction | T2 | T1 living alone | -0.17 | 0.03 | -0.22 | -0.11 | 9905 | sex, age, education, ethnicity, UK nation, rural, social class, tenure, disability, illness |
| **USOC** | Life satisfaction | T3 | T1 living alone | -0.24 | 0.03 | -0.29 | -0.18 | 9913 | -- |
| **USOC** | Life satisfaction | T3 | T1 living alone | -0.20 | 0.03 | -0.25 | -0.14 | 9913 | sex, age, education, ethnicity, UK nation, rural, social class, tenure, disability, illness |
| **BCS** | Psych distress | T0 | T0 living alone | 0.08 | 0.08 | -0.08 | 0.24 | 2793 | -- |
| **BCS** | Psych distress | T0 | T0 living alone | 0.01 | 0.07 | -0.13 | 0.15 | 2461 | sex, education, UK nation, occupation, housing tenure, disability, chronic illness |
| **BCS** | Psych distress | T1 | T1 living alone | 0.02 | 0.07 | -0.12 | 0.15 | 2935 | -- |
| **BCS** | Psych distress | T1 | T1 living alone | -0.04 | 0.07 | -0.18 | 0.09 | 2616 | sex, education, UK nation, occupation, housing tenure, disability, chronic illness |
| **BCS** | Psych distress | T2 | T1 living alone | 0.09 | 0.07 | -0.04 | 0.22 | 2673 | -- |

| | | | | | | | | | |
|---|---|---|---|---|---|---|---|---|---|
| **BCS** | Psych distress | T2 | T1 living alone | 0.04 | 0.06 | -0.08 | 0.17 | 2673 | sex, education, UK nation, occupation, housing tenure, disability, chronic illness |
| **BCS** | Psych distress | T3 | T1 living alone | 0.16 | 0.07 | 0.02 | 0.30 | 2725 | -- |
| **BCS** | Psych distress | T3 | T1 living alone | 0.13 | 0.06 | 0.01 | 0.25 | 2406 | sex, education, UK nation, occupation, housing tenure, disability, chronic illness |
| **BCS** | Life satisfaction | T0 | T0 living alone | -0.49 | 0.07 | -0.64 | -0.35 | 2788 | -- |
| **BCS** | Life satisfaction | T0 | T0 living alone | -0.43 | 0.07 | -0.56 | -0.30 | 2457 | sex, education, UK nation, occupation, housing tenure, disability, chronic illness |
| **BCS** | Life satisfaction | T1 | T1 living alone | -0.24 | 0.08 | -0.40 | -0.08 | 2960 | -- |
| **BCS** | Life satisfaction | T1 | T1 living alone | -0.17 | 0.09 | -0.33 | 0.00 | 2638 | sex, education, UK nation, occupation, housing tenure, disability, chronic illness |
| **BCS** | Life satisfaction | T2 | T1 living alone | -0.39 | 0.06 | -0.51 | -0.26 | 2678 | -- |
| **BCS** | Life satisfaction | T2 | T1 living alone | -0.32 | 0.06 | -0.44 | -0.21 | 2678 | sex, education, UK nation, occupation, housing tenure, disability, chronic illness |
| **BCS** | Life satisfaction | T3 | T1 living alone | -0.24 | 0.07 | -0.38 | -0.10 | 2731 | -- |
| **BCS** | Life satisfaction | T3 | T1 living alone | -0.19 | 0.06 | -0.31 | -0.06 | 2412 | sex, education, UK nation, occupation, housing tenure, disability, chronic illness |
| **Next Steps** | Psych distress | T0 | T0 living alone | 0.03 | 0.09 | -0.15 | 0.20 | 1262 | -- |
| **Next Steps** | Psych distress | T0 | T0 living alone | 0.06 | 0.08 | -0.10 | 0.21 | 1114 | sex, ethnicity, education, UK nation, occupation, housing tenure, disability, chronic illness |
| **Next Steps** | Psych distress | T1 | T1 living alone | 0.28 | 0.16 | -0.04 | 0.59 | 1260 | -- |
| **Next Steps** | Psych distress | T1 | T1 living alone | 0.36 | 0.13 | 0.10 | 0.61 | 1135 | sex, ethnicity, education, UK nation, occupation, housing tenure, disability, chronic illness |
| **Next Steps** | Psych distress | T2 | T1 living alone | 0.19 | 0.10 | -0.01 | 0.38 | 1182 | -- |
| **Next Steps** | Psych distress | T2 | T1 living alone | 0.16 | 0.09 | -0.02 | 0.35 | 1182 | sex, ethnicity, education, UK nation, occupation, housing tenure, disability, chronic illness |
| **Next Steps** | Psych distress | T3 | T1 living alone | 0.10 | 0.10 | -0.10 | 0.29 | 1248 | -- |
| **Next Steps** | Psych distress | T3 | T1 living alone | 0.14 | 0.09 | -0.03 | 0.32 | 1103 | sex, ethnicity, education, UK nation, occupation, housing tenure, disability, chronic illness |
| **Next Steps** | Life satisfaction | T0 | T0 living alone | 0.05 | 0.02 | 0.01 | 0.09 | 1262 | -- |
| **Next Steps** | Life satisfaction | T0 | T0 living alone | 0.04 | 0.02 | 0.00 | 0.09 | 1114 | sex, ethnicity, education, UK nation, occupation, housing tenure, disability, chronic illness |
| **Next Steps** | Life satisfaction | T1 | T1 living alone | -0.27 | 0.11 | -0.48 | -0.06 | 1289 | -- |
| **Next Steps** | Life satisfaction | T1 | T1 living alone | -0.25 | 0.11 | -0.47 | -0.04 | 1158 | sex, ethnicity, education, UK nation, occupation, housing tenure, disability, chronic illness |
| **Next Steps** | Life satisfaction | T2 | T1 living alone | -0.14 | 0.06 | -0.27 | -0.02 | 1187 | -- |
| **Next Steps** | Life satisfaction | T2 | T1 living alone | -0.12 | 0.06 | -0.24 | 0.00 | 1187 | sex, ethnicity, education, UK nation, occupation, housing tenure, disability, chronic illness |
| **Next Steps** | Life satisfaction | T3 | T1 living alone | -0.06 | 0.07 | -0.20 | 0.07 | 1253 | -- |
| **Next Steps** | Life satisfaction | T3 | T1 living alone | -0.05 | 0.06 | -0.17 | 0.07 | 1106 | sex, ethnicity, education, UK nation, occupation, housing tenure, disability, chronic illness |
| **ELSA** | Psych distress | T0 | T0 living alone | 0.35 | 0.05 | 0.26 | 0.44 | 5471 | -- |
| **ELSA** | Psych distress | T0 | T0 living alone | 0.20 | 0.04 | 0.12 | 0.28 | 5471 | Sex ; Age; Education; Ethnicity, Occupational class; Home ownership; Disability; chronic il[illegible] |
| **ELSA** | Psych distress | T1 | T1 living alone | 0.37 | 0.05 | 0.28 | 0.47 | 5470 | -- |
| **ELSA** | Psych distress | T1 | T1 living alone | 0.26 | 0.04 | 0.17 | 0.35 | 5470 | Sex ; Age; Education; Ethnicity, Occupational class; Home ownership; Disability; chronic il[illegible] |
| **ELSA** | Psych distress | T3 | T1 living alone | 0.34 | 0.05 | 0.25 | 0.44 | 5046 | -- |

| | | | | | | | | | |
|---|---|---|---|---|---|---|---|---|---|
| **ELSA** | Psych distress | T3 | T1 living alone | 0.23 | 0.04 | 0.14 | 0.31 | 5046 | Sex ; Age; Education; Ethnicity, Occupational class; Home ownership; Disability; chronic il |
| **ELSA** | Life Satisfaction | T0 | T0 living alone | -0.38 | 0.04 | -0.46 | -0.29 | 5090 | -- |
| **ELSA** | Life Satisfaction | T0 | T0 living alone | -0.33 | 0.04 | -0.41 | -0.24 | 5090 | Sex ; Age; Education; Ethnicity, Occupational class; Home ownership; Disability; chronic il |
| **ELSA** | Life Satisfaction | T1 | T1 living alone | -0.16 | 0.04 | -0.25 | -0.08 | 5453 | -- |
| **ELSA** | Life Satisfaction | T1 | T1 living alone | -0.12 | 0.04 | -0.21 | -0.04 | 5453 | Sex ; Age; Education; Ethnicity, Occupational class; Home ownership; Disability; chronic il |
| **ELSA** | Life Satisfaction | T3 | T1 living alone | -0.23 | 0.05 | -0.32 | -0.14 | 5038 | -- |
| **ELSA** | Life Satisfaction | T3 | T1 living alone | -0.21 | 0.05 | -0.30 | -0.12 | 5038 | Sex ; Age; Education; Ethnicity, Occupational class; Home ownership; Disability; chronic il |
| **TwinsUK** | Psych distress | T0 | Live Alone | 0.05 | 0.05 | -0.05 | 0.15 | 2327 | -- |
| **TwinsUK** | Psych distress | T1 | Live Alone | -0.03 | 0.05 | -0.13 | 0.07 | 2323 | -- |
| **TwinsUK** | Psych distress | T2 | Live Alone | -0.05 | 0.06 | -0.16 | 0.06 | 2020 | -- |
| **TwinsUK** | Psych distress | T3 | Live Alone | -0.07 | 0.05 | -0.17 | 0.04 | 2141 | -- |
| **TwinsUK** | Life satisfaction | T0 | Live Alone | -- | -- | -- | -- | | |
| **TwinsUK** | Life satisfaction | T1 | Live Alone | -0.08 | 0.05 | -0.19 | 0.02 | 2327 | -- |
| **TwinsUK** | Life satisfaction | T2 | Live Alone | -0.19 | 0.05 | -0.30 | -0.08 | 2026 | -- |
| **TwinsUK** | Life satisfaction | T3 | Live Alone | -0.12 | 0.05 | -0.22 | -0.02 | 2155 | -- |
| **TwinsUK** | Psych distress | T0 | Live Alone | 0.06 | 0.05 | -0.04 | 0.15 | 2327 | sex, age, ethnicity, edu, country, ruc, homeownership, disability, srh_pp_bin |
| **TwinsUK** | Psych distress | T1 | Live Alone | 0.02 | 0.05 | -0.08 | 0.11 | 2323 | sex, age, ethnicity, edu, country, ruc, homeownership, disability, srh_pp_bin |
| **TwinsUK** | Psych distress | T2 | Live Alone | -0.02 | 0.05 | -0.13 | 0.08 | 2020 | sex, age, ethnicity, edu, country, ruc, homeownership, disability, srh_pp_bin |
| **TwinsUK** | Psych distress | T3 | Live Alone | -0.02 | 0.05 | -0.13 | 0.08 | 2141 | sex, age, ethnicity, edu, country, ruc, homeownership, disability, srh_pp_bin |
| **TwinsUK** | Life satisfaction | T0 | Live Alone | - | - | - | - | - | |
| **TwinsUK** | Life satisfaction | T1 | Live Alone | -0.12 | 0.05 | -0.22 | -0.02 | 2327 | sex, age, ethnicity, edu, country, ruc, homeownership, disability, srh_pp_bin |
| **TwinsUK** | Life satisfaction | T2 | Live Alone | -0.15 | 0.05 | -0.26 | -0.05 | 2026 | sex, age, ethnicity, edu, country, ruc, homeownership, disability, srh_pp_bin |
| **TwinsUK** | Life satisfaction | T3 | Live Alone | -0.13 | 0.05 | -0.24 | -0.03 | 2155 | sex, age, ethnicity, edu, country, ruc, homeownership, disability, srh_pp_bin |
| **ALSPAC-G1** | Psych distress | T0 | Live Alone | 0.23 | 0.08 | 0.07 | 0.40 | 2252 | -- |
| **ALSPAC-G1** | Psych distress | T1 | Live Alone | 0.31 | 0.08 | 0.15 | 0.47 | 2793 | -- |
| **ALSPAC-G1** | Psych distress | T2 | Live Alone | 0.21 | 0.09 | 0.03 | 0.39 | 2073 | -- |
| **ALSPAC-G1** | Psych distress | T3 | Live Alone | 0.28 | 0.08 | 0.12 | 0.45 | 2468 | -- |
| **ALSPAC-G1** | Life satisfaction | T0 | Live Alone | -0.29 | 0.09 | -0.46 | -0.12 | 2178 | -- |
| **ALSPAC-G1** | Life satisfaction | T1 | Live Alone | -0.34 | 0.08 | -0.50 | -0.18 | 2813 | -- |

| | | | | | | | | | |
|---|---|---|---|---|---|---|---|---|---|
| **ALSPAC-G1** | Life satisfaction | T2 | Live Alone | -0.16 | 0.09 | -0.34 | 0.01 | 2080 | -- |
| **ALSPAC-G1** | Life satisfaction | T3 | Live Alone | -0.26 | 0.08 | -0.43 | -0.10 | 2474 | -- |
| **ALSPAC-G1** | Psych distress | T0 | Live Alone | 0.21 | 0.09 | 0.04 | 0.39 | 1809 | sex, ethnicity, occupational class, education, chronic illness |
| **ALSPAC-G1** | Psych distress | T1 | Live Alone | 0.27 | 0.08 | 0.10 | 0.43 | 2064 | sex, ethnicity, occupational class, education, chronic illness |
| **ALSPAC-G1** | Psych distress | T2 | Live Alone | 0.18 | 0.09 | -0.01 | 0.36 | 1610 | sex, ethnicity, occupational class, education, chronic illness |
| **ALSPAC-G1** | Psych distress | T3 | Live Alone | 0.26 | 0.09 | 0.09 | 0.44 | 1919 | sex, ethnicity, occupational class, education, chronic illness |
| **ALSPAC-G1** | Life satisfaction | T0 | Live Alone | -0.27 | 0.09 | -0.45 | -0.09 | 1720 | sex, ethnicity, occupational class, education, chronic illness |
| **ALSPAC-G1** | Life satisfaction | T1 | Live Alone | -0.34 | 0.09 | -0.51 | -0.17 | 2075 | sex, ethnicity, occupational class, education, chronic illness |
| **ALSPAC-G1** | Life satisfaction | T2 | Live Alone | -0.16 | 0.10 | -0.35 | 0.03 | 1616 | sex, ethnicity, occupational class, education, chronic illness |
| **ALSPAC-G1** | Life satisfaction | T3 | Live Alone | -0.29 | 0.09 | -0.47 | -0.12 | 1920 | sex, ethnicity, occupational class, education, chronic illness |
| **ALSPAC-G0** | Psych distress | T0 | Live Alone | 0.23 | 0.07 | 0.09 | 0.37 | 2773 | -- |
| **ALSPAC-G0** | Psych distress | T1 | Live Alone | 0.56 | 0.07 | 0.43 | 0.69 | 3284 | -- |
| **ALSPAC-G0** | Psych distress | T2 | Live Alone | 0.51 | 0.07 | 0.37 | 0.65 | 2827 | -- |
| **ALSPAC-G0** | Psych distress | T3 | Live Alone | 0.38 | 0.07 | 0.24 | 0.51 | 2886 | -- |
| **ALSPAC-G0** | Psych distress | T0 | Live alone | 0.20 | 0.07 | 0.06 | 0.34 | 2702 | sex, age, ethnicity, occupational class, education, chronic illness |
| **ALSPAC-G0** | Psych distress | T1 | Live alone | 0.48 | 0.07 | 0.35 | 0.61 | 3177 | sex, age, ethnicity, occupational class, education, chronic illness |
| **ALSPAC-G0** | Psych distress | T2 | Live alone | 0.46 | 0.07 | 0.32 | 0.59 | 2746 | sex, age, ethnicity, occupational class, education, chronic illness |
| **ALSPAC-G0** | Psych distress | T3 | Live alone | 0.35 | 0.07 | 0.22 | 0.48 | 2808 | sex, age, ethnicity, occupational class, education, chronic illness |

**Note: TP = timepoint; coef = coefficient; se = standard error ; l_ci = lower 95% confidence interval ; u_ci = cupper 95% confidence interval**

#### Table S4 Regression coefficients (non-exponentiated) from cross-sectional models with binary outcomes

| cohort | outcome | TP | exposure | coef | se | l_ci | u_ci | n | Adjusted for |
|---|---|---|---|---|---|---|---|---|---|
| **NCDS** | Psych distress | T0 | T0 living alone | 0.35 | 0.14 | 0.08 | 0.62 | 3772 | -- |
| **NCDS** | Psych distress | T0 | T0 living alone | 0.24 | 0.14 | -0.03 | 0.52 | 3551 | sex, education, UK nation, occupation, housing tenure, disability, chronic illness |
| **NCDS** | Psych distress | T1 | T1 living alone | 0.35 | 0.14 | 0.08 | 0.63 | 3777 | -- |
| **NCDS** | Psych distress | T1 | T1 living alone | 0.18 | 0.14 | -0.10 | 0.47 | 3567 | sex, education, UK nation, occupation, housing tenure, disability, chronic illness |
| **NCDS** | Psych distress | T2 | T1 living alone | 0.20 | 0.10 | 0.00 | 0.40 | 3693 | -- |
| **NCDS** | Psych distress | T2 | T1 living alone | 0.08 | 0.10 | -0.12 | 0.27 | 3692 | sex, education, UK nation, occupation, housing tenure, disability, chronic illness |
| **NCDS** | Psych distress | T3 | T1 living alone | 0.16 | 0.11 | -0.04 | 0.37 | 3724 | -- |
| **NCDS** | Psych distress | T3 | T1 living alone | 0.04 | 0.11 | -0.17 | 0.25 | 3512 | sex, education, UK nation, occupation, housing tenure, disability, chronic illness |
| **NCDS** | Life satisfaction | T0 | T0 living alone | -0.32 | 0.04 | -0.41 | -0.24 | 3772 | -- |
| **NCDS** | Life satisfaction | T0 | T0 living alone | -0.29 | 0.04 | -0.38 | -0.21 | 3551 | sex, education, UK nation, occupation, housing tenure, disability, chronic illness |
| **NCDS** | Life satisfaction | T1 | T1 living alone | -0.17 | 0.03 | -0.23 | -0.10 | 3834 | -- |
| **NCDS** | Life satisfaction | T1 | T1 living alone | -0.13 | 0.03 | -0.20 | -0.07 | 3623 | sex, education, UK nation, occupation, housing tenure, disability, chronic illness |
| **NCDS** | Life satisfaction | T2 | T1 living alone | -0.18 | 0.03 | -0.23 | -0.13 | 3697 | -- |
| **NCDS** | Life satisfaction | T2 | T1 living alone | -0.16 | 0.03 | -0.21 | -0.10 | 3696 | sex, education, UK nation, occupation, housing tenure, disability, chronic illness |
| **NCDS** | Life satisfaction | T3 | T1 living alone | -0.15 | 0.03 | -0.22 | -0.09 | 3732 | -- |
| **NCDS** | Life satisfaction | T3 | T1 living alone | -0.16 | 0.03 | -0.22 | -0.09 | 3520 | sex, education, UK nation, occupation, housing tenure, disability, chronic illness |
| **BCS** | Psych distress | T0 | T0 living alone | 0.30 | 0.17 | -0.04 | 0.64 | 2793 | -- |
| **BCS** | Psych distress | T0 | T0 living alone | 0.08 | 0.15 | -0.22 | 0.38 | 2461 | sex, education, UK nation, occupation, housing tenure, disability, chronic illness |
| **BCS** | Psych distress | T1 | T1 living alone | -0.04 | 0.18 | -0.40 | 0.32 | 2935 | -- |
| **BCS** | Psych distress | T1 | T1 living alone | -0.16 | 0.15 | -0.46 | 0.14 | 2616 | sex, education, UK nation, occupation, housing tenure, disability, chronic illness |
| **BCS** | Psych distress | T2 | T1 living alone | 0.10 | 0.12 | -0.13 | 0.32 | 2673 | -- |
| **BCS** | Psych distress | T2 | T1 living alone | 0.03 | 0.12 | -0.19 | 0.26 | 2673 | sex, education, UK nation, occupation, housing tenure, disability, chronic illness |
| **BCS** | Psych distress | T3 | T1 living alone | 0.26 | 0.13 | 0.01 | 0.51 | 2725 | -- |
| **BCS** | Psych distress | T3 | T1 living alone | 0.22 | 0.11 | 0.00 | 0.45 | 2406 | sex, education, UK nation, occupation, housing tenure, disability, chronic illness |
| **BCS** | Life satisfaction | T0 | T0 living alone | -0.25 | 0.06 | -0.37 | -0.14 | 2788 | -- |
| **BCS** | Life satisfaction | T0 | T0 living alone | -0.20 | 0.05 | -0.29 | -0.10 | 2457 | sex, education, UK nation, occupation, housing tenure, disability, chronic illness |
| **BCS** | Life satisfaction | T1 | T1 living alone | -0.17 | 0.07 | -0.31 | -0.04 | 2960 | -- |
| **BCS** | Life satisfaction | T1 | T1 living alone | -0.10 | 0.06 | -0.21 | 0.01 | 2638 | sex, education, UK nation, occupation, housing tenure, disability, chronic illness |
| **BCS** | Life satisfaction | T2 | T1 living alone | -0.22 | 0.05 | -0.31 | -0.13 | 2678 | -- |

| | | | | | | | | | |
|---|---|---|---|---|---|---|---|---|---|
| **BCS** | Life satisfaction | T2 | T1 living alone | -0.19 | 0.05 | -0.27 | -0.10 | 2678 | sex, education, UK nation, occupation, housing tenure, disability, chronic illness |
| **BCS** | Life satisfaction | T3 | T1 living alone | -0.14 | 0.05 | -0.25 | -0.04 | 2731 | -- |
| **BCS** | Life satisfaction | T3 | T1 living alone | -0.04 | 0.05 | -0.14 | 0.05 | 2412 | sex, education, UK nation, occupation, housing tenure, disability, chronic illness |
| **NS** | Psych distress | T0 | T0 living alone | 0.08 | 0.13 | -0.17 | 0.33 | 1262 | -- |
| **NS** | Psych distress | T0 | T0 living alone | 0.07 | 0.12 | -0.18 | 0.31 | 1114 | sex, ethnicity, education, UK nation, occupation, housing tenure, disability, chronic ill |
| **NS** | Psych distress | T1 | T1 living alone | 0.44 | 0.15 | 0.14 | 0.73 | 1260 | -- |
| **NS** | Psych distress | T1 | T1 living alone | 0.48 | 0.12 | 0.24 | 0.71 | 1135 | sex, ethnicity, education, UK nation, occupation, housing tenure, disability, chronic ill |
| **NS** | Psych distress | T2 | T1 living alone | 0.05 | 0.13 | -0.21 | 0.32 | 1182 | -- |
| **NS** | Psych distress | T2 | T1 living alone | 0.02 | 0.13 | -0.24 | 0.28 | 1182 | sex, ethnicity, education, UK nation, occupation, housing tenure, disability, chronic ill |
| **NS** | Psych distress | T3 | T1 living alone | 0.19 | 0.12 | -0.04 | 0.42 | 1248 | -- |
| **NS** | Psych distress | T3 | T1 living alone | 0.21 | 0.11 | 0.00 | 0.42 | 1103 | sex, ethnicity, education, UK nation, occupation, housing tenure, disability, chronic ill |
| **NS** | Life satisfaction | T0 | T0 living alone | -0.05 | 0.04 | -0.13 | 0.03 | 1262 | -- |
| **NS** | Life satisfaction | T0 | T0 living alone | -0.07 | 0.04 | -0.15 | 0.01 | 1114 | sex, education, UK nation, occupation, housing tenure, disability, chronic illness |
| **NS** | Life satisfaction | T1 | T1 living alone | -0.17 | 0.10 | -0.37 | 0.02 | 1289 | -- |
| **NS** | Life satisfaction | T1 | T1 living alone | -0.15 | 0.11 | -0.38 | 0.07 | 1158 | sex, education, UK nation, occupation, housing tenure, disability, chronic illness |
| **NS** | Life satisfaction | T2 | T1 living alone | -0.11 | 0.07 | -0.24 | 0.03 | 1187 | -- |
| **NS** | Life satisfaction | T2 | T1 living alone | -0.09 | 0.07 | -0.22 | 0.04 | 1187 | sex, education, UK nation, occupation, housing tenure, disability, chronic illness |
| **NS** | Life satisfaction | T3 | T1 living alone | -0.04 | 0.08 | -0.20 | 0.12 | 1253 | -- |
| **NS** | Life satisfaction | T3 | T1 living alone | -0.08 | 0.09 | -0.25 | 0.10 | 1106 | sex, education, UK nation, occupation, housing tenure, disability, chronic illness |
| **NSHD** | Psych distress | T0 | T0 living alone | 0.02 | 0.02 | -0.01 | 0.06 | 1431 | **--** |
| **NSHD** | Psych distress | T0 | T0 living alone | 0.00 | 0.02 | -0.04 | 0.03 | 1300 | sex, education, UK nation, tenure, disability |
| **NSHD** | Psych distress | T1 | T1 living alone | 0.10 | 0.03 | 0.05 | 0.16 | 1618 | **--** |
| **NSHD** | Psych distress | T1 | T1 living alone | 0.08 | 0.03 | 0.02 | 0.14 | 1439 | sex, education, UK nation, tenure, disability |
| **NSHD** | Psych distress | T2 | T1 living alone | 0.10 | 0.03 | 0.05 | 0.16 | 1526 | **--** |
| **NSHD** | Psych distress | T2 | T1 living alone | 0.07 | 0.03 | 0.01 | 0.12 | 1445 | sex, education, UK nation, tenure, disability |
| **NSHD** | Psych distress | T3 | T1 living alone | 0.06 | 0.03 | 0.00 | 0.12 | 1112 | **--** |
| **NSHD** | Psych distress | T3 | T1 living alone | 0.03 | 0.03 | -0.03 | 0.09 | 1053 | sex, education, UK nation, tenure, disability |
| **GS** | Psych distress | T0 | Live Alone | 0.26 | 0.10 | 0.03 | 0.47 | 2984 | -- |
| **GS** | Psych distress | T1 | Live Alone | 0.13 | 0.07 | -0.05 | 0.30 | 2882 | -- |
| **GS** | Psych distress | T2 | Live Alone | 0.23 | 0.09 | 0.02 | 0.44 | 2106 | -- |

| | | | | | | | | | |
|---|---|---|---|---|---|---|---|---|---|
| **GS** | Psych distress | T3 | Live Alone | 0.15 | 0.08 | -0.05 | 0.34 | 1932 | -- |
| **GS** | Life satisfaction | T0 | Live Alone | -- | -- | -- | -- | -- | No T0 Life Satisfaction |
| **GS** | Life satisfaction | T1 | Live Alone | 0.36 | 0.06 | 0.20 | 0.51 | 2982 | -- |
| **GS** | Life satisfaction | T2 | Live Alone | 0.44 | 0.10 | 0.21 | 0.66 | 2184 | -- |
| **GS** | Life satisfaction | T3 | Live Alone | 0.38 | 0.07 | 0.20 | 0.56 | 2024 | -- |
| **GS** | Psych distress | T0 | Live Alone | 0.21 | 0.10 | -0.02 | 0.43 | 2984 | sex, age, education, ethnicity, tenure, occupation, urbanicity |
| **GS** | Psych distress | T1 | Live Alone | 0.16 | 0.07 | -0.02 | 0.34 | 2882 | sex, age, education, ethnicity, tenure, occupation, urbanicity |
| **GS** | Psych distress | T2 | Live Alone | 0.25 | 0.09 | 0.03 | 0.46 | 2106 | sex, age, education, ethnicity, tenure, occupation, urbanicity |
| **GS** | Psych distress | T3 | Live Alone | 0.13 | 0.08 | -0.07 | 0.32 | 1932 | sex, age, education, ethnicity, tenure, occupation, urbanicity |
| **GS** | Life satisfaction | T0 | Live Alone | -- | -- | -- | -- | -- | No T0 Life Satisfaction |
| **GS** | Life satisfaction | T1 | Live Alone | 0.34 | 0.06 | 0.18 | 0.50 | 2982 | sex, age, education, ethnicity, tenure, occupation, urbanicity |
| **GS** | Life satisfaction | T2 | Live Alone | 0.45 | 0.10 | 0.21 | 0.68 | 2184 | sex, age, education, ethnicity, tenure, occupation, urbanicity |
| **GS** | Life satisfaction | T3 | Live Alone | 0.36 | 0.07 | 0.17 | 0.54 | 2024 | sex, age, education, ethnicity, tenure, occupation, urbanicity |
| **USOC** | Psych distress | T0 | T0 living alone | 0.06 | 0.04 | -0.02 | 0.15 | 22187 | -- |
| **USOC** | Psych distress | T1 | T1 living alone | 0.19 | 0.05 | 0.10 | 0.28 | 12088 | -- |
| **USOC** | Psych distress | T2 | T1 living alone | 0.31 | 0.06 | 0.20 | 0.43 | 9905 | -- |
| **USOC** | Psych distress | T3 | T1 living alone | 0.16 | 0.05 | 0.06 | 0.27 | 9913 | -- |
| **USOC** | Life satisfaction | T0 | T0 living alone | 0.13 | 0.04 | 0.06 | 0.20 | 22187 | -- |
| **USOC** | Life satisfaction | T1 | T1 living alone | 0.33 | 0.04 | 0.25 | 0.42 | 10132 | -- |
| **USOC** | Life satisfaction | T2 | T1 living alone | 0.32 | 0.04 | 0.24 | 0.40 | 9905 | -- |
| **USOC** | Life satisfaction | T3 | T1 living alone | 0.31 | 0.03 | 0.24 | 0.38 | 9913 | -- |
| **USOC** | Psych distress | T0 | T0 living alone | 0.04 | 0.04 | -0.04 | 0.13 | 22187 | sex, age, education, ethnicity, UK nation, rural, social class, tenure, disability, illness |
| **USOC** | Psych distress | T1 | T1 living alone | 0.29 | 0.05 | 0.20 | 0.38 | 12088 | sex, age, education, ethnicity, UK nation, rural, social class, tenure, disability, illness |
| **USOC** | Psych distress | T2 | T1 living alone | 0.31 | 0.06 | 0.19 | 0.43 | 9905 | sex, age, education, ethnicity, UK nation, rural, social class, tenure, disability, illness |
| **USOC** | Psych distress | T3 | T1 living alone | 0.18 | 0.05 | 0.07 | 0.29 | 9913 | sex, age, education, ethnicity, UK nation, rural, social class, tenure, disability, illness |
| **USOC** | Life satisfaction | T0 | T0 living alone | 0.04 | 0.04 | -0.03 | 0.11 | 22187 | sex, age, education, ethnicity, UK nation, rural, social class, tenure, disability, illness |
| **USOC** | Life satisfaction | T1 | T1 living alone | 0.29 | 0.04 | 0.20 | 0.37 | 10132 | sex, age, education, ethnicity, UK nation, rural, social class, tenure, disability, illness |
| **USOC** | Life satisfaction | T2 | T1 living alone | 0.23 | 0.04 | 0.15 | 0.31 | 9905 | sex, age, education, ethnicity, UK nation, rural, social class, tenure, disability, illness |
| **USOC** | Life satisfaction | T3 | T1 living alone | 0.23 | 0.04 | 0.16 | 0.30 | 9913 | sex, age, education, ethnicity, UK nation, rural, social class, tenure, disability, illness |
| **ELSA** | Psych distress | T0 | T0 living alone | 0.75 | 0.09 | 0.58 | 0.93 | 5471 | **--** |
| **ELSA** | Psych distress | T0 | T0 living alone | 0.50 | 0.09 | 0.32 | 0.67 | 5471 | Sex ; Age; Education; Ethnicity, Occupational class; Home ownership; Disability; Ch |
| **ELSA** | Psych distress | T1 | T1 living alone | 0.48 | 0.07 | 0.35 | 0.61 | 5470 | **--** |

| | | | | | | | | | |
|---|---|---|---|---|---|---|---|---|---|
| **ELSA** | Psych distress | T1 | T1 living alone | 0.31 | 0.06 | 0.19 | 0.44 | 5470 | Sex ; Age; Education; Ethnicity, Occupational class; Home ownership; Disability; Chr |
| **ELSA** | Psych distress | T3 | T1 living alone | 0.42 | 0.06 | 0.30 | 0.54 | 5046 | **--** |
| **ELSA** | Psych distress | T3 | T1 living alone | 0.31 | 0.06 | 0.19 | 0.42 | 5046 | Sex ; Age; Education; Ethnicity, Occupational class; Home ownership; Disability; Chr |
| **ELSA** | Low Life Satisfaction | T0 | T0 living alone | 0.49 | 0.06 | 0.37 | 0.60 | 5090 | **--** |
| **ELSA** | Low Life Satisfaction | T0 | T0 living alone | 0.37 | 0.06 | 0.26 | 0.49 | 5090 | Sex ; Age; Education; Ethnicity, Occupational class; Home ownership; Disability; Chr |
| **ELSA** | Low Life Satisfaction | T1 | T1 living alone | 0.21 | 0.05 | 0.11 | 0.31 | 5453 | **--** |
| **ELSA** | Low Life Satisfaction | T1 | T1 living alone | 0.14 | 0.05 | 0.03 | 0.24 | 5453 | Sex ; Age; Education; Ethnicity, Occupational class; Home ownership; Disability; Chr |
| **ELSA** | Low Life Satisfaction | T3 | T1 living alone | 0.24 | 0.05 | 0.14 | 0.34 | 5038 | **--** |
| **ELSA** | Low Life Satisfaction | T3 | T1 living alone | 0.19 | 0.05 | 0.08 | 0.29 | 5038 | Sex ; Age; Education; Ethnicity, Occupational class; Home ownership; Disability; Chr |
| **TwinsUK** | Psych distress | T0 | Live Alone | 0.27 | 0.25 | -0.22 | 0.75 | 2327 | -- |
| **TwinsUK** | Psych distress | T1 | Live Alone | -0.23 | 0.20 | -0.62 | 0.16 | 2327 | -- |
| **TwinsUK** | Psych distress | T2 | Live Alone | -0.06 | 0.26 | -0.57 | 0.45 | 2026 | -- |
| **TwinsUK** | Psych distress | T3 | Live Alone | 0.13 | 0.24 | -0.35 | 0.60 | 2148 | -- |
| **TwinsUK** | Life satisfaction | T0 | Live Alone | - | - | - | - | - | |
| **TwinsUK** | Life satisfaction | T1 | Live Alone | -0.04 | 0.06 | -0.16 | 0.09 | 2327 | |
| **TwinsUK** | Life satisfaction | T2 | Live Alone | -0.13 | 0.07 | -0.26 | 0.01 | 2026 | |
| **TwinsUK** | Life satisfaction | T3 | Live Alone | -0.03 | 0.06 | -0.16 | 0.09 | 2155 | |
| **TwinsUK** | Psych distress | T0 | Live Alone | 0.33 | 0.21 | -0.08 | 0.74 | 2327 | sex, age, ethnicity, edu, country, ruc, homeownership, disability, srh_pp_bin |
| **TwinsUK** | Psych distress | T1 | Live Alone | -0.10 | 0.20 | -0.49 | 0.30 | 2327 | sex, age, ethnicity, edu, country, ruc, homeownership, disability, srh_pp_bin |
| **TwinsUK** | Psych distress | T2 | Live Alone | -0.04 | 0.23 | -0.49 | 0.40 | 2026 | sex, age, ethnicity, edu, country, ruc, homeownership, disability, srh_pp_bin |
| **TwinsUK** | Psych distress | T3 | Live Alone | 0.33 | 0.23 | -0.12 | 0.78 | 2148 | sex, age, ethnicity, edu, country, ruc, homeownership, disability, srh_pp_bin |
| **TwinsUK** | Life satisfaction | T0 | Live Alone | - | - | - | - | | |
| **TwinsUK** | Life satisfaction | T1 | Live Alone | -0.13 | 0.05 | -0.24 | -0.03 | 2327 | sex, age, ethnicity, edu, country, ruc, homeownership, disability, srh_pp_bin |
| **TwinsUK** | Life satisfaction | T2 | Live Alone | -0.17 | 0.06 | -0.28 | -0.06 | 2026 | sex, age, ethnicity, edu, country, ruc, homeownership, disability, srh_pp_bin |
| **TwinsUK** | Life satisfaction | T3 | Live Alone | -0.14 | 0.05 | -0.24 | -0.04 | 2155 | sex, age, ethnicity, edu, country, ruc, homeownership, disability, srh_pp_bin |
| **ALSPAC-G1** | Psych distress | T0 | Live Alone | 0.39 | 0.15 | 0.09 | 0.69 | 2252 | -- |
| **ALSPAC-G1** | Psych distress | T1 | Live Alone | 0.31 | 0.17 | -0.01 | 0.64 | 2793 | -- |
| **ALSPAC-G1** | Psych distress | T2 | Live Alone | 0.27 | 0.18 | -0.08 | 0.61 | 2073 | -- |
| **ALSPAC-G1** | Psych distress | T3 | Live Alone | 0.43 | 0.15 | 0.12 | 0.73 | 2468 | -- |
| **ALSPAC-G1** | Life satisfaction | T0 | Live Alone | 0.39 | 0.18 | 0.04 | 0.75 | 2178 | -- |

| | | | | | | | | | |
|---|---|---|---|---|---|---|---|---|---|
| **ALSPAC-G1** | Life satisfaction | T1 | Live Alone | 0.31 | 0.13 | 0.06 | 0.56 | 2813 | -- |
| **ALSPAC-G1** | Life satisfaction | T2 | Live Alone | 0.26 | 0.14 | -0.02 | 0.54 | 2080 | -- |
| **ALSPAC-G1** | Life satisfaction | T3 | Live Alone | 0.34 | 0.13 | 0.08 | 0.60 | 2474 | -- |
| **ALSPAC-G1** | Psych distress | T0 | Live Alone | 0.36 | 0.18 | 0.02 | 0.70 | 1809 | sex, ethnicity, occupational class, education, chronic illness |
| **ALSPAC-G1** | Psych distress | T1 | Live Alone | 0.24 | 0.20 | -0.15 | 0.64 | 2064 | sex, ethnicity, occupational class, education, chronic illness |
| **ALSPAC-G1** | Psych distress | T2 | Live Alone | 0.24 | 0.21 | -0.17 | 0.65 | 1610 | sex, ethnicity, occupational class, education, chronic illness |
| **ALSPAC-G1** | Psych distress | T3 | Live Alone | 0.46 | 0.17 | 0.12 | 0.80 | 1919 | sex, ethnicity, occupational class, education, chronic illness |
| **ALSPAC-G1** | Life satisfaction | T0 | Live Alone | 0.32 | 0.21 | -0.09 | 0.73 | 1720 | sex, ethnicity, occupational class, education, chronic illness |
| **ALSPAC-G1** | Life satisfaction | T1 | Live Alone | 0.36 | 0.14 | 0.08 | 0.64 | 2075 | sex, ethnicity, occupational class, education, chronic illness |
| **ALSPAC-G1** | Life satisfaction | T2 | Live Alone | 0.22 | 0.17 | -0.11 | 0.55 | 1616 | sex, ethnicity, occupational class, education, chronic illness |
| **ALSPAC-G1** | Life satisfaction | T3 | Live Alone | 0.41 | 0.15 | 0.12 | 0.70 | 1920 | sex, ethnicity, occupational class, education, chronic illness |
| **ALSPAC-G0** | Psych distress | T0 | Live Alone | 0.31 | 0.15 | 0.01 | 0.60 | 2773 | -- |
| **ALSPAC-G0** | Psych distress | T1 | Live Alone | 1.09 | 0.21 | 0.67 | 1.51 | 3284 | -- |
| **ALSPAC-G0** | Psych distress | T2 | Live Alone | 0.94 | 0.21 | 0.53 | 1.36 | 2827 | -- |
| **ALSPAC-G0** | Psych distress | T3 | Live Alone | 0.76 | 0.21 | 0.35 | 1.17 | 2886 | -- |
| **ALSPAC-G0** | Psych distress | T0 | Live alone | 0.28 | 0.16 | -0.03 | 0.58 | 2702 | sex, age, ethnicity, occupational class, education, chronic illness |
| **ALSPAC-G0** | Psych distress | T1 | Live alone | 0.97 | 0.23 | 0.52 | 1.43 | 3177 | sex, age, ethnicity, occupational class, education, chronic illness |
| **ALSPAC-G0** | Psych distress | T2 | Live alone | 0.88 | 0.22 | 0.44 | 1.31 | 2746 | sex, age, ethnicity, occupational class, education, chronic illness |
| **ALSPAC-G0** | Psych distress | T3 | Live alone | 0.75 | 0.22 | 0.32 | 1.18 | 2808 | sex, age, ethnicity, occupational class, education, chronic illness |

**Note: TP = timepoint; coef = coefficient; se = standard error ; l_ci = lower 95% confidence interval ; u_ci = cupper 95% confidence interval**

### Table S5. Interactions effects from cross-sectional models with continuous outcomes

| cohort | outcome (Z) | TP | exposure | modifier | modifier category 1 | coef_interaction | coef_se | lower_ci | upper_ci | n |
|---|---|---|---|---|---|---|---|---|---|---|
| **NCDS** | Psych distress | 0 | 0 living alone | loneliness at 0 | lonely | -0.31 | 0.23 | -0.76 | 0.13 | 3448 |
| **NCDS** | Psych distress | 0 | 0 living alone | shielding status at 1 | shielding | -0.13 | 0.20 | -0.52 | 0.26 | 3527 |
| **NCDS** | Psych distress | 0 | 0 living alone | sex | female | -0.21 | 0.12 | -0.44 | 0.02 | 3551 |
| **NCDS** | Psych distress | 0 | 0 living alone | Prior mental ill-health | yes | -0.14 | 0.10 | -0.34 | 0.07 | 3551 |
| **NCDS** | Psych distress | 1 | 1 living alone | loneliness at 1 | lonely | -0.42 | 0.23 | -0.88 | 0.03 | 3547 |
| **NCDS** | Psych distress | 1 | 1 living alone | shielding status at 1 | shielding | -0.05 | 0.21 | -0.47 | 0.37 | 3545 |
| **NCDS** | Psych distress | 1 | 1 living alone | sex | female | 0.01 | 0.09 | -0.18 | 0.19 | 3567 |
| **NCDS** | Psych distress | 1 | 1 living alone | Prior mental ill-health | yes | 0.08 | 0.21 | -0.33 | 0.49 | 3567 |
| **NCDS** | Psych distress | 2 | 1 living alone | loneliness at 1 | lonely | -0.48 | 0.20 | -0.87 | -0.09 | 3581 |
| **NCDS** | Psych distress | 2 | 1 living alone | shielding status at 1 | shielding | 0.03 | 0.17 | -0.30 | 0.37 | 3667 |
| **NCDS** | Psych distress | 2 | 1 living alone | sex | female | -0.07 | 0.08 | -0.24 | 0.09 | 3692 |
| **NCDS** | Psych distress | 2 | 1 living alone | Prior mental ill-health | yes | -0.20 | 0.15 | -0.50 | 0.10 | 3692 |
| **NCDS** | Psych distress | 3 | 1 living alone | loneliness at 1 | lonely | -0.49 | 0.21 | -0.90 | -0.07 | 3414 |
| **NCDS** | Psych distress | 3 | 1 living alone | shielding status at 1 | shielding | 0.00 | 0.18 | -0.36 | 0.36 | 3489 |
| **NCDS** | Psych distress | 3 | 1 living alone | sex | female | -0.03 | 0.08 | -0.18 | 0.13 | 3512 |
| **NCDS** | Psych distress | 3 | 1 living alone | Prior mental ill-health | yes | -0.15 | 0.15 | -0.44 | 0.15 | 3512 |
| **NCDS** | Life satisfaction | 0 | 1 living alone | loneliness at 1 | lonely | 0.29 | 0.19 | -0.09 | 0.66 | 3448 |
| **NCDS** | Life satisfaction | 0 | 1 living alone | shielding status at 1 | shielding | 0.10 | 0.23 | -0.36 | 0.55 | 3527 |
| **NCDS** | Life satisfaction | 0 | 1 living alone | sex | female | 0.01 | 0.11 | -0.20 | 0.23 | 3551 |
| **NCDS** | Life satisfaction | 0 | 1 living alone | Prior mental ill-health | yes | -0.11 | 0.15 | -0.41 | 0.18 | 3551 |
| **NCDS** | Life satisfaction | 1 | 1 living alone | loneliness at 1 | lonely | 0.20 | 0.22 | -0.23 | 0.62 | 3595 |
| **NCDS** | Life satisfaction | 1 | 1 living alone | shielding status at 1 | shielding | 0.14 | 0.22 | -0.29 | 0.57 | 3600 |
| **NCDS** | Life satisfaction | 1 | 1 living alone | sex | female | 0.14 | 0.12 | -0.09 | 0.36 | 3623 |
| **NCDS** | Life satisfaction | 1 | 1 living alone | Prior mental ill-health | yes | -0.05 | 0.21 | -0.46 | 0.37 | 3623 |
| **NCDS** | Life satisfaction | 2 | 1 living alone | loneliness at 1 | lonely | 0.22 | 0.19 | -0.15 | 0.60 | 3585 |
| **NCDS** | Life satisfaction | 2 | 1 living alone | shielding status at 1 | shielding | -0.12 | 0.20 | -0.52 | 0.27 | 3671 |
| **NCDS** | Life satisfaction | 2 | 1 living alone | sex | female | -0.02 | 0.08 | -0.19 | 0.15 | 3696 |
| **NCDS** | Life satisfaction | 2 | 1 living alone | Prior mental ill-health | yes | 0.18 | 0.14 | -0.09 | 0.46 | 3696 |
| **NCDS** | Life satisfaction | 3 | 1 living alone | loneliness at 1 | lonely | 0.10 | 0.20 | -0.29 | 0.49 | 3420 |
| **NCDS** | Life satisfaction | 3 | 1 living alone | shielding status at 1 | shielding | -0.08 | 0.20 | -0.47 | 0.32 | 3496 |

| | | | | | | | | | | |
|---|---|---|---|---|---|---|---|---|---|---|
| **NCDS** | Life satisfaction | 3 | 1 living alone | sex | female | -0.10 | 0.09 | -0.28 | 0.07 | 3520 |
| **NCDS** | Life satisfaction | 3 | 1 living alone | Prior mental ill-health | yes | -0.01 | 0.14 | -0.29 | 0.27 | 3520 |
| **GS** | Psych distress | 0 | Live Alone | 1 loneliness | lonely | 0.06 | 0.19 | -0.19 | 0.31 | 2968 |
| **GS** | Psych distress | 0 | Live Alone | shielding | shielding status at 1 | -0.06 | 0.26 | -0.39 | 0.27 | 2984 |
| **GS** | Psych distress | 0 | Live Alone | sex | female | 0.02 | 0.09 | -0.13 | 0.17 | 2984 |
| **GS** | Psych distress | 0 | Live Alone | prepandemic MH | prior mental ill health | 0.19 | 0.08 | 0.09 | 0.29 | 2984 |
| **GS** | Psych distress | 0 | Live Alone | age group | 35 - 44 | -0.37 | 0.23 | -0.74 | 0.01 | 2984 |
| **GS** | Psych distress | 0 | Live Alone | age group | 45 - 54 | 0.01 | 0.22 | -0.33 | 0.34 | 2984 |
| **GS** | Psych distress | 0 | Live Alone | age group | 54 - 64 | -0.19 | 0.20 | -0.50 | 0.12 | 2984 |
| **GS** | Psych distress | 0 | Live Alone | age group | 65 - 74 | -0.29 | 0.20 | -0.59 | 0.02 | 2984 |
| **GS** | Psych distress | 0 | Live Alone | age group | 75+ | -0.14 | 0.22 | -0.54 | 0.26 | 2984 |
| **GS** | Life satisfaction | 1 | Live Alone | 1 loneliness | lonely | 0.31 | 0.18 | -0.05 | 0.66 | 2968 |
| **GS** | Life satisfaction | 1 | Live Alone | shielding | shielding status at 1 | 0.04 | 0.25 | -0.45 | 0.52 | 2982 |
| **GS** | Life satisfaction | 1 | Live Alone | sex | female | 0.02 | 0.11 | -0.19 | 0.24 | 2982 |
| **GS** | Life satisfaction | 1 | Live Alone | prepandemic MH | prior mental ill health | -0.02 | 0.13 | -0.26 | 0.23 | 2982 |
| **GS** | Life satisfaction | 1 | Live Alone | age group | 35 - 44 | 0.15 | 0.28 | -0.40 | 0.71 | 2982 |
| **GS** | Life satisfaction | 1 | Live Alone | age group | 45 - 54 | -0.20 | 0.27 | -0.69 | 0.29 | 2982 |
| **GS** | Life satisfaction | 1 | Live Alone | age group | 54 - 64 | 0.19 | 0.25 | -0.26 | 0.64 | 2982 |
| **GS** | Life satisfaction | 1 | Live Alone | age group | 65 - 74 | 0.16 | 0.24 | -0.29 | 0.60 | 2982 |
| **GS** | Life satisfaction | 1 | Live Alone | age group | 75+ | 0.55 | 0.30 | -0.04 | 1.14 | 2982 |
| **GS** | Psych distress | 1 | Live Alone | 1 loneliness | lonely | -0.36 | 0.28 | -0.72 | 0.00 | 2871 |
| **GS** | Psych distress | 1 | Live Alone | shielding | shielding status at 1 | 0.10 | 0.31 | -0.40 | 0.60 | 2882 |
| **GS** | Psych distress | 1 | Live Alone | sex | female | -0.16 | 0.13 | -0.39 | 0.06 | 2882 |
| **GS** | Psych distress | 1 | Live Alone | prepandemic MH | prior mental ill health | 0.10 | 0.18 | -0.15 | 0.35 | 2882 |
| **GS** | Psych distress | 1 | Live Alone | age group | 35 - 44 | -0.36 | 0.41 | -0.94 | 0.22 | 2882 |
| **GS** | Psych distress | 1 | Live Alone | age group | 45 - 54 | -0.20 | 0.37 | -0.73 | 0.32 | 2882 |
| **GS** | Psych distress | 1 | Live Alone | age group | 54 - 64 | -0.40 | 0.34 | -0.89 | 0.08 | 2882 |
| **GS** | Psych distress | 1 | Live Alone | age group | 65 - 74 | -0.65 | 0.33 | -1.12 | -0.17 | 2882 |
| **GS** | Psych distress | 1 | Live Alone | age group | 75+ | -0.60 | 0.37 | -1.22 | 0.02 | 2882 |
| **GS** | Life satisfaction | 2 | Live Alone | 1 loneliness | lonely | 0.25 | 0.24 | -0.14 | 0.64 | 2172 |
| **GS** | Life satisfaction | 2 | Live Alone | shielding | shielding status at 1 | -0.06 | 0.26 | -0.55 | 0.43 | 2184 |
| **GS** | Life satisfaction | 2 | Live Alone | sex | female | 0.00 | 0.12 | -0.22 | 0.23 | 2184 |

| | | | | | | | | | | |
|---|---|---|---|---|---|---|---|---|---|---|
| **GS** | Life satisfaction | 2 | Live Alone | prepandemic MH | prior mental ill health | 0.07 | 0.15 | -0.17 | 0.32 | 2184 |
| **GS** | Life satisfaction | 2 | Live Alone | age group | 35 - 44 | 0.10 | 0.37 | -0.55 | 0.75 | 2184 |
| **GS** | Life satisfaction | 2 | Live Alone | age group | 45 - 54 | -0.12 | 0.36 | -0.69 | 0.45 | 2184 |
| **GS** | Life satisfaction | 2 | Live Alone | age group | 54 - 64 | -0.02 | 0.34 | -0.56 | 0.53 | 2184 |
| **GS** | Life satisfaction | 2 | Live Alone | age group | 65 - 74 | 0.13 | 0.33 | -0.41 | 0.66 | 2184 |
| **GS** | Life satisfaction | 2 | Live Alone | age group | 75+ | 0.37 | 0.36 | -0.30 | 1.03 | 2184 |
| **GS** | Psych distress | 2 | Live Alone | 1 loneliness | lonely | -0.71 | 0.33 | -1.15 | -0.26 | 2096 |
| **GS** | Psych distress | 2 | Live Alone | shielding | shielding status at 1 | 0.41 | 0.39 | -0.14 | 0.97 | 2106 |
| **GS** | Psych distress | 2 | Live Alone | sex | female | -0.17 | 0.15 | -0.42 | 0.09 | 2106 |
| **GS** | Psych distress | 2 | Live Alone | prepandemic MH | prior mental ill health | -0.07 | 0.20 | -0.35 | 0.20 | 2106 |
| **GS** | Psych distress | 2 | Live Alone | age group | 35 - 44 | -0.58 | 0.55 | -1.34 | 0.18 | 2106 |
| **GS** | Psych distress | 2 | Live Alone | age group | 45 - 54 | -0.39 | 0.50 | -1.06 | 0.29 | 2106 |
| **GS** | Psych distress | 2 | Live Alone | age group | 54 - 64 | -0.50 | 0.49 | -1.14 | 0.14 | 2106 |
| **GS** | Psych distress | 2 | Live Alone | age group | 65 - 74 | -0.66 | 0.48 | -1.28 | -0.03 | 2106 |
| **GS** | Psych distress | 2 | Live Alone | age group | 75+ | -0.86 | 0.50 | -1.63 | -0.09 | 2106 |
| **GS** | Life satisfaction | 3 | Live Alone | 1 loneliness | lonely | 0.49 | 0.25 | 0.05 | 0.93 | 2013 |
| **GS** | Life satisfaction | 3 | Live Alone | shielding | shielding status at 1 | -0.43 | 0.31 | -1.00 | 0.14 | 2024 |
| **GS** | Life satisfaction | 3 | Live Alone | sex | female | 0.03 | 0.13 | -0.22 | 0.29 | 2024 |
| **GS** | Life satisfaction | 3 | Live Alone | prepandemic MH | prior mental ill health | -0.07 | 0.15 | -0.36 | 0.22 | 2024 |
| **GS** | Life satisfaction | 3 | Live Alone | age group | 35 - 44 | -0.05 | 0.35 | -0.74 | 0.65 | 2024 |
| **GS** | Life satisfaction | 3 | Live Alone | age group | 45 - 54 | -0.31 | 0.34 | -0.91 | 0.29 | 2024 |
| **GS** | Life satisfaction | 3 | Live Alone | age group | 54 - 64 | -0.27 | 0.32 | -0.83 | 0.29 | 2024 |
| **GS** | Life satisfaction | 3 | Live Alone | age group | 65 - 74 | -0.15 | 0.31 | -0.70 | 0.40 | 2024 |
| **GS** | Life satisfaction | 3 | Live Alone | age group | 75+ | -0.01 | 0.35 | -0.74 | 0.72 | 2024 |
| **GS** | Psych distress | 3 | Live Alone | 1 loneliness | lonely | -0.45 | 0.32 | -0.94 | 0.03 | 1923 |
| **GS** | Psych distress | 3 | Live Alone | shielding | shielding status at 1 | 0.30 | 0.47 | -0.35 | 0.95 | 1932 |
| **GS** | Psych distress | 3 | Live Alone | sex | female | -0.27 | 0.16 | -0.56 | 0.02 | 1932 |
| **GS** | Psych distress | 3 | Live Alone | prepandemic MH | prior mental ill health | 0.06 | 0.21 | -0.25 | 0.37 | 1932 |
| **GS** | Psych distress | 3 | Live Alone | age group | 35 - 44 | 0.49 | 0.46 | -0.28 | 1.26 | 1932 |
| **GS** | Psych distress | 3 | Live Alone | age group | 45 - 54 | 0.31 | 0.39 | -0.36 | 0.97 | 1932 |
| **GS** | Psych distress | 3 | Live Alone | age group | 54 - 64 | 0.43 | 0.38 | -0.19 | 1.06 | 1932 |
| **GS** | Psych distress | 3 | Live Alone | age group | 65 - 74 | 0.21 | 0.37 | -0.41 | 0.82 | 1932 |
| **GS** | Psych distress | 3 | Live Alone | age group | 75+ | 0.14 | 0.41 | -0.71 | 0.99 | 1932 |

| | | | | | | | | | | |
|---|---|---|---|---|---|---|---|---|---|---|
| **USOC** | Psych distress | 0 | Live Alone | shielding | shielding status at 1 | 0.21 | 0.13 | -0.05 | 0.46 | 12088 |
| **USOC** | Life satisfaction | 0 | Live Alone | shielding | shielding status at 1 | -0.07 | 0.12 | -0.30 | 0.16 | 12088 |
| **USOC** | Life satisfaction | 1 | Live Alone | shielding | shielding status at 1 | -0.18 | 0.15 | -0.48 | 0.12 | 10132 |
| **USOC** | Psych distress | 0 | Live Alone | 0 loneliness | lonely | -0.20 | 0.09 | -0.37 | -0.03 | 22187 |
| **USOC** | Life satisfaction | 0 | Live Alone | 0 loneliness | lonely | 0.03 | 0.07 | -0.11 | 0.18 | 22187 |
| **USOC** | Life satisfaction | 1 | Live Alone | 0 loneliness | lonely | 0.04 | 0.11 | -0.19 | 0.26 | 10132 |
| **USOC** | Psych distress | 0 | Live Alone | sex | female | -0.04 | 0.04 | -0.13 | 0.04 | 22187 |
| **USOC** | Life satisfaction | 0 | Live Alone | sex | female | 0.04 | 0.04 | -0.04 | 0.12 | 22187 |
| **USOC** | Life satisfaction | 1 | Live Alone | sex | female | 0.09 | 0.08 | -0.07 | 0.26 | 10132 |
| **USOC** | Psych distress | 0 | Live Alone | age group | 16-24 | 0.20 | 0.16 | -0.11 | 0.52 | 22187 |
| **USOC** | Psych distress | 0 | Live Alone | age group | 25-34 | -0.06 | 0.11 | -0.27 | 0.15 | 22187 |
| **USOC** | Psych distress | 0 | Live Alone | age group | 35-44 | -0.05 | 0.11 | -0.27 | 0.17 | 22187 |
| **USOC** | Psych distress | 0 | Live Alone | age group | 55-64 | 0.09 | 0.08 | -0.07 | 0.25 | 22187 |
| **USOC** | Psych distress | 0 | Live Alone | age group | 65-74 | -0.08 | 0.07 | -0.22 | 0.06 | 22187 |
| **USOC** | Psych distress | 0 | Live Alone | age group | 75+ | -0.18 | 0.07 | -0.32 | -0.04 | 22187 |
| **USOC** | Life satisfaction | 0 | Live Alone | age group | 16-24 | -0.03 | 0.14 | -0.31 | 0.25 | 22187 |
| **USOC** | Life satisfaction | 0 | Live Alone | age group | 25-34 | 0.09 | 0.09 | -0.09 | 0.27 | 22187 |
| **USOC** | Life satisfaction | 0 | Live Alone | age group | 35-44 | -0.03 | 0.11 | -0.24 | 0.18 | 22187 |
| **USOC** | Life satisfaction | 0 | Live Alone | age group | 55-64 | -0.05 | 0.08 | -0.21 | 0.11 | 22187 |
| **USOC** | Life satisfaction | 0 | Live Alone | age group | 65-74 | 0.05 | 0.07 | -0.09 | 0.19 | 22187 |
| **USOC** | Life satisfaction | 0 | Live Alone | age group | 75+ | 0.20 | 0.08 | 0.05 | 0.36 | 22187 |
| **USOC** | Life satisfaction | 1 | Live Alone | age group | 16-24 | 0.18 | 0.35 | -0.51 | 0.87 | 10132 |
| **USOC** | Life satisfaction | 1 | Live Alone | age group | 25-34 | -0.40 | 0.20 | -0.80 | -0.01 | 10132 |
| **USOC** | Life satisfaction | 1 | Live Alone | age group | 35-44 | -0.39 | 0.17 | -0.72 | -0.05 | 10132 |
| **USOC** | Life satisfaction | 1 | Live Alone | age group | 55-64 | -0.10 | 0.13 | -0.35 | 0.16 | 10132 |
| **USOC** | Life satisfaction | 1 | Live Alone | age group | 65-74 | -0.14 | 0.13 | -0.38 | 0.11 | 10132 |
| **USOC** | Life satisfaction | 1 | Live Alone | age group | 75+ | 0.09 | 0.15 | -0.20 | 0.38 | 10132 |
| **USOC** | Life satisfaction | 1 | Live Alone | prepandemic MH | prior mental ill-health | -0.14 | 0.10 | -0.33 | 0.05 | 10132 |
| **USOC** | Psych distress | 1 | Live Alone | shielding | shielding status at 1 | 0.10 | 0.12 | -0.13 | 0.32 | 12088 |
| **USOC** | Psych distress | 2 | Live Alone | shielding | shielding status at 1 | 0.12 | 0.13 | -0.13 | 0.37 | 9905 |
| **USOC** | Psych distress | 3 | Live Alone | shielding | shielding status at 1 | 0.02 | 0.12 | -0.22 | 0.26 | 9913 |
| **USOC** | Life satisfaction | 2 | Live Alone | shielding | shielding status at 1 | -0.01 | 0.11 | -0.23 | 0.21 | 9905 |

| | | | | | | | | | | |
|---|---|---|---|---|---|---|---|---|---|---|
| **USOC** | Life satisfaction | 3 | Live Alone | shielding | shielding status at 1 | -0.07 | 0.10 | -0.27 | 0.13 | 9913 |
| **USOC** | Psych distress | 1 | Live Alone | 1 loneliness | lonely | -0.08 | 0.09 | -0.26 | 0.11 | 12088 |
| **USOC** | Psych distress | 2 | Live Alone | 1 loneliness | lonely | 0.01 | 0.11 | -0.21 | 0.24 | 9905 |
| **USOC** | Psych distress | 3 | Live Alone | 1 loneliness | lonely | -0.05 | 0.11 | -0.27 | 0.17 | 9913 |
| **USOC** | Life satisfaction | 2 | Live Alone | 1 loneliness | lonely | -0.08 | 0.09 | -0.26 | 0.09 | 9905 |
| **USOC** | Life satisfaction | 3 | Live Alone | 1 loneliness | lonely | 0.02 | 0.08 | -0.15 | 0.18 | 9913 |
| **USOC** | Psych distress | 1 | Live Alone | sex | female | -0.07 | 0.06 | -0.19 | 0.04 | 12088 |
| **USOC** | Psych distress | 2 | Live Alone | sex | female | -0.09 | 0.06 | -0.21 | 0.03 | 9905 |
| **USOC** | Psych distress | 3 | Live Alone | sex | female | -0.03 | 0.06 | -0.15 | 0.09 | 9913 |
| **USOC** | Life satisfaction | 2 | Live Alone | sex | female | 0.17 | 0.06 | 0.05 | 0.29 | 9905 |
| **USOC** | Life satisfaction | 3 | Live Alone | sex | female | 0.18 | 0.06 | 0.07 | 0.29 | 9913 |
| **USOC** | Psych distress | 1 | Live Alone | age group | 16-24 | -0.49 | 0.37 | -1.21 | 0.24 | 12088 |
| **USOC** | Psych distress | 1 | Live Alone | age group | 25-34 | 0.10 | 0.16 | -0.21 | 0.42 | 12088 |
| **USOC** | Psych distress | 1 | Live Alone | age group | 35-44 | -0.13 | 0.15 | -0.43 | 0.17 | 12088 |
| **USOC** | Psych distress | 1 | Live Alone | age group | 55-64 | -0.07 | 0.10 | -0.26 | 0.13 | 12088 |
| **USOC** | Psych distress | 1 | Live Alone | age group | 65-74 | -0.06 | 0.10 | -0.25 | 0.12 | 12088 |
| **USOC** | Psych distress | 1 | Live Alone | age group | 75+ | -0.16 | 0.10 | -0.36 | 0.05 | 12088 |
| **USOC** | Psych distress | 2 | Live Alone | age group | 16-24 | -0.50 | 0.32 | -1.13 | 0.13 | 9905 |
| **USOC** | Psych distress | 2 | Live Alone | age group | 25-34 | 0.28 | 0.18 | -0.08 | 0.64 | 9905 |
| **USOC** | Psych distress | 2 | Live Alone | age group | 35-44 | 0.03 | 0.17 | -0.31 | 0.37 | 9905 |
| **USOC** | Psych distress | 2 | Live Alone | age group | 55-64 | -0.05 | 0.10 | -0.25 | 0.15 | 9905 |
| **USOC** | Psych distress | 2 | Live Alone | age group | 65-74 | -0.07 | 0.10 | -0.27 | 0.12 | 9905 |
| **USOC** | Psych distress | 2 | Live Alone | age group | 75+ | -0.18 | 0.11 | -0.40 | 0.03 | 9905 |
| **USOC** | Psych distress | 3 | Live Alone | age group | 16-24 | -0.37 | 0.41 | -1.17 | 0.42 | 9913 |
| **USOC** | Psych distress | 3 | Live Alone | age group | 25-34 | 0.00 | 0.17 | -0.34 | 0.34 | 9913 |
| **USOC** | Psych distress | 3 | Live Alone | age group | 35-44 | -0.04 | 0.16 | -0.35 | 0.26 | 9913 |
| **USOC** | Psych distress | 3 | Live Alone | age group | 55-64 | -0.06 | 0.10 | -0.26 | 0.15 | 9913 |
| **USOC** | Psych distress | 3 | Live Alone | age group | 65-74 | -0.04 | 0.10 | -0.24 | 0.16 | 9913 |
| **USOC** | Psych distress | 3 | Live Alone | age group | 75+ | -0.09 | 0.11 | -0.31 | 0.12 | 9913 |
| **USOC** | Life satisfaction | 2 | Live Alone | age group | 16-24 | 0.28 | 0.26 | -0.22 | 0.78 | 9905 |
| **USOC** | Life satisfaction | 2 | Live Alone | age group | 25-34 | -0.17 | 0.13 | -0.43 | 0.09 | 9905 |
| **USOC** | Life satisfaction | 2 | Live Alone | age group | 35-44 | -0.23 | 0.14 | -0.50 | 0.03 | 9905 |
| **USOC** | Life satisfaction | 2 | Live Alone | age group | 55-64 | -0.02 | 0.10 | -0.21 | 0.17 | 9905 |

| | | | | | | | | | | |
|---|---|---|---|---|---|---|---|---|---|---|
| **USOC** | Life satisfaction | 2 | Live Alone | age group | 65-74 | 0.13 | 0.09 | -0.05 | 0.31 | 9905 |
| **USOC** | Life satisfaction | 2 | Live Alone | age group | 75+ | 0.33 | 0.11 | 0.11 | 0.54 | 9905 |
| **USOC** | Life satisfaction | 3 | Live Alone | age group | 16-24 | 0.43 | 0.24 | -0.04 | 0.89 | 9913 |
| **USOC** | Life satisfaction | 3 | Live Alone | age group | 25-34 | 0.00 | 0.13 | -0.25 | 0.24 | 9913 |
| **USOC** | Life satisfaction | 3 | Live Alone | age group | 35-44 | -0.06 | 0.12 | -0.30 | 0.19 | 9913 |
| **USOC** | Life satisfaction | 3 | Live Alone | age group | 55-64 | -0.05 | 0.09 | -0.22 | 0.13 | 9913 |
| **USOC** | Life satisfaction | 3 | Live Alone | age group | 65-74 | 0.05 | 0.09 | -0.12 | 0.23 | 9913 |
| **USOC** | Life satisfaction | 3 | Live Alone | age group | 75+ | 0.06 | 0.10 | -0.14 | 0.26 | 9913 |
| **USOC** | Psych distress | 1 | Live Alone | prepandemic MH | prior mental ill-health | 0.14 | 0.08 | -0.02 | 0.30 | 12088 |
| **USOC** | Psych distress | 2 | Live Alone | prepandemic MH | prior mental ill-health | 0.22 | 0.09 | 0.05 | 0.40 | 9905 |
| **USOC** | Psych distress | 3 | Live Alone | prepandemic MH | prior mental ill-health | 0.13 | 0.09 | -0.05 | 0.30 | 9913 |
| **USOC** | Life satisfaction | 2 | Live Alone | prepandemic MH | prior mental ill-health | -0.26 | 0.07 | -0.40 | -0.11 | 9905 |
| **USOC** | Life satisfaction | 3 | Live Alone | prepandemic MH | prior mental ill-health | -0.19 | 0.07 | -0.32 | -0.05 | 9913 |
| **BCS** | Psych distress | 0 | 0 living alone | loneliness at 0 | lonely | -0.34 | 0.26 | -0.84 | 0.16 | 2405 |
| **BCS** | Psych distress | 0 | 0 living alone | shielding status at 1 | shielding | 0.09 | 0.31 | -0.51 | 0.69 | 2455 |
| **BCS** | Psych distress | 0 | 0 living alone | sex | female | 0.22 | 0.14 | -0.06 | 0.49 | 2461 |
| **BCS** | Psych distress | 0 | 0 living alone | Prior mental ill-health | yes | -0.02 | 0.12 | -0.25 | 0.21 | 2461 |
| **BCS** | Psych distress | 1 | 1 living alone | loneliness at 1 | lonely | -0.43 | 0.20 | -0.83 | -0.03 | 2608 |
| **BCS** | Psych distress | 1 | 1 living alone | shielding status at 1 | shielding | -0.10 | 0.36 | -0.81 | 0.61 | 2609 |
| **BCS** | Psych distress | 1 | 1 living alone | sex | female | 0.00 | 0.13 | -0.26 | 0.27 | 2616 |
| **BCS** | Psych distress | 1 | 1 living alone | Prior mental ill-health | yes | -0.12 | 0.18 | -0.47 | 0.24 | 2616 |
| **BCS** | Psych distress | 2 | 1 living alone | loneliness at 1 | lonely | -0.42 | 0.22 | -0.85 | 0.02 | 2611 |
| **BCS** | Psych distress | 2 | 1 living alone | shielding status at 1 | shielding | 0.52 | 0.24 | 0.05 | 0.99 | 2666 |
| **BCS** | Psych distress | 2 | 1 living alone | sex | female | 0.00 | 0.13 | -0.25 | 0.25 | 2673 |
| **BCS** | Psych distress | 2 | 1 living alone | Prior mental ill-health | yes | 0.28 | 0.15 | -0.03 | 0.58 | 2673 |
| **BCS** | Psych distress | 3 | 1 living alone | loneliness at 1 | lonely | -0.17 | 0.22 | -0.60 | 0.26 | 2356 |
| **BCS** | Psych distress | 3 | 1 living alone | shielding status at 1 | shielding | 0.32 | 0.28 | -0.23 | 0.88 | 2401 |
| **BCS** | Psych distress | 3 | 1 living alone | sex | female | 0.00 | 0.13 | -0.24 | 0.25 | 2406 |
| **BCS** | Psych distress | 3 | 1 living alone | Prior mental ill-health | yes | 0.20 | 0.17 | -0.13 | 0.52 | 2406 |
| **BCS** | Life satisfaction | 0 | 1 living alone | loneliness at 1 | lonely | -0.37 | 0.23 | -0.83 | 0.08 | 2401 |
| **BCS** | Life satisfaction | 0 | 1 living alone | shielding status at 1 | shielding | 0.31 | 0.31 | -0.29 | 0.92 | 2451 |
| **BCS** | Life satisfaction | 0 | 1 living alone | sex | female | 0.05 | 0.13 | -0.22 | 0.31 | 2457 |

| | | | | | | | | | | |
|---|---|---|---|---|---|---|---|---|---|---|
| **BCS** | Life satisfaction | 0 | 1 living alone | Prior mental ill-health | yes | -0.25 | 0.19 | -0.62 | 0.12 | 2457 |
| **BCS** | Life satisfaction | 1 | 1 living alone | loneliness at 1 | lonely | -0.06 | 0.24 | -0.53 | 0.40 | 2628 |
| **BCS** | Life satisfaction | 1 | 1 living alone | shielding status at 1 | shielding | 0.03 | 0.44 | -0.83 | 0.88 | 2631 |
| **BCS** | Life satisfaction | 1 | 1 living alone | sex | female | 0.01 | 0.16 | -0.31 | 0.33 | 2638 |
| **BCS** | Life satisfaction | 1 | 1 living alone | Prior mental ill-health | yes | -0.08 | 0.21 | -0.49 | 0.32 | 2638 |
| **BCS** | Life satisfaction | 2 | 1 living alone | loneliness at 1 | lonely | -0.15 | 0.20 | -0.54 | 0.25 | 2616 |
| **BCS** | Life satisfaction | 2 | 1 living alone | shielding status at 1 | shielding | -0.29 | 0.26 | -0.80 | 0.23 | 2671 |
| **BCS** | Life satisfaction | 2 | 1 living alone | sex | female | 0.18 | 0.12 | -0.05 | 0.42 | 2678 |
| **BCS** | Life satisfaction | 2 | 1 living alone | Prior mental ill-health | yes | -0.05 | 0.16 | -0.36 | 0.26 | 2678 |
| **BCS** | Life satisfaction | 3 | 1 living alone | loneliness at 1 | lonely | -0.06 | 0.22 | -0.50 | 0.38 | 2362 |
| **BCS** | Life satisfaction | 3 | 1 living alone | shielding status at 1 | shielding | -0.11 | 0.28 | -0.66 | 0.45 | 2407 |
| **BCS** | Life satisfaction | 3 | 1 living alone | sex | female | 0.15 | 0.13 | -0.10 | 0.41 | 2412 |
| **BCS** | Life satisfaction | 3 | 1 living alone | Prior mental ill-health | yes | 0.05 | 0.18 | -0.29 | 0.40 | 2412 |
| **NS** | Psych distress | 0 | 0 living alone | loneliness at 0 | lonely | 0.05 | 0.32 | -0.58 | 0.68 | 1080 |
| **NS** | Psych distress | 0 | 0 living alone | shielding status at 1 | shielding | -0.47 | 0.36 | -1.18 | 0.24 | 1110 |
| **NS** | Psych distress | 0 | 0 living alone | sex | female | -0.40 | 0.15 | -0.70 | -0.10 | 1114 |
| **NS** | Psych distress | 0 | 0 living alone | Prior mental ill-health | yes | 0.17 | 0.14 | -0.11 | 0.44 | 1114 |
| **NS** | Psych distress | 1 | 1 living alone | loneliness at 1 | lonely | -0.15 | 0.36 | -0.86 | 0.55 | 1135 |
| **NS** | Psych distress | 1 | 1 living alone | shielding status at 1 | shielding | 0.13 | 0.42 | -0.70 | 0.97 | 1131 |
| **NS** | Psych distress | 1 | 1 living alone | sex | female | 0.09 | 0.22 | -0.35 | 0.52 | 1135 |
| **NS** | Psych distress | 1 | 1 living alone | Prior mental ill-health | yes | -0.03 | 0.28 | -0.58 | 0.52 | 1135 |
| **NS** | Psych distress | 2 | 1 living alone | loneliness at 1 | lonely | -0.61 | 0.28 | -1.16 | -0.06 | 1141 |
| **NS** | Psych distress | 2 | 1 living alone | shielding status at 1 | shielding | -0.27 | 0.51 | -1.27 | 0.73 | 1178 |
| **NS** | Psych distress | 2 | 1 living alone | sex | female | -0.14 | 0.18 | -0.49 | 0.22 | 1182 |
| **NS** | Psych distress | 2 | 1 living alone | Prior mental ill-health | yes | 0.03 | 0.23 | -0.42 | 0.48 | 1182 |
| **NS** | Psych distress | 3 | 1 living alone | loneliness at 1 | lonely | -0.64 | 0.28 | -1.18 | -0.10 | 1071 |
| **NS** | Psych distress | 3 | 1 living alone | shielding status at 1 | shielding | -0.15 | 0.58 | -1.30 | 0.99 | 1099 |
| **NS** | Psych distress | 3 | 1 living alone | sex | female | -0.26 | 0.18 | -0.61 | 0.09 | 1103 |
| **NS** | Psych distress | 3 | 1 living alone | Prior mental ill-health | yes | -0.05 | 0.23 | -0.51 | 0.41 | 1103 |
| **NS** | Life satisfaction | 0 | 1 living alone | loneliness at 1 | lonely | 0.00 | 0.08 | -0.17 | 0.16 | 1080 |
| **NS** | Life satisfaction | 0 | 1 living alone | shielding status at 1 | shielding | -0.25 | 0.07 | -0.40 | -0.11 | 1110 |
| **NS** | Life satisfaction | 0 | 1 living alone | sex | female | -0.07 | 0.04 | -0.15 | 0.01 | 1114 |

| | | | | | | | | | | |
|---|---|---|---|---|---|---|---|---|---|---|
| **NS** | Life satisfaction | 0 | 1 living alone | Prior mental ill-health | yes | 0.04 | 0.05 | -0.06 | 0.14 | 1114 |
| **NS** | Life satisfaction | 1 | 1 living alone | loneliness at 1 | lonely | -0.17 | 0.26 | -0.68 | 0.35 | 1157 |
| **NS** | Life satisfaction | 1 | 1 living alone | shielding status at 1 | shielding | 0.10 | 0.29 | -0.47 | 0.68 | 1154 |
| **NS** | Life satisfaction | 1 | 1 living alone | sex | female | 0.04 | 0.20 | -0.35 | 0.44 | 1158 |
| **NS** | Life satisfaction | 1 | 1 living alone | Prior mental ill-health | yes | 0.31 | 0.24 | -0.16 | 0.79 | 1158 |
| **NS** | Life satisfaction | 2 | 1 living alone | loneliness at 1 | lonely | 0.15 | 0.17 | -0.19 | 0.48 | 1145 |
| **NS** | Life satisfaction | 2 | 1 living alone | shielding status at 1 | shielding | -0.01 | 0.29 | -0.58 | 0.56 | 1183 |
| **NS** | Life satisfaction | 2 | 1 living alone | sex | female | -0.03 | 0.11 | -0.25 | 0.18 | 1187 |
| **NS** | Life satisfaction | 2 | 1 living alone | Prior mental ill-health | yes | 0.01 | 0.15 | -0.29 | 0.31 | 1187 |
| **NS** | Life satisfaction | 3 | 1 living alone | loneliness at 1 | lonely | 0.29 | 0.18 | -0.06 | 0.64 | 1074 |
| **NS** | Life satisfaction | 3 | 1 living alone | shielding status at 1 | shielding | 0.22 | 0.28 | -0.33 | 0.76 | 1102 |
| **NS** | Life satisfaction | 3 | 1 living alone | sex | female | 0.25 | 0.12 | 0.01 | 0.50 | 1106 |
| **NS** | Life satisfaction | 3 | 1 living alone | Prior mental ill-health | yes | -0.07 | 0.16 | -0.38 | 0.24 | 1106 |
| **ELSA** | Psych distress | 0 | 0 living alone | shielding status at 1 | shielding | -0.09 | 0.12 | -0.32 | 0.15 | 5463 |
| **ELSA** | Psych distress | 0 | 0 living alone | lionliness 0 | often lonely | -0.26 | 0.20 | -0.66 | 0.14 | 5117 |
| **ELSA** | Psych distress | 0 | 0 living alone | sex | female | 0.16 | 0.09 | -0.03 | 0.34 | 5470 |
| **ELSA** | Psych distress | 0 | 0 living alone | age group | 55-64 | -0.05 | 0.20 | -0.43 | 0.34 | 5470 |
| **ELSA** | Psych distress | 0 | 0 living alone | age group | 65-74 | -0.15 | 0.17 | -0.49 | 0.20 | 5470 |
| **ELSA** | Psych distress | 0 | 0 living alone | age group | 75+ | -0.13 | 0.18 | -0.48 | 0.23 | 5470 |
| **ELSA** | Psych distress | 1 | 1 living alone | shielding status at 1 | shielding | -0.08 | 0.10 | -0.29 | 0.12 | 5463 |
| **ELSA** | Psych distress | 1 | 1 living alone | lionliness 1 | often lonely | -0.14 | 0.13 | -0.39 | 0.11 | 5465 |
| **ELSA** | Psych distress | 1 | 1 living alone | sex | female | 0.05 | 0.09 | -0.13 | 0.23 | 5470 |
| **ELSA** | Psych distress | 1 | 1 living alone | age group | 55-64 | -0.17 | 0.20 | -0.57 | 0.22 | 5470 |
| **ELSA** | Psych distress | 1 | 1 living alone | age group | 65-74 | -0.16 | 0.18 | -0.51 | 0.19 | 5470 |
| **ELSA** | Psych distress | 1 | 1 living alone | age group | 75+ | -0.32 | 0.18 | -0.67 | 0.03 | 5470 |
| **ELSA** | Psych distress | 1 | 1 living alone | prior mental ill-health | depressed at 0 | -0.05 | 0.12 | -0.29 | 0.19 | 5470 |
| **ELSA** | Psych distress | 3 | 1 living alone | shielding status at 1 | shielding | 0.06 | 0.10 | -0.14 | 0.26 | 5040 |
| **ELSA** | Psych distress | 3 | 1 living alone | lionliness 1 | often lonely | 0.12 | 0.13 | -0.14 | 0.38 | 5042 |
| **ELSA** | Psych distress | 3 | 1 living alone | sex | female | -0.08 | 0.09 | -0.25 | 0.09 | 5046 |
| **ELSA** | Psych distress | 3 | 1 living alone | age group | 55-64 | 0.12 | 0.19 | -0.26 | 0.50 | 5046 |
| **ELSA** | Psych distress | 3 | 1 living alone | age group | 65-74 | -0.01 | 0.18 | -0.36 | 0.34 | 5046 |
| **ELSA** | Psych distress | 3 | 1 living alone | age group | 75+ | -0.12 | 0.18 | -0.47 | 0.23 | 5046 |

| | | | | | | | | | | |
|---|---|---|---|---|---|---|---|---|---|---|
| **ELSA** | Psych distress | 3 | 1 living alone | prior mental ill-health | depressed at 0 | -0.06 | 0.11 | -0.27 | 0.16 | 5046 |
| **ELSA** | Life satisfaction | 0 | 0 living alone | shielding status at 1 | shielding | 0.03 | 0.12 | -0.21 | 0.26 | 5085 |
| **ELSA** | Life satisfaction | 0 | 0 living alone | lionliness 0 | often lonely | 0.27 | 0.20 | -0.11 | 0.65 | 5058 |
| **ELSA** | Life satisfaction | 0 | 0 living alone | sex | female | -0.05 | 0.09 | -0.23 | 0.14 | 5090 |
| **ELSA** | Life satisfaction | 0 | 0 living alone | age group | 55-64 | 0.09 | 0.21 | -0.32 | 0.51 | 5090 |
| **ELSA** | Life satisfaction | 0 | 0 living alone | age group | 65-74 | 0.02 | 0.20 | -0.36 | 0.40 | 5090 |
| **ELSA** | Life satisfaction | 0 | 0 living alone | age group | 75+ | 0.12 | 0.20 | -0.27 | 0.51 | 5090 |
| **ELSA** | Life satisfaction | 0 | 1 living alone | prior mental ill-health | depressed at 0 | 0.18 | 0.14 | -0.10 | 0.46 | 5090 |
| **ELSA** | Life satisfaction | 1 | 1 living alone | shielding status at 1 | shielding | 0.01 | 0.11 | -0.20 | 0.23 | 5446 |
| **ELSA** | Life satisfaction | 1 | 1 living alone | lionliness 1 | often lonely | 0.22 | 0.16 | -0.10 | 0.54 | 5448 |
| **ELSA** | Life satisfaction | 1 | 1 living alone | sex | female | 0.04 | 0.08 | -0.13 | 0.20 | 5453 |
| **ELSA** | Life satisfaction | 1 | 1 living alone | age group | 55-64 | 0.02 | 0.21 | -0.38 | 0.43 | 5453 |
| **ELSA** | Life satisfaction | 1 | 1 living alone | age group | 65-74 | -0.09 | 0.19 | -0.47 | 0.28 | 5453 |
| **ELSA** | Life satisfaction | 1 | 1 living alone | age group | 75+ | 0.19 | 0.19 | -0.19 | 0.57 | 5453 |
| **ELSA** | Life satisfaction | 1 | 1 living alone | prior mental ill-health | depressed at 0 | 0.18 | 0.12 | -0.06 | 0.41 | 5453 |
| **ELSA** | Life satisfaction | 3 | 1 living alone | shielding status at 1 | shielding | -0.03 | 0.11 | -0.25 | 0.18 | 5031 |
| **ELSA** | Life satisfaction | 3 | 1 living alone | lionliness 1 | often lonely | -0.01 | 0.17 | -0.35 | 0.33 | 5034 |
| **ELSA** | Life satisfaction | 3 | 1 living alone | sex | female | 0.01 | 0.09 | -0.17 | 0.20 | 5038 |
| **ELSA** | Life satisfaction | 3 | 1 living alone | age group | 55-64 | -0.16 | 0.21 | -0.58 | 0.25 | 5038 |
| **ELSA** | Life satisfaction | 3 | 1 living alone | age group | 65-74 | -0.12 | 0.19 | -0.50 | 0.25 | 5038 |
| **ELSA** | Life satisfaction | 3 | 1 living alone | age group | 75+ | 0.01 | 0.19 | -0.37 | 0.39 | 5038 |
| **ELSA** | Life satisfaction | 3 | 1 living alone | prior mental ill-health | depressed at 0 | 0.18 | 0.14 | -0.09 | 0.45 | 5038 |
| **TwinsUK** | Psych distress | 0 | Live Alone | 1 loneliness | lonely | 1.12 | 0.14 | 0.93 | 1.31 | 2325 |
| **TwinsUK** | Psych distress | 0 | Live Alone | shielding | shielding status at 1 | 0.20 | 0.05 | 0.11 | 0.30 | 2326 |
| **TwinsUK** | Psych distress | 0 | Live Alone | sex | female | 0.25 | 0.07 | 0.09 | 0.40 | 2327 |
| **TwinsUK** | Psych distress | 0 | Live Alone | prepandemic MH | prior mental ill health | 2.48 | 0.09 | 2.32 | 2.63 | 2327 |
| **TwinsUK** | Psych distress | 0 | Live Alone | age group | 16-24 | -0.36 | 0.53 | -1.20 | 0.49 | 2327 |
| **TwinsUK** | Psych distress | 0 | Live Alone | age group | 25-34 | 0.15 | 0.24 | -0.29 | 0.60 | 2327 |
| **TwinsUK** | Psych distress | 0 | Live Alone | age group | 35-44 | -0.01 | 0.15 | -0.27 | 0.25 | 2327 |
| **TwinsUK** | Psych distress | 0 | Live Alone | age group | 55-64 | -0.10 | 0.11 | -0.32 | 0.11 | 2327 |
| **TwinsUK** | Psych distress | 0 | Live Alone | age group | 65-74 | -0.18 | 0.15 | -0.49 | 0.12 | 2327 |
| **TwinsUK** | Psych distress | 0 | Live Alone | age group | 75+ | -0.08 | 0.21 | -0.52 | 0.35 | 2327 |

| | | | | | | | | | | |
|---|---|---|---|---|---|---|---|---|---|---|
| **TwinsUK** | Psych distress | 1 | Live Alone | 1 loneliness | lonely | 1.65 | 0.13 | 1.45 | 1.86 | 2321 |
| **TwinsUK** | Psych distress | 1 | Live Alone | shielding | shielding status at 1 | 0.23 | 0.06 | 0.12 | 0.33 | 2322 |
| **TwinsUK** | Psych distress | 1 | Live Alone | sex | female | 0.33 | 0.08 | 0.15 | 0.50 | 2323 |
| **TwinsUK** | Psych distress | 1 | Live Alone | prepandemic MH | prior mental ill health | 1.34 | 0.13 | 1.14 | 1.55 | 2323 |
| **TwinsUK** | Psych distress | 1 | Live Alone | age group | 16-24 | -0.16 | 0.50 | -1.11 | 0.78 | 2323 |
| **TwinsUK** | Psych distress | 1 | Live Alone | age group | 25-34 | 0.41 | 0.23 | -0.09 | 0.90 | 2323 |
| **TwinsUK** | Psych distress | 1 | Live Alone | age group | 35-44 | 0.32 | 0.15 | 0.02 | 0.61 | 2323 |
| **TwinsUK** | Psych distress | 1 | Live Alone | age group | 55-64 | 0.01 | 0.12 | -0.22 | 0.25 | 2323 |
| **TwinsUK** | Psych distress | 1 | Live Alone | age group | 65-74 | -0.17 | 0.17 | -0.52 | 0.17 | 2323 |
| **TwinsUK** | Psych distress | 1 | Live Alone | age group | 75+ | -0.21 | 0.24 | -0.70 | 0.27 | 2323 |
| **TwinsUK** | Psych distress | 2 | Live Alone | 1 loneliness | lonely | 1.22 | 0.15 | 1.03 | 1.42 | 2321 |
| **TwinsUK** | Psych distress | 2 | Live Alone | shielding | shielding status at 1 | 0.22 | 0.05 | 0.12 | 0.31 | 2019 |
| **TwinsUK** | Psych distress | 2 | Live Alone | sex | female | 0.35 | 0.07 | 0.19 | 0.51 | 2020 |
| **TwinsUK** | Psych distress | 2 | Live Alone | prepandemic MH | prior mental ill health | 1.33 | 0.12 | 1.15 | 1.51 | 2020 |
| **TwinsUK** | Psych distress | 2 | Live Alone | age group | 16-24 | 0.64 | 0.66 | -0.30 | 1.58 | 2020 |
| **TwinsUK** | Psych distress | 2 | Live Alone | age group | 25-34 | 0.62 | 0.21 | 0.17 | 1.06 | 2020 |
| **TwinsUK** | Psych distress | 2 | Live Alone | age group | 35-44 | 0.31 | 0.14 | 0.05 | 0.58 | 2020 |
| **TwinsUK** | Psych distress | 2 | Live Alone | age group | 55-64 | 0.04 | 0.11 | -0.18 | 0.25 | 2020 |
| **TwinsUK** | Psych distress | 2 | Live Alone | age group | 65-74 | -0.07 | 0.16 | -0.38 | 0.24 | 2020 |
| **TwinsUK** | Psych distress | 2 | Live Alone | age group | 75+ | -0.09 | 0.22 | -0.53 | 0.35 | 2020 |
| **TwinsUK** | Psych distress | 3 | Live Alone | 1 loneliness | lonely | 1.12 | 0.12 | 0.93 | 1.31 | 2139 |
| **TwinsUK** | Psych distress | 3 | Live Alone | shielding | shielding status at 1 | 0.22 | 0.05 | 0.12 | 0.31 | 2140 |
| **TwinsUK** | Psych distress | 3 | Live Alone | sex | female | 0.34 | 0.07 | 0.18 | 0.50 | 2141 |
| **TwinsUK** | Psych distress | 3 | Live Alone | prepandemic MH | prior mental ill health | 1.34 | 0.12 | 1.16 | 1.53 | 2141 |
| **TwinsUK** | Psych distress | 3 | Live Alone | age group 45-54(ref) | 16-24 | -0.05 | 0.76 | -1.01 | 0.91 | 2141 |
| **TwinsUK** | Psych distress | 3 | Live Alone | age group | 25-34 | 0.47 | 0.24 | 0.01 | 0.93 | 2141 |
| **TwinsUK** | Psych distress | 3 | Live Alone | age group | 35-44 | 0.13 | 0.14 | -0.13 | 0.40 | 2141 |
| **TwinsUK** | Psych distress | 3 | Live Alone | age group | 55-64 | -0.23 | 0.12 | -0.45 | -0.02 | 2141 |
| **TwinsUK** | Psych distress | 3 | Live Alone | age group | 65-74 | -0.42 | 0.16 | -0.73 | -0.10 | 2141 |
| **TwinsUK** | Psych distress | 3 | Live Alone | age group | 75+ | -0.45 | 0.22 | -0.89 | -0.01 | 2141 |
| **TwinsUK** | Life satisfaction | 1 | Live Alone | 1 loneliness | lonely | -1.21 | 0.12 | -1.41 | -1.00 | 2325 |
| **TwinsUK** | Life satisfaction | 1 | Live Alone | shielding | shielding status at 1 | -0.05 | 0.05 | -0.15 | 0.05 | 2327 |
| **TwinsUK** | Life satisfaction | 1 | Live Alone | sex | female | -0.11 | 0.07 | -0.27 | 0.06 | 2327 |

| | | | | | | | | | | |
|---|---|---|---|---|---|---|---|---|---|---|
| **TwinsUK** | Life satisfaction | 1 | Live Alone | prepandemic MH | prior mental ill health | -0.61 | 0.12 | -0.82 | -0.41 | 2327 |
| **TwinsUK** | Life satisfaction | 1 | Live Alone | age group | 16-24 | -0.23 | 0.37 | -1.13 | 0.66 | 2327 |
| **TwinsUK** | Life satisfaction | 1 | Live Alone | age group | 25-34 | -0.42 | 0.23 | -0.88 | 0.05 | 2327 |
| **TwinsUK** | Life satisfaction | 1 | Live Alone | age group | 35-44 | -0.29 | 0.15 | -0.57 | -0.02 | 2327 |
| **TwinsUK** | Life satisfaction | 1 | Live Alone | age group | 55-64 | -0.01 | 0.11 | -0.23 | 0.21 | 2327 |
| **TwinsUK** | Life satisfaction | 1 | Live Alone | age group | 65-74 | 0.11 | 0.17 | -0.21 | 0.43 | 2327 |
| **TwinsUK** | Life satisfaction | 1 | Live Alone | age group | 75+ | 0.20 | 0.23 | -0.25 | 0.66 | 2327 |
| **TwinsUK** | Life satisfaction | 2 | Live Alone | 1 loneliness | lonely | -1.16 | 0.14 | -1.38 | -0.94 | 2024 |
| **TwinsUK** | Life satisfaction | 2 | Live Alone | shielding | shielding status at 1 | -0.16 | 0.06 | -0.26 | -0.05 | 2025 |
| **TwinsUK** | Life satisfaction | 2 | Live Alone | sex | female | -0.26 | 0.08 | -0.43 | -0.09 | 2026 |
| **TwinsUK** | Life satisfaction | 2 | Live Alone | prepandemic MH | prior mental ill health | -0.76 | 0.13 | -0.97 | -0.55 | 2026 |
| **TwinsUK** | Life satisfaction | 2 | Live Alone | age group | 16-24 | -1.03 | 0.64 | -2.04 | -0.01 | 2026 |
| **TwinsUK** | Life satisfaction | 2 | Live Alone | age group | 25-34 | -0.62 | 0.23 | -1.10 | -0.14 | 2026 |
| **TwinsUK** | Life satisfaction | 2 | Live Alone | age group | 35-44 | -0.27 | 0.15 | -0.55 | 0.02 | 2026 |
| **TwinsUK** | Life satisfaction | 2 | Live Alone | age group | 55-64 | -0.02 | 0.12 | -0.26 | 0.21 | 2026 |
| **TwinsUK** | Life satisfaction | 2 | Live Alone | age group | 65-74 | 0.02 | 0.17 | -0.31 | 0.36 | 2026 |
| **TwinsUK** | Life satisfaction | 2 | Live Alone | age group | 75+ | 0.03 | 0.25 | -0.45 | 0.51 | 2026 |
| **TwinsUK** | Life satisfaction | 3 | Live Alone | 1 loneliness | lonely | -0.99 | 0.13 | -1.18 | -0.79 | 2153 |
| **TwinsUK** | Life satisfaction | 3 | Live Alone | shielding | shielding status at 1 | -0.14 | 0.05 | -0.24 | -0.04 | 2154 |
| **TwinsUK** | Life satisfaction | 3 | Live Alone | sex | female | -0.13 | 0.07 | -0.28 | 0.03 | 2155 |
| **TwinsUK** | Life satisfaction | 3 | Live Alone | prepandemic MH | prior mental ill health | -0.61 | 0.13 | -0.81 | -0.42 | 2155 |
| **TwinsUK** | Life satisfaction | 3 | Live Alone | age group 45-54(ref) | 16-24 | 0.05 | 0.47 | -0.92 | 1.01 | 2155 |
| **TwinsUK** | Life satisfaction | 3 | Live Alone | age group | 25-34 | -0.43 | 0.23 | -0.89 | 0.03 | 2155 |
| **TwinsUK** | Life satisfaction | 3 | Live Alone | age group | 35-44 | -0.16 | 0.14 | -0.43 | 0.10 | 2155 |
| **TwinsUK** | Life satisfaction | 3 | Live Alone | age group | 55-64 | 0.04 | 0.11 | -0.18 | 0.25 | 2155 |
| **TwinsUK** | Life satisfaction | 3 | Live Alone | age group | 65-74 | 0.14 | 0.16 | -0.17 | 0.45 | 2155 |
| **TwinsUK** | Life satisfaction | 3 | Live Alone | age group | 75+ | 0.07 | 0.22 | -0.36 | 0.51 | 2155 |
| **ALSPAC-G1** | Psych distress | 0 | Live Alone | sex | female | 0.23 | 0.19 | -0.15 | 0.61 | 1809 |
| **ALSPAC-G1** | Psych distress | 0 | Live Alone | prepandemic MH | prior mental ill health | -0.20 | 0.11 | -0.41 | 0.01 | 1809 |
| **ALSPAC-G1** | Psych distress | 1 | Live Alone | sex | female | 0.09 | 0.19 | -0.27 | 0.46 | 2064 |
| **ALSPAC-G1** | Psych distress | 1 | Live Alone | prepandemic MH | prior mental ill health | 0.11 | 0.18 | -0.23 | 0.46 | 1774 |
| **ALSPAC-G1** | Psych distress | 2 | Live Alone | sex | female | 0.19 | 0.20 | -0.21 | 0.59 | 1610 |

| | | | | | | | | | | |
|---|---|---|---|---|---|---|---|---|---|---|
| **ALSPAC-G1** | Psych distress | 2 | Live Alone | prepandemic MH | prior mental ill health | 0.11 | 0.20 | -0.27 | 0.50 | 1413 |
| **ALSPAC-G1** | Psych distress | 3 | Live Alone | sex | female | 0.14 | 0.19 | -0.24 | 0.52 | 1919 |
| **ALSPAC-G1** | Psych distress | 3 | Live Alone | prepandemic MH | prior mental ill health | 0.29 | 0.18 | -0.06 | 0.65 | 1675 |
| **ALSPAC-G1** | Life satisfaction | 0 | Live Alone | sex | female | 0.07 | 0.20 | -0.32 | 0.46 | 1720 |
| **ALSPAC-G1** | Life satisfaction | 0 | Live Alone | prepandemic MH | prior mental ill health | 0.23 | 0.16 | -0.08 | 0.54 | 1720 |
| **ALSPAC-G1** | Life satisfaction | 1 | Live Alone | sex | female | 0.05 | 0.19 | -0.31 | 0.42 | 2075 |
| **ALSPAC-G1** | Life satisfaction | 1 | Live Alone | prepandemic MH | prior mental ill health | 0.37 | 0.21 | -0.05 | 0.79 | 1696 |
| **ALSPAC-G1** | Life satisfaction | 2 | Live Alone | sex | female | -0.20 | 0.21 | -0.60 | 0.21 | 1616 |
| **ALSPAC-G1** | Life satisfaction | 2 | Live Alone | prepandemic MH | prior mental ill health | 0.39 | 0.24 | -0.07 | 0.86 | 1351 |
| **ALSPAC-G1** | Life satisfaction | 3 | Live Alone | sex | female | -0.11 | 0.20 | -0.49 | 0.27 | 1920 |
| **ALSPAC-G1** | Life satisfaction | 3 | Live Alone | prepandemic MH | prior mental ill health | 0.13 | 0.22 | -0.29 | 0.56 | 1600 |
| **ALSPAC-G0** | Psych distress | 0 | Live Alone | sex | female | 0.21 | 0.17 | -0.12 | 0.54 | 2702 |
| **ALSPAC-G0** | Psych distress | 0 | Live Alone | prepandemic MH | prior mental ill health | 0.32 | 0.11 | 0.11 | 0.52 | 2702 |
| **ALSPAC-G0** | Psych distress | 1 | Live Alone | sex | female | 0.23 | 0.15 | -0.07 | 0.54 | 3177 |
| **ALSPAC-G0** | Psych distress | 1 | Live Alone | prepandemic MH | prior mental ill health | 0.31 | 0.16 | 0.00 | 0.63 | 2579 |
| **ALSPAC-G0** | Psych distress | 2 | Live Alone | sex | female | 0.10 | 0.16 | -0.22 | 0.42 | 2746 |
| **ALSPAC-G0** | Psych distress | 2 | Live Alone | prepandemic MH | prior mental ill health | 0.48 | 0.17 | 0.14 | 0.82 | 2279 |
| **ALSPAC-G0** | Psych distress | 3 | Live Alone | sex | female | 0.07 | 0.16 | -0.25 | 0.38 | 2808 |
| **ALSPAC-G0** | Psych distress | 3 | Live Alone | prepandemic MH | prior mental ill health | 0.40 | 0.17 | 0.07 | 0.73 | 2349 |
| **ALSPAC-G0** | Psych distress | 0 | Live Alone | age group | 45 - 54 | -0.02 | 0.26 | -0.52 | 0.49 | 2702 |
| **ALSPAC-G0** | Psych distress | 0 | Live Alone | age group | 65 - 74 | -0.61 | 0.19 | -0.99 | -0.24 | 2702 |
| **ALSPAC-G0** | Psych distress | 0 | Live Alone | age group | 75+ | 0.70 | 1.02 | -1.30 | 2.70 | 2702 |
| **ALSPAC-G0** | Psych distress | 1 | Live Alone | age group | 45 - 54 | -0.14 | 0.23 | -0.59 | 0.30 | 3177 |
| **ALSPAC-G0** | Psych distress | 1 | Live Alone | age group | 65 - 74 | -0.08 | 0.18 | -0.44 | 0.27 | 3177 |
| **ALSPAC-G0** | Psych distress | 1 | Live Alone | age group | 75+ | -0.81 | 1.02 | -2.80 | 1.18 | 3177 |
| **ALSPAC-G0** | Psych distress | 2 | Live Alone | age group | 45 - 54 | 0.03 | 0.24 | -0.44 | 0.50 | 2746 |
| **ALSPAC-G0** | Psych distress | 2 | Live Alone | age group | 65 - 74 | -0.11 | 0.19 | -0.48 | 0.26 | 2746 |
| **ALSPAC-G0** | Psych distress | 2 | Live Alone | age group | 75+ | -0.44 | 1.02 | -2.43 | 1.56 | 2746 |
| **ALSPAC-G0** | Psych distress | 3 | Live Alone | age group | 45 - 54 | 0.15 | 0.27 | -0.38 | 0.69 | 2808 |
| **ALSPAC-G0** | Psych distress | 3 | Live Alone | age group | 65 - 74 | 0.02 | 0.17 | -0.31 | 0.35 | 2808 |
| **ALSPAC-G0** | Psych distress | 3 | Live Alone | age group | 75+ | -0.14 | 1.01 | -2.13 | 1.85 | 2808 |

**Note: TP = timepoint; coef = coefficient; se = standard error ; l_ci = lower 95% confidence interval ; u_ci = cupper 95% confidence interval. Models adjust for covariates.**

### Table S6. Regression coefficients from stratified cross-sectional models with continuous outcomes

| cohort | outcome (Z) | TP | stratifier | stratified group | coef | coef_se | lower_ci | upper_ci | n |
|---|---|---|---|---|---|---|---|---|---|
| **NCDS** | Psych distress | T0 | Loneliness at T0 | not lonely | 0.09 | 0.06 | -0.03 | 0.21 | 3290 |
| **NCDS** | Psych distress | T0 | Loneliness at T0 | lonely | -0.16 | 0.25 | -0.65 | 0.32 | 158 |
| **NCDS** | Psych distress | T1 | Loneliness at T1 | not lonely | -0.03 | 0.04 | -0.12 | 0.05 | 3382 |
| **NCDS** | Psych distress | T1 | Loneliness at T1 | lonely | -0.47 | 0.22 | -0.91 | -0.03 | 165 |
| **NCDS** | Psych distress | T2 | Loneliness at T1 | not lonely | -0.03 | 0.04 | -0.11 | 0.05 | 3411 |
| **NCDS** | Psych distress | T2 | Loneliness at T1 | lonely | -0.52 | 0.20 | -0.92 | -0.12 | 170 |
| **NCDS** | Psych distress | T3 | Loneliness at T1 | not lonely | -0.06 | 0.04 | -0.13 | 0.02 | 3260 |
| **NCDS** | Psych distress | T3 | Loneliness at T1 | lonely | -0.55 | 0.21 | -0.97 | -0.12 | 154 |
| **NCDS** | Psych distress | T0 | Sex | male | 0.24 | 0.09 | 0.07 | 0.41 | 1681 |
| **NCDS** | Psych distress | T0 | Sex | female | 0.00 | 0.08 | -0.15 | 0.15 | 1870 |
| **NCDS** | Psych distress | T1 | Sex | male | 0.07 | 0.06 | -0.06 | 0.19 | 1714 |
| **NCDS** | Psych distress | T1 | Sex | female | 0.07 | 0.07 | -0.06 | 0.21 | 1853 |
| **NCDS** | Psych distress | T2 | Sex | male | 0.08 | 0.06 | -0.03 | 0.19 | 1768 |
| **NCDS** | Psych distress | T2 | Sex | female | 0.01 | 0.06 | -0.11 | 0.13 | 1924 |
| **NCDS** | Psych distress | T3 | Sex | male | 0.03 | 0.05 | -0.08 | 0.13 | 1662 |
| **NCDS** | Psych distress | T3 | Sex | female | -0.01 | 0.06 | -0.12 | 0.11 | 1850 |
| **NCDS** | Psych distress | T0 | Shielding status at T1 | not shielding | 0.11 | 0.06 | -0.01 | 0.23 | 3320 |
| **NCDS** | Psych distress | T0 | Shielding status at T1 | shielding | -0.09 | 0.20 | -0.49 | 0.31 | 207 |
| **NCDS** | Psych distress | T1 | Shielding status at T1 | not shielding | 0.07 | 0.05 | -0.03 | 0.16 | 3345 |
| **NCDS** | Psych distress | T1 | Shielding status at T1 | shielding | -0.03 | 0.19 | -0.41 | 0.35 | 200 |
| **NCDS** | Psych distress | T2 | Shielding status at T1 | not shielding | 0.03 | 0.04 | -0.05 | 0.12 | 3456 |
| **NCDS** | Psych distress | T2 | Shielding status at T1 | shielding | 0.07 | 0.18 | -0.28 | 0.41 | 211 |
| **NCDS** | Psych distress | T3 | Shielding status at T1 | not shielding | 0.00 | 0.04 | -0.08 | 0.08 | 3286 |
| **NCDS** | Psych distress | T3 | Shielding status at T1 | shielding | -0.03 | 0.20 | -0.42 | 0.35 | 203 |
| **NCDS** | Psych distress | T0 | Prior mental ill-health | no prior MH | 0.05 | 0.04 | -0.02 | 0.12 | 3175 |
| **NCDS** | Psych distress | T0 | Prior mental ill-health | prior MH | -0.14 | 0.09 | -0.33 | 0.04 | 376 |
| **NCDS** | Psych distress | T1 | Prior mental ill-health | no prior MH | 0.02 | 0.04 | -0.06 | 0.10 | 3197 |
| **NCDS** | Psych distress | T1 | Prior mental ill-health | prior MH | 0.04 | 0.18 | -0.31 | 0.40 | 370 |
| **NCDS** | Psych distress | T2 | Prior mental ill-health | no prior MH | 0.03 | 0.04 | -0.04 | 0.11 | 3299 |
| **NCDS** | Psych distress | T2 | Prior mental ill-health | prior MH | -0.20 | 0.15 | -0.49 | 0.10 | 393 |
| **NCDS** | Psych distress | T3 | Prior mental ill-health | no prior MH | -0.02 | 0.04 | -0.09 | 0.06 | 3141 |

| | | | | | | | | | |
|---|---|---|---|---|---|---|---|---|---|
| **NCDS** | Psych distress | T3 | Prior mental ill-health | prior MH | -0.22 | 0.15 | -0.51 | 0.08 | 371 |
| **NCDS** | Life satisfaction | T0 | Loneliness at T0 | not lonely | -0.49 | 0.06 | -0.61 | -0.38 | 3290 |
| **NCDS** | Life satisfaction | T0 | Loneliness at T0 | lonely | -0.24 | 0.21 | -0.65 | 0.17 | 158 |
| **NCDS** | Life satisfaction | T1 | Loneliness at T1 | not lonely | -0.18 | 0.05 | -0.28 | -0.08 | 3425 |
| **NCDS** | Life satisfaction | T1 | Loneliness at T1 | lonely | 0.01 | 0.20 | -0.39 | 0.41 | 170 |
| **NCDS** | Life satisfaction | T2 | Loneliness at T1 | not lonely | -0.20 | 0.04 | -0.28 | -0.12 | 3415 |
| **NCDS** | Life satisfaction | T2 | Loneliness at T1 | lonely | 0.05 | 0.20 | -0.35 | 0.46 | 170 |
| **NCDS** | Life satisfaction | T3 | Loneliness at T1 | not lonely | -0.17 | 0.05 | -0.26 | -0.09 | 3265 |
| **NCDS** | Life satisfaction | T3 | Loneliness at T1 | lonely | -0.01 | 0.22 | -0.44 | 0.43 | 155 |
| **NCDS** | Life satisfaction | T0 | Sex | male | -0.51 | 0.08 | -0.67 | -0.35 | 1681 |
| **NCDS** | Life satisfaction | T0 | Sex | female | -0.49 | 0.07 | -0.63 | -0.35 | 1870 |
| **NCDS** | Life satisfaction | T1 | Sex | male | -0.35 | 0.08 | -0.50 | -0.19 | 1739 |
| **NCDS** | Life satisfaction | T1 | Sex | female | -0.29 | 0.08 | -0.44 | -0.14 | 1884 |
| **NCDS** | Life satisfaction | T2 | Sex | male | -0.26 | 0.06 | -0.39 | -0.14 | 1770 |
| **NCDS** | Life satisfaction | T2 | Sex | female | -0.33 | 0.06 | -0.44 | -0.22 | 1926 |
| **NCDS** | Life satisfaction | T3 | Sex | male | -0.19 | 0.06 | -0.31 | -0.06 | 1665 |
| **NCDS** | Life satisfaction | T3 | Sex | female | -0.32 | 0.06 | -0.45 | -0.19 | 1855 |
| **NCDS** | Life satisfaction | T0 | Shielding status at T1 | not shielding | -0.50 | 0.06 | -0.61 | -0.39 | 3320 |
| **NCDS** | Life satisfaction | T0 | Shielding status at T1 | shielding | -0.44 | 0.23 | -0.91 | 0.02 | 207 |
| **NCDS** | Life satisfaction | T1 | Shielding status at T1 | not shielding | -0.33 | 0.06 | -0.44 | -0.22 | 3394 |
| **NCDS** | Life satisfaction | T1 | Shielding status at T1 | shielding | -0.21 | 0.21 | -0.62 | 0.20 | 206 |
| **NCDS** | Life satisfaction | T2 | Shielding status at T1 | not shielding | -0.30 | 0.04 | -0.38 | -0.21 | 3459 |
| **NCDS** | Life satisfaction | T2 | Shielding status at T1 | shielding | -0.46 | 0.20 | -0.84 | -0.07 | 212 |
| **NCDS** | Life satisfaction | T3 | Shielding status at T1 | not shielding | -0.25 | 0.05 | -0.35 | -0.16 | 3293 |
| **NCDS** | Life satisfaction | T3 | Shielding status at T1 | shielding | -0.36 | 0.20 | -0.76 | 0.04 | 203 |
| **NCDS** | Life satisfaction | T0 | Prior mental ill-health | no prior MH | -0.46 | 0.06 | -0.57 | -0.35 | 3175 |
| **NCDS** | Life satisfaction | T0 | Prior mental ill-health | prior MH | -0.56 | 0.14 | -0.84 | -0.28 | 376 |
| **NCDS** | Life satisfaction | T1 | Prior mental ill-health | no prior MH | -0.28 | 0.05 | -0.38 | -0.18 | 3239 |
| **NCDS** | Life satisfaction | T1 | Prior mental ill-health | prior MH | -0.38 | 0.17 | -0.71 | -0.04 | 384 |
| **NCDS** | Life satisfaction | T2 | Prior mental ill-health | no prior MH | -0.31 | 0.04 | -0.40 | -0.22 | 3301 |
| **NCDS** | Life satisfaction | T2 | Prior mental ill-health | prior MH | -0.14 | 0.14 | -0.41 | 0.12 | 395 |
| **NCDS** | Life satisfaction | T3 | Prior mental ill-health | no prior MH | -0.24 | 0.05 | -0.34 | -0.15 | 3148 |
| **NCDS** | Life satisfaction | T3 | Prior mental ill-health | prior MH | -0.20 | 0.14 | -0.47 | 0.07 | 372 |

| | | | | | | | | | |
|---|---|---|---|---|---|---|---|---|---|
| **NSHD** | Psych distress | T0 | Loneliness at T0 | not lonely | -0.12 | 0.14 | -0.40 | 0.16 | 1113 |
| **NSHD** | Psych distress | T0 | Loneliness at T0 | lonely | -0.49 | 0.27 | -1.02 | 0.03 | 187 |
| **NSHD** | Psych distress | T1 | Loneliness at T1 | not lonely | 0.34 | 0.17 | 0.01 | 0.66 | 1238 |
| **NSHD** | Psych distress | T1 | Loneliness at T1 | lonely | -0.68 | 0.37 | -1.41 | 0.05 | 202 |
| **NSHD** | Psych distress | T2 | Loneliness at T1 | not lonely | 0.35 | 0.16 | 0.04 | 0.67 | 1242 |
| **NSHD** | Psych distress | T2 | Loneliness at T1 | lonely | -0.53 | 0.31 | -1.14 | 0.08 | 204 |
| **NSHD** | Psych distress | T3 | Loneliness at T1 | not lonely | 0.06 | 0.17 | -0.28 | 0.40 | 864 |
| **NSHD** | Psych distress | T3 | Loneliness at T1 | lonely | -0.61 | 0.34 | -1.28 | 0.07 | 190 |
| **NSHD** | Psych distress | T0 | Sex | male | -0.16 | 0.19 | -0.53 | 0.22 | 590 |
| **NSHD** | Psych distress | T0 | Sex | female | -0.02 | 0.16 | -0.33 | 0.29 | 710 |
| **NSHD** | Psych distress | T1 | Sex | male | -0.03 | 0.23 | -0.48 | 0.43 | 645 |
| **NSHD** | Psych distress | T1 | Sex | female | 0.66 | 0.19 | 0.28 | 1.03 | 795 |
| **NSHD** | Psych distress | T2 | Sex | male | 0.05 | 0.22 | -0.39 | 0.49 | 649 |
| **NSHD** | Psych distress | T2 | Sex | female | 0.46 | 0.17 | 0.12 | 0.80 | 797 |
| **NSHD** | Psych distress | T3 | Sex | male | -0.30 | 0.23 | -0.75 | 0.15 | 493 |
| **NSHD** | Psych distress | T3 | Sex | female | 0.48 | 0.19 | 0.10 | 0.86 | 561 |
| **NSHD** | Psych distress | T0 | Shielding status at T1 | not shielding | -0.08 | 0.13 | -0.33 | 0.17 | 1161 |
| **NSHD** | Psych distress | T0 | Shielding status at T1 | shielding | 0.14 | 0.37 | -0.58 | 0.87 | 139 |
| **NSHD** | Psych distress | T1 | Shielding status at T1 | not shielding | 0.35 | 0.15 | 0.05 | 0.66 | 1285 |
| **NSHD** | Psych distress | T1 | Shielding status at T1 | shielding | 0.94 | 0.44 | 0.07 | 1.82 | 155 |
| **NSHD** | Psych distress | T2 | Shielding status at T1 | not shielding | 0.38 | 0.14 | 0.10 | 0.66 | 1286 |
| **NSHD** | Psych distress | T2 | Shielding status at T1 | shielding | -0.04 | 0.44 | -0.92 | 0.83 | 160 |
| **NSHD** | Psych distress | T3 | Shielding status at T1 | not shielding | 0.21 | 0.15 | -0.10 | 0.51 | 964 |
| **NSHD** | Psych distress | T3 | Shielding status at T1 | shielding | 0.39 | 0.53 | -0.66 | 1.44 | 90 |
| **NSHD** | Psych distress | T0 | Prior mental ill-health | no prior MH | -0.03 | 0.07 | -0.18 | 0.11 | 1083 |
| **NSHD** | Psych distress | T0 | Prior mental ill-health | prior MH | -0.30 | 0.31 | -0.91 | 0.30 | 217 |
| **NSHD** | Psych distress | T1 | Prior mental ill-health | no prior MH | 0.31 | 0.16 | -0.01 | 0.63 | 1074 |
| **NSHD** | Psych distress | T1 | Prior mental ill-health | prior MH | 1.27 | 0.40 | 0.49 | 2.05 | 215 |
| **NSHD** | Psych distress | T2 | Prior mental ill-health | no prior MH | 0.28 | 0.15 | -0.01 | 0.57 | 1074 |
| **NSHD** | Psych distress | T2 | Prior mental ill-health | prior MH | 0.39 | 0.37 | -0.33 | 1.11 | 216 |
| **NSHD** | Psych distress | T3 | Prior mental ill-health | no prior MH | 0.12 | 0.15 | -0.18 | 0.42 | 820 |
| **NSHD** | Psych distress | T3 | Prior mental ill-health | prior MH | 0.29 | 0.43 | -0.57 | 1.15 | 162 |
| **GS** | Life satisfaction | 1 | Prior MH | not shielding | -0.33 | 0.06 | -0.44 | -0.22 | 2457 |

| | | | | | | | | | |
|---|---|---|---|---|---|---|---|---|---|
| **GS** | Life satisfaction | 1 | Prior MH | shielding | -0.37 | 0.13 | -0.62 | -0.13 | 525 |
| **GS** | Life satisfaction | 2 | Prior MH | not shielding | -0.27 | 0.06 | -0.38 | -0.16 | 1811 |
| **GS** | Life satisfaction | 2 | Prior MH | shielding | -0.22 | 0.14 | -0.49 | 0.04 | 373 |
| **GS** | Life satisfaction | 3 | Prior MH | not shielding | -0.26 | 0.07 | -0.39 | -0.14 | 1684 |
| **GS** | Life satisfaction | 3 | Prior MH | shielding | -0.32 | 0.14 | -0.62 | -0.02 | 340 |
| **GS** | Life satisfaction | 1 | sex | male | -0.33 | 0.09 | -0.51 | -0.16 | 1146 |
| **GS** | Life satisfaction | 1 | sex | female | -0.32 | 0.07 | -0.44 | -0.19 | 1836 |
| **GS** | Life satisfaction | 2 | sex | male | -0.27 | 0.10 | -0.46 | -0.09 | 822 |
| **GS** | Life satisfaction | 2 | sex | female | -0.25 | 0.07 | -0.38 | -0.12 | 1362 |
| **GS** | Life satisfaction | 3 | sex | male | -0.29 | 0.11 | -0.49 | -0.08 | 768 |
| **GS** | Life satisfaction | 3 | sex | female | -0.23 | 0.08 | -0.38 | -0.08 | 1256 |
| **GS** | Life satisfaction | 1 | Shielding | not shielding | -0.35 | 0.05 | -0.46 | -0.25 | 2874 |
| **GS** | Life satisfaction | 1 | Shielding | shielding | -0.39 | 0.26 | -0.93 | 0.15 | 108 |
| **GS** | Life satisfaction | 2 | Shielding | not shielding | -0.27 | 0.06 | -0.38 | -0.17 | 2103 |
| **GS** | Life satisfaction | 2 | Shielding | shielding | -0.41 | 0.28 | -0.98 | 0.15 | 81 |
| **GS** | Life satisfaction | 3 | Shielding | not shielding | -0.27 | 0.06 | -0.39 | -0.14 | 1951 |
| **GS** | Life satisfaction | 3 | Shielding | shielding | -1.06 | 0.33 | -1.67 | -0.45 | 73 |
| **GS** | Life satisfaction | 1 | T1 loneliness | not lonely | -0.24 | 0.05 | -0.35 | -0.14 | 2846 |
| **GS** | Life satisfaction | 1 | T1 loneliness | lonely | 0.12 | 0.17 | -0.22 | 0.46 | 122 |
| **GS** | Life satisfaction | 2 | T1 loneliness | not lonely | -0.20 | 0.06 | -0.30 | -0.09 | 2088 |
| **GS** | Life satisfaction | 2 | T1 loneliness | lonely | 0.08 | 0.27 | -0.43 | 0.58 | 84 |
| **GS** | Life satisfaction | 3 | T1 loneliness | not lonely | -0.24 | 0.07 | -0.37 | -0.12 | 1931 |
| **GS** | Life satisfaction | 3 | T1 loneliness | lonely | 0.27 | 0.28 | -0.26 | 0.81 | 82 |
| **GS** | Psych distress | 0 | Prior MH | not shielding | 0.02 | 0.02 | -0.02 | 0.06 | 2458 |
| **GS** | Psych distress | 0 | Prior MH | shielding | 0.20 | 0.08 | 0.05 | 0.34 | 526 |
| **GS** | Psych distress | 1 | Prior MH | not shielding | 0.13 | 0.06 | 0.03 | 0.23 | 2390 |
| **GS** | Psych distress | 1 | Prior MH | shielding | 0.20 | 0.18 | -0.12 | 0.52 | 492 |
| **GS** | Psych distress | 2 | Prior MH | not shielding | 0.22 | 0.06 | 0.11 | 0.34 | 1747 |
| **GS** | Psych distress | 2 | Prior MH | shielding | 0.15 | 0.20 | -0.22 | 0.52 | 359 |
| **GS** | Psych distress | 3 | Prior MH | not shielding | 0.16 | 0.07 | 0.03 | 0.30 | 1612 |
| **GS** | Psych distress | 3 | Prior MH | shielding | 0.12 | 0.21 | -0.26 | 0.50 | 320 |
| **GS** | Psych distress | 0 | sex | male | 0.10 | 0.07 | -0.01 | 0.22 | 1148 |
| **GS** | Psych distress | 0 | sex | female | 0.12 | 0.05 | 0.04 | 0.21 | 1836 |
| **GS** | Psych distress | 1 | sex | male | 0.24 | 0.12 | 0.06 | 0.42 | 1110 |

| | | | | | | | | | |
|---|---|---|---|---|---|---|---|---|---|
| **GS** | Psych distress | 1 | sex | female | 0.09 | 0.07 | -0.04 | 0.22 | 1772 |
| **GS** | Psych distress | 2 | sex | male | 0.33 | 0.13 | 0.12 | 0.54 | 805 |
| **GS** | Psych distress | 2 | sex | female | 0.18 | 0.08 | 0.03 | 0.33 | 1301 |
| **GS** | Psych distress | 3 | sex | male | 0.39 | 0.14 | 0.16 | 0.61 | 742 |
| **GS** | Psych distress | 3 | sex | female | 0.08 | 0.09 | -0.09 | 0.25 | 1190 |
| **GS** | Psych distress | 0 | Shielding | not shielding | 0.12 | 0.04 | 0.05 | 0.19 | 2876 |
| **GS** | Psych distress | 0 | Shielding | shielding | 0.16 | 0.30 | -0.26 | 0.58 | 108 |
| **GS** | Psych distress | 1 | Shielding | not shielding | 0.16 | 0.06 | 0.05 | 0.27 | 2783 |
| **GS** | Psych distress | 1 | Shielding | shielding | 0.36 | 0.37 | -0.32 | 1.04 | 99 |
| **GS** | Psych distress | 2 | Shielding | not shielding | 0.22 | 0.07 | 0.10 | 0.34 | 2026 |
| **GS** | Psych distress | 2 | Shielding | shielding | 0.90 | 0.42 | 0.24 | 1.56 | 80 |
| **GS** | Psych distress | 3 | Shielding | not shielding | 0.19 | 0.07 | 0.06 | 0.33 | 1864 |
| **GS** | Psych distress | 3 | Shielding | shielding | 0.84 | 0.58 | -0.01 | 1.70 | 68 |
| **GS** | Psych distress | 0 | T1 loneliness | not lonely | 0.05 | 0.04 | -0.02 | 0.12 | 2846 |
| **GS** | Psych distress | 0 | T1 loneliness | lonely | 0.12 | 0.19 | -0.27 | 0.50 | 122 |
| **GS** | Psych distress | 1 | T1 loneliness | not lonely | 0.00 | 0.05 | -0.11 | 0.10 | 2758 |
| **GS** | Psych distress | 1 | T1 loneliness | lonely | -0.27 | 0.27 | -0.83 | 0.30 | 113 |
| **GS** | Psych distress | 2 | T1 loneliness | not lonely | 0.11 | 0.06 | -0.01 | 0.23 | 2018 |
| **GS** | Psych distress | 2 | T1 loneliness | lonely | -0.52 | 0.34 | -1.22 | 0.19 | 78 |
| **GS** | Psych distress | 3 | T1 loneliness | not lonely | 0.09 | 0.07 | -0.04 | 0.23 | 1846 |
| **GS** | Psych distress | 3 | T1 loneliness | lonely | -0.29 | 0.32 | -0.95 | 0.36 | 77 |
| **BCS** | Psych distress | T0 | Loneliness at T0 | not lonely | -0.02 | 0.07 | -0.16 | 0.12 | 2266 |
| **BCS** | Psych distress | T0 | Loneliness at T0 | lonely | -0.19 | 0.28 | -0.75 | 0.36 | 139 |
| **BCS** | Psych distress | T1 | Loneliness at T1 | not lonely | -0.11 | 0.06 | -0.23 | 0.02 | 2456 |
| **BCS** | Psych distress | T1 | Loneliness at T1 | lonely | -0.45 | 0.19 | -0.83 | -0.07 | 152 |
| **BCS** | Psych distress | T2 | Loneliness at T1 | not lonely | 0.00 | 0.07 | -0.13 | 0.13 | 2459 |
| **BCS** | Psych distress | T2 | Loneliness at T1 | lonely | -0.47 | 0.22 | -0.91 | -0.03 | 152 |
| **BCS** | Psych distress | T3 | Loneliness at T1 | not lonely | 0.06 | 0.06 | -0.06 | 0.19 | 2221 |
| **BCS** | Psych distress | T3 | Loneliness at T1 | lonely | -0.24 | 0.21 | -0.66 | 0.18 | 135 |
| **BCS** | Psych distress | T0 | Sex | male | -0.12 | 0.09 | -0.30 | 0.05 | 961 |
| **BCS** | Psych distress | T0 | Sex | female | 0.14 | 0.11 | -0.08 | 0.37 | 1500 |
| **BCS** | Psych distress | T1 | Sex | male | -0.07 | 0.10 | -0.26 | 0.12 | 1054 |
| **BCS** | Psych distress | T1 | Sex | female | -0.02 | 0.10 | -0.22 | 0.17 | 1562 |

| | | | | | | | | | |
|---|---|---|---|---|---|---|---|---|---|
| **BCS** | Psych distress | T2 | Sex | male | 0.03 | 0.09 | -0.15 | 0.21 | 1073 |
| **BCS** | Psych distress | T2 | Sex | female | 0.05 | 0.09 | -0.13 | 0.23 | 1600 |
| **BCS** | Psych distress | T3 | Sex | male | 0.10 | 0.09 | -0.08 | 0.27 | 938 |
| **BCS** | Psych distress | T3 | Sex | female | 0.15 | 0.09 | -0.02 | 0.32 | 1468 |
| **BCS** | Psych distress | T0 | Shielding status at T1 | not shielding | 0.01 | 0.07 | -0.14 | 0.15 | 2333 |
| **BCS** | Psych distress | T0 | Shielding status at T1 | shielding | -0.12 | 0.35 | -0.82 | 0.59 | 122 |
| **BCS** | Psych distress | T1 | Shielding status at T1 | not shielding | -0.02 | 0.07 | -0.15 | 0.11 | 2483 |
| **BCS** | Psych distress | T1 | Shielding status at T1 | shielding | -0.31 | 0.33 | -0.96 | 0.34 | 126 |
| **BCS** | Psych distress | T2 | Shielding status at T1 | not shielding | 0.02 | 0.07 | -0.11 | 0.15 | 2537 |
| **BCS** | Psych distress | T2 | Shielding status at T1 | shielding | 0.39 | 0.30 | -0.20 | 0.97 | 129 |
| **BCS** | Psych distress | T3 | Shielding status at T1 | not shielding | 0.11 | 0.06 | -0.01 | 0.24 | 2280 |
| **BCS** | Psych distress | T3 | Shielding status at T1 | shielding | 0.32 | 0.31 | -0.29 | 0.93 | 121 |
| **BCS** | Psych distress | T0 | Prior mental ill-health | no prior MH | -0.02 | 0.04 | -0.10 | 0.07 | 2095 |
| **BCS** | Psych distress | T0 | Prior mental ill-health | prior MH | -0.03 | 0.12 | -0.26 | 0.20 | 366 |
| **BCS** | Psych distress | T1 | Prior mental ill-health | no prior MH | -0.04 | 0.06 | -0.16 | 0.07 | 2216 |
| **BCS** | Psych distress | T1 | Prior mental ill-health | prior MH | -0.09 | 0.18 | -0.45 | 0.28 | 400 |
| **BCS** | Psych distress | T2 | Prior mental ill-health | no prior MH | -0.07 | 0.06 | -0.18 | 0.04 | 2266 |
| **BCS** | Psych distress | T2 | Prior mental ill-health | prior MH | 0.29 | 0.16 | -0.02 | 0.60 | 407 |
| **BCS** | Psych distress | T3 | Prior mental ill-health | no prior MH | 0.04 | 0.05 | -0.07 | 0.15 | 2049 |
| **BCS** | Psych distress | T3 | Prior mental ill-health | prior MH | 0.27 | 0.16 | -0.05 | 0.60 | 357 |
| **BCS** | Life satisfaction | T0 | Loneliness at T0 | not lonely | -0.32 | 0.07 | -0.45 | -0.19 | 2264 |
| **BCS** | Life satisfaction | T0 | Loneliness at T0 | lonely | -0.68 | 0.23 | -1.15 | -0.22 | 137 |
| **BCS** | Life satisfaction | T1 | Loneliness at T1 | not lonely | -0.02 | 0.07 | -0.16 | 0.12 | 2474 |
| **BCS** | Life satisfaction | T1 | Loneliness at T1 | lonely | -0.01 | 0.21 | -0.43 | 0.40 | 154 |
| **BCS** | Life satisfaction | T2 | Loneliness at T1 | not lonely | -0.21 | 0.06 | -0.33 | -0.10 | 2463 |
| **BCS** | Life satisfaction | T2 | Loneliness at T1 | lonely | -0.23 | 0.20 | -0.63 | 0.16 | 153 |
| **BCS** | Life satisfaction | T3 | Loneliness at T1 | not lonely | -0.10 | 0.07 | -0.23 | 0.03 | 2226 |
| **BCS** | Life satisfaction | T3 | Loneliness at T1 | lonely | 0.07 | 0.21 | -0.34 | 0.48 | 136 |
| **BCS** | Life satisfaction | T0 | Sex | male | -0.48 | 0.10 | -0.67 | -0.29 | 959 |
| **BCS** | Life satisfaction | T0 | Sex | female | -0.40 | 0.10 | -0.60 | -0.21 | 1498 |
| **BCS** | Life satisfaction | T1 | Sex | male | -0.13 | 0.13 | -0.38 | 0.12 | 1059 |
| **BCS** | Life satisfaction | T1 | Sex | female | -0.18 | 0.09 | -0.35 | -0.01 | 1579 |
| **BCS** | Life satisfaction | T2 | Sex | male | -0.43 | 0.09 | -0.61 | -0.26 | 1077 |
| **BCS** | Life satisfaction | T2 | Sex | female | -0.25 | 0.08 | -0.40 | -0.09 | 1601 |

| | | | | | | | | | |
|---|---|---|---|---|---|---|---|---|---|
| **BCS** | Life satisfaction | T3 | Sex | male | -0.26 | 0.10 | -0.46 | -0.06 | 940 |
| **BCS** | Life satisfaction | T3 | Sex | female | -0.14 | 0.08 | -0.30 | 0.03 | 1472 |
| **BCS** | Life satisfaction | T0 | Shielding status at T1 | not shielding | -0.45 | 0.07 | -0.59 | -0.32 | 2329 |
| **BCS** | Life satisfaction | T0 | Shielding status at T1 | shielding | -0.06 | 0.33 | -0.71 | 0.58 | 122 |
| **BCS** | Life satisfaction | T1 | Shielding status at T1 | not shielding | -0.18 | 0.08 | -0.34 | -0.01 | 2504 |
| **BCS** | Life satisfaction | T1 | Shielding status at T1 | shielding | 0.20 | 0.41 | -0.60 | 1.01 | 127 |
| **BCS** | Life satisfaction | T2 | Shielding status at T1 | not shielding | -0.31 | 0.06 | -0.43 | -0.19 | 2542 |
| **BCS** | Life satisfaction | T2 | Shielding status at T1 | shielding | -0.41 | 0.30 | -1.00 | 0.19 | 129 |
| **BCS** | Life satisfaction | T3 | Shielding status at T1 | not shielding | -0.17 | 0.07 | -0.30 | -0.04 | 2286 |
| **BCS** | Life satisfaction | T3 | Shielding status at T1 | shielding | -0.25 | 0.30 | -0.83 | 0.34 | 121 |
| **BCS** | Life satisfaction | T0 | Prior mental ill-health | no prior MH | -0.38 | 0.07 | -0.51 | -0.25 | 2091 |
| **BCS** | Life satisfaction | T0 | Prior mental ill-health | prior MH | -0.63 | 0.19 | -1.00 | -0.25 | 366 |
| **BCS** | Life satisfaction | T1 | Prior mental ill-health | no prior MH | -0.15 | 0.08 | -0.31 | 0.01 | 2234 |
| **BCS** | Life satisfaction | T1 | Prior mental ill-health | prior MH | -0.21 | 0.20 | -0.61 | 0.19 | 404 |
| **BCS** | Life satisfaction | T2 | Prior mental ill-health | no prior MH | -0.28 | 0.06 | -0.41 | -0.16 | 2270 |
| **BCS** | Life satisfaction | T2 | Prior mental ill-health | prior MH | -0.37 | 0.16 | -0.68 | -0.06 | 408 |
| **BCS** | Life satisfaction | T3 | Prior mental ill-health | no prior MH | -0.19 | 0.07 | -0.32 | -0.05 | 2054 |
| **BCS** | Life satisfaction | T3 | Prior mental ill-health | prior MH | -0.09 | 0.17 | -0.42 | 0.24 | 358 |
| **NS** | Psych distress | T0 | Loneliness at T0 | not lonely | 0.04 | 0.08 | -0.11 | 0.19 | 968 |
| **NS** | Psych distress | T0 | Loneliness at T0 | lonely | -0.05 | 0.31 | -0.67 | 0.58 | 112 |
| **NS** | Psych distress | T1 | Loneliness at T1 | not lonely | 0.27 | 0.12 | 0.04 | 0.49 | 1017 |
| **NS** | Psych distress | T1 | Loneliness at T1 | lonely | 0.17 | 0.31 | -0.45 | 0.79 | 118 |
| **NS** | Psych distress | T2 | Loneliness at T1 | not lonely | 0.23 | 0.10 | 0.03 | 0.43 | 1023 |
| **NS** | Psych distress | T2 | Loneliness at T1 | lonely | -0.37 | 0.24 | -0.84 | 0.10 | 118 |
| **NS** | Psych distress | T3 | Loneliness at T1 | not lonely | 0.21 | 0.10 | 0.03 | 0.40 | 961 |
| **NS** | Psych distress | T3 | Loneliness at T1 | lonely | -0.49 | 0.25 | -0.98 | 0.00 | 110 |
| **NS** | Psych distress | T0 | Sex | male | 0.27 | 0.11 | 0.05 | 0.50 | 380 |
| **NS** | Psych distress | T0 | Sex | female | -0.11 | 0.10 | -0.31 | 0.09 | 734 |
| **NS** | Psych distress | T1 | Sex | male | 0.31 | 0.17 | -0.02 | 0.64 | 389 |
| **NS** | Psych distress | T1 | Sex | female | 0.43 | 0.17 | 0.10 | 0.76 | 746 |
| **NS** | Psych distress | T2 | Sex | male | 0.23 | 0.14 | -0.03 | 0.50 | 401 |
| **NS** | Psych distress | T2 | Sex | female | 0.12 | 0.12 | -0.12 | 0.37 | 781 |
| **NS** | Psych distress | T3 | Sex | male | 0.30 | 0.14 | 0.03 | 0.58 | 379 |

| | | | | | | | | | |
|---|---|---|---|---|---|---|---|---|---|
| **NS** | Psych distress | T3 | Sex | female | 0.04 | 0.11 | -0.18 | 0.27 | 724 |
| **NS** | Psych distress | T0 | Shielding status at T1 | not shielding | 0.07 | 0.08 | -0.08 | 0.23 | 1078 |
| **NS** | Psych distress | T0 | Shielding status at T1 | shielding | -0.66 | 0.40 | -1.51 | 0.19 | 32 |
| **NS** | Psych distress | T1 | Shielding status at T1 | not shielding | 0.36 | 0.14 | 0.09 | 0.62 | 1099 |
| **NS** | Psych distress | T1 | Shielding status at T1 | shielding | 0.43 | 0.34 | -0.28 | 1.15 | 32 |
| **NS** | Psych distress | T2 | Shielding status at T1 | not shielding | 0.17 | 0.10 | -0.02 | 0.36 | 1144 |
| **NS** | Psych distress | T2 | Shielding status at T1 | shielding | -0.04 | 0.36 | -0.79 | 0.72 | 34 |
| **NS** | Psych distress | T3 | Shielding status at T1 | not shielding | 0.15 | 0.09 | -0.03 | 0.33 | |
| **NS** | Psych distress | T3 | Shielding status at T1 | shielding | -0.17 | 0.62 | -1.49 | 1.15 | 31 |
| **NS** | Psych distress | T0 | Prior mental ill-health | no prior MH | 0.00 | 0.05 | -0.10 | 0.11 | 849 |
| **NS** | Psych distress | T0 | Prior mental ill-health | prior MH | 0.12 | 0.13 | -0.14 | 0.37 | 265 |
| **NS** | Psych distress | T1 | Prior mental ill-health | no prior MH | 0.29 | 0.17 | -0.04 | 0.62 | 864 |
| **NS** | Psych distress | T1 | Prior mental ill-health | prior MH | 0.23 | 0.22 | -0.22 | 0.67 | 271 |
| **NS** | Psych distress | T2 | Prior mental ill-health | no prior MH | 0.12 | 0.11 | -0.10 | 0.34 | 902 |
| **NS** | Psych distress | T2 | Prior mental ill-health | prior MH | 0.17 | 0.20 | -0.22 | 0.55 | 280 |
| **NS** | Psych distress | T3 | Prior mental ill-health | no prior MH | 0.15 | 0.09 | -0.03 | 0.33 | 841 |
| **NS** | Psych distress | T3 | Prior mental ill-health | prior MH | 0.10 | 0.20 | -0.30 | 0.50 | 262 |
| **NS** | Life satisfaction | T0 | Loneliness at T0 | not lonely | 0.04 | 0.02 | 0.00 | 0.09 | 968 |
| **NS** | Life satisfaction | T0 | Loneliness at T0 | lonely | 0.01 | 0.08 | -0.14 | 0.16 | 112 |
| **NS** | Life satisfaction | T1 | Loneliness at T1 | not lonely | -0.16 | 0.08 | -0.32 | 0.00 | 1037 |
| **NS** | Life satisfaction | T1 | Loneliness at T1 | lonely | -0.34 | 0.20 | -0.73 | 0.06 | 120 |
| **NS** | Life satisfaction | T2 | Loneliness at T1 | not lonely | -0.10 | 0.06 | -0.23 | 0.02 | 1027 |
| **NS** | Life satisfaction | T2 | Loneliness at T1 | lonely | 0.03 | 0.15 | -0.28 | 0.34 | 118 |
| **NS** | Life satisfaction | T3 | Loneliness at T1 | not lonely | -0.06 | 0.06 | -0.19 | 0.06 | 964 |
| **NS** | Life satisfaction | T3 | Loneliness at T1 | lonely | 0.28 | 0.16 | -0.04 | 0.60 | 110 |
| **NS** | Life satisfaction | T0 | Sex | male | 0.09 | 0.03 | 0.02 | 0.15 | 380 |
| **NS** | Life satisfaction | T0 | Sex | female | 0.02 | 0.03 | -0.04 | 0.07 | 734 |
| **NS** | Life satisfaction | T1 | Sex | male | -0.28 | 0.11 | -0.49 | -0.08 | 396 |
| **NS** | Life satisfaction | T1 | Sex | female | -0.25 | 0.17 | -0.58 | 0.08 | 762 |
| **NS** | Life satisfaction | T2 | Sex | male | -0.09 | 0.08 | -0.25 | 0.08 | 404 |
| **NS** | Life satisfaction | T2 | Sex | female | -0.13 | 0.08 | -0.29 | 0.03 | 783 |
| **NS** | Life satisfaction | T3 | Sex | male | -0.18 | 0.09 | -0.37 | 0.00 | 379 |
| **NS** | Life satisfaction | T3 | Sex | female | 0.05 | 0.08 | -0.10 | 0.20 | 727 |
| **NS** | Life satisfaction | T0 | Shielding status at T1 | not shielding | 0.05 | 0.02 | 0.01 | 0.10 | 1078 |

| | | | | | | | | | |
|---|---|---|---|---|---|---|---|---|---|
| **NS** | Life satisfaction | T0 | Shielding status at T1 | shielding | -0.13 | 0.09 | -0.32 | 0.05 | 32 |
| **NS** | Life satisfaction | T1 | Shielding status at T1 | not shielding | -0.26 | 0.11 | -0.48 | -0.03 | 1122 |
| **NS** | Life satisfaction | T1 | Shielding status at T1 | shielding | -0.31 | 0.28 | -0.89 | 0.28 | 32 |
| **NS** | Life satisfaction | T2 | Shielding status at T1 | not shielding | -0.12 | 0.06 | -0.25 | 0.00 | 1149 |
| **NS** | Life satisfaction | T2 | Shielding status at T1 | shielding | 0.23 | 0.25 | -0.30 | 0.76 | 34 |
| **NS** | Life satisfaction | T3 | Shielding status at T1 | not shielding | -0.06 | 0.06 | -0.18 | 0.07 | 1071 |
| **NS** | Life satisfaction | T3 | Shielding status at T1 | shielding | -0.17 | 0.29 | -0.80 | 0.46 | 31 |
| **NS** | Life satisfaction | T0 | Prior mental ill-health | no prior MH | 0.03 | 0.02 | -0.01 | 0.08 | 849 |
| **NS** | Life satisfaction | T0 | Prior mental ill-health | prior MH | 0.08 | 0.05 | -0.01 | 0.17 | 265 |
| **NS** | Life satisfaction | T1 | Prior mental ill-health | no prior MH | -0.33 | 0.13 | -0.58 | -0.08 | 879 |
| **NS** | Life satisfaction | T1 | Prior mental ill-health | prior MH | -0.03 | 0.18 | -0.39 | 0.34 | 279 |
| **NS** | Life satisfaction | T2 | Prior mental ill-health | no prior MH | -0.10 | 0.07 | -0.25 | 0.04 | 904 |
| **NS** | Life satisfaction | T2 | Prior mental ill-health | prior MH | -0.10 | 0.13 | -0.35 | 0.15 | 283 |
| **NS** | Life satisfaction | T3 | Prior mental ill-health | no prior MH | -0.02 | 0.07 | -0.16 | 0.11 | 844 |
| **NS** | Life satisfaction | T3 | Prior mental ill-health | prior MH | -0.08 | 0.13 | -0.34 | 0.18 | 262 |
| **ELSA** | Psych distress | T0 | Loneliness at T0 | not lonely | 0.07 | 0.04 | 0.00 | 0.15 | 4837 |
| **ELSA** | Psych distress | T0 | Loneliness at T0 | lonely | -0.19 | 0.17 | -0.52 | 0.14 | 280 |
| **ELSA** | Psych distress | T1 | Loneliness at T1 | not lonely | 0.13 | 0.04 | 0.04 | 0.22 | 5161 |
| **ELSA** | Psych distress | T1 | Loneliness at T1 | lonely | 0.09 | 0.12 | -0.14 | 0.31 | 304 |
| **ELSA** | Psych distress | T3 | Loneliness at T1 | not lonely | 0.10 | 0.04 | 0.01 | 0.18 | 4771 |
| **ELSA** | Psych distress | T3 | Loneliness at T1 | lonely | 0.30 | 0.13 | 0.05 | 0.56 | 271 |
| **ELSA** | Psych distress | T0 | Sex | male | 0.16 | 0.07 | 0.03 | 0.29 | 2336 |
| **ELSA** | Psych distress | T0 | Sex | female | 0.23 | 0.05 | 0.12 | 0.34 | 3135 |
| **ELSA** | Psych distress | T1 | Sex | male | 0.25 | 0.08 | 0.10 | 0.40 | 2335 |
| **ELSA** | Psych distress | T1 | Sex | female | 0.29 | 0.05 | 0.19 | 0.40 | 3135 |
| **ELSA** | Psych distress | T3 | Sex | male | 0.28 | 0.07 | 0.14 | 0.42 | 2164 |
| **ELSA** | Psych distress | T3 | Sex | female | 0.20 | 0.05 | 0.09 | 0.30 | 2882 |
| **ELSA** | Psych distress | T0 | Shielding status at T1 | not shielding | 0.22 | 0.05 | 0.13 | 0.31 | 4543 |
| **ELSA** | Psych distress | T0 | Shielding status at T1 | shielding | 0.09 | 0.10 | -0.11 | 0.29 | 921 |
| **ELSA** | Psych distress | T1 | Shielding status at T1 | not shielding | 0.28 | 0.05 | 0.18 | 0.37 | 4542 |
| **ELSA** | Psych distress | T1 | Shielding status at T1 | shielding | 0.18 | 0.10 | -0.01 | 0.38 | 921 |
| **ELSA** | Psych distress | T3 | Shielding status at T1 | not shielding | 0.22 | 0.05 | 0.12 | 0.31 | 4240 |
| **ELSA** | Psych distress | T3 | Shielding status at T1 | shielding | 0.26 | 0.10 | 0.07 | 0.45 | 800 |

| | | | | | | | | | |
|---|---|---|---|---|---|---|---|---|---|
| **ELSA** | Psych distress | T1 | Prior mental ill-health | good MH | 0.20 | 0.05 | 0.11 | 0.29 | 4880 |
| **ELSA** | Psych distress | T1 | Prior mental ill-health | ill health | 0.22 | 0.11 | 0.01 | 0.44 | 590 |
| **ELSA** | Psych distress | T3 | Prior mental ill-health | good MH | 0.15 | 0.04 | 0.07 | 0.24 | 4528 |
| **ELSA** | Psych distress | T3 | Prior mental ill-health | ill health | 0.18 | 0.11 | -0.03 | 0.40 | 518 |
| **ELSA** | Life satisfaction | T0 | Loneliness at T0 | not lonely | -0.23 | 0.04 | -0.31 | -0.14 | 4790 |
| **ELSA** | Life satisfaction | T0 | Loneliness at T0 | lonely | -0.08 | 0.16 | -0.39 | 0.23 | 268 |
| **ELSA** | Life satisfaction | T1 | Loneliness at T1 | not lonely | -0.04 | 0.04 | -0.12 | 0.04 | 5145 |
| **ELSA** | Life satisfaction | T1 | Loneliness at T1 | lonely | 0.16 | 0.17 | -0.16 | 0.49 | 303 |
| **ELSA** | Life satisfaction | T3 | Loneliness at T1 | not lonely | -0.12 | 0.05 | -0.20 | -0.03 | 4764 |
| **ELSA** | Life satisfaction | T3 | Loneliness at T1 | lonely | -0.18 | 0.18 | -0.54 | 0.17 | 270 |
| **ELSA** | Life satisfaction | T0 | Sex | male | -0.30 | 0.07 | -0.44 | -0.16 | 2193 |
| **ELSA** | Life satisfaction | T0 | Sex | female | -0.33 | 0.05 | -0.43 | -0.23 | 2897 |
| **ELSA** | Life satisfaction | T1 | Sex | male | -0.11 | 0.07 | -0.25 | 0.04 | 2333 |
| **ELSA** | Life satisfaction | T1 | Sex | female | -0.14 | 0.05 | -0.25 | -0.04 | 3120 |
| **ELSA** | Life satisfaction | T3 | Sex | male | -0.20 | 0.08 | -0.36 | -0.05 | 2161 |
| **ELSA** | Life satisfaction | T3 | Sex | female | -0.21 | 0.05 | -0.31 | -0.10 | 2877 |
| **ELSA** | Life satisfaction | T0 | Shielding status at T1 | not shielding | -0.33 | 0.05 | -0.42 | -0.24 | 4261 |
| **ELSA** | Life satisfaction | T0 | Shielding status at T1 | shielding | -0.31 | 0.11 | -0.52 | -0.09 | 824 |
| **ELSA** | Life satisfaction | T1 | Shielding status at T1 | not shielding | -0.13 | 0.05 | -0.22 | -0.04 | 4528 |
| **ELSA** | Life satisfaction | T1 | Shielding status at T1 | shielding | -0.08 | 0.11 | -0.29 | 0.13 | 918 |
| **ELSA** | Life satisfaction | T3 | Shielding status at T1 | not shielding | -0.21 | 0.05 | -0.31 | -0.11 | 4233 |
| **ELSA** | Life satisfaction | T3 | Shielding status at T1 | shielding | -0.21 | 0.10 | -0.40 | -0.01 | 798 |
| **ELSA** | Life satisfaction | T0 | Prior mental ill-health | good MH | -0.27 | 0.04 | -0.35 | -0.19 | 4589 |
| **ELSA** | Life satisfaction | T0 | Prior mental ill-health | ill health | -0.13 | 0.13 | -0.38 | 0.13 | 501 |
| **ELSA** | Life satisfaction | T1 | Prior mental ill-health | good MH | -0.10 | 0.05 | -0.19 | -0.01 | 4866 |
| **ELSA** | Life satisfaction | T1 | Prior mental ill-health | ill health | 0.04 | 0.11 | -0.18 | 0.27 | 587 |
| **ELSA** | Life satisfaction | T3 | Prior mental ill-health | good MH | -0.18 | 0.05 | -0.27 | -0.08 | 4523 |
| **ELSA** | Life satisfaction | T3 | Prior mental ill-health | ill health | -0.09 | 0.13 | -0.33 | 0.16 | 515 |
| **USOC** | Psych distress | T0 | Shielding status at T1 | not shielding | 0.06 | 0.03 | -0.01 | 0.13 | 12200 |
| **USOC** | Psych distress | T0 | Shielding status at T1 | shielding | 0.19 | 0.12 | -0.04 | 0.42 | 784 |
| **USOC** | Life satisfaction | T0 | Shielding status at T1 | not shielding | -0.10 | 0.03 | -0.16 | -0.03 | 12200 |
| **USOC** | Life satisfaction | T0 | Shielding status at T1 | shielding | -0.11 | 0.11 | -0.33 | 0.12 | 784 |
| **USOC** | Life satisfaction | T1 | Shielding status at T1 | not shielding | -0.25 | 0.04 | -0.33 | -0.17 | 9530 |

| | | | | | | | | | |
|---|---|---|---|---|---|---|---|---|---|
| **USOC** | Life satisfaction | T1 | Shielding status at T1 | shielding | -0.32 | 0.14 | -0.59 | -0.04 | 602 |
| **USOC** | Psych distress | T0 | Loneliness at T0 | not lonely | -0.03 | 0.02 | -0.07 | 0.01 | 20541 |
| **USOC** | Psych distress | T0 | Loneliness at T0 | lonely | -0.13 | 0.10 | -0.33 | 0.06 | 1646 |
| **USOC** | Life satisfaction | T0 | Loneliness at T0 | not lonely | -0.01 | 0.02 | -0.05 | 0.04 | 20541 |
| **USOC** | Life satisfaction | T0 | Loneliness at T0 | lonely | -0.03 | 0.08 | -0.20 | 0.13 | 1646 |
| **USOC** | Life satisfaction | T1 | Loneliness at T1 | not lonely | -0.18 | 0.04 | -0.27 | -0.10 | 9470 |
| **USOC** | Life satisfaction | T1 | Loneliness at T1 | lonely | -0.05 | 0.11 | -0.28 | 0.17 | 662 |
| **USOC** | Psych distress | T0 | Sex | male | 0.03 | 0.03 | -0.03 | 0.10 | 9917 |
| **USOC** | Psych distress | T0 | Sex | female | 0.02 | 0.03 | -0.04 | 0.08 | 12270 |
| **USOC** | Life satisfaction | T0 | Sex | male | -0.09 | 0.03 | -0.16 | -0.02 | 9917 |
| **USOC** | Life satisfaction | T0 | Sex | female | -0.04 | 0.03 | -0.10 | 0.02 | 12270 |
| **USOC** | Life satisfaction | T1 | Sex | male | -0.32 | 0.07 | -0.45 | -0.18 | 4210 |
| **USOC** | Life satisfaction | T1 | Sex | female | -0.22 | 0.05 | -0.31 | -0.12 | 5922 |
| **USOC** | Life satisfaction | T1 | Prior mental ill-health | good MH | -0.22 | 0.05 | -0.31 | -0.13 | 8341 |
| **USOC** | Life satisfaction | T1 | Prior mental ill-health | ill health | -0.33 | 0.08 | -0.49 | -0.17 | 1791 |
| **USOC** | Psych distress | T0 | Age | 16-24 | 0.28 | 0.14 | 0.00 | 0.55 | 2615 |
| **USOC** | Psych distress | T0 | Age | 25-34 | 0.01 | 0.08 | -0.16 | 0.17 | 2522 |
| **USOC** | Psych distress | T0 | Age | 35-44 | 0.00 | 0.09 | -0.17 | 0.18 | 3227 |
| **USOC** | Psych distress | T0 | Age | 45-54 | 0.02 | 0.06 | -0.10 | 0.14 | 4088 |
| **USOC** | Psych distress | T0 | Age | 55-64 | 0.13 | 0.05 | 0.02 | 0.23 | 3978 |
| **USOC** | Psych distress | T0 | Age | 65-74 | 0.03 | 0.04 | -0.04 | 0.10 | 3503 |
| **USOC** | Psych distress | T0 | Age | 75+ | -0.07 | 0.04 | -0.14 | 0.00 | 2254 |
| **USOC** | Life satisfaction | T0 | Age | 16-24 | -0.12 | 0.13 | -0.36 | 0.13 | 2615 |
| **USOC** | Life satisfaction | T0 | Age | 25-34 | -0.05 | 0.07 | -0.18 | 0.09 | 2522 |
| **USOC** | Life satisfaction | T0 | Age | 35-44 | -0.12 | 0.09 | -0.29 | 0.04 | 3227 |
| **USOC** | Life satisfaction | T0 | Age | 45-54 | -0.05 | 0.06 | -0.18 | 0.07 | 4088 |
| **USOC** | Life satisfaction | T0 | Age | 55-64 | -0.14 | 0.05 | -0.24 | -0.04 | 3978 |
| **USOC** | Life satisfaction | T0 | Age | 65-74 | -0.09 | 0.04 | -0.18 | -0.01 | 3503 |
| **USOC** | Life satisfaction | T0 | Age | 75+ | 0.04 | 0.05 | -0.06 | 0.13 | 2254 |
| **USOC** | Life satisfaction | T1 | Age | 16-24 | -0.05 | 0.37 | -0.77 | 0.67 | 651 |
| **USOC** | Life satisfaction | T1 | Age | 25-34 | -0.56 | 0.16 | -0.88 | -0.24 | 935 |
| **USOC** | Life satisfaction | T1 | Age | 35-44 | -0.50 | 0.14 | -0.78 | -0.22 | 1506 |
| **USOC** | Life satisfaction | T1 | Age | 45-54 | -0.11 | 0.11 | -0.32 | 0.10 | 2069 |
| **USOC** | Life satisfaction | T1 | Age | 55-64 | -0.27 | 0.07 | -0.42 | -0.13 | 2255 |

| | | | | | | | | | |
|---|---|---|---|---|---|---|---|---|---|
| **USOC** | Life satisfaction | T1 | Age | 65-74 | -0.28 | 0.07 | -0.42 | -0.14 | 1986 |
| **USOC** | Life satisfaction | T1 | Age | 75+ | -0.12 | 0.11 | -0.33 | 0.09 | 1524 |
| **USOC** | Psych distress | T1 | Shielding status at T1 | not shielding | 0.17 | 0.03 | 0.11 | 0.23 | 11364 |
| **USOC** | Psych distress | T1 | Shielding status at T1 | shielding | 0.23 | 0.11 | 0.01 | 0.44 | 724 |
| **USOC** | Psych distress | T2 | Shielding status at T1 | not shielding | 0.14 | 0.03 | 0.08 | 0.20 | 9309 |
| **USOC** | Psych distress | T2 | Shielding status at T1 | shielding | 0.25 | 0.12 | 0.01 | 0.50 | 596 |
| **USOC** | Psych distress | T3 | Shielding status at T1 | not shielding | 0.12 | 0.03 | 0.06 | 0.18 | 9304 |
| **USOC** | Psych distress | T3 | Shielding status at T1 | shielding | 0.10 | 0.12 | -0.14 | 0.34 | 609 |
| **USOC** | Life satisfaction | T2 | Shielding status at T1 | not shielding | -0.16 | 0.03 | -0.22 | -0.10 | 9309 |
| **USOC** | Life satisfaction | T2 | Shielding status at T1 | shielding | -0.17 | 0.11 | -0.39 | 0.04 | 596 |
| **USOC** | Life satisfaction | T3 | Shielding status at T1 | not shielding | -0.19 | 0.03 | -0.25 | -0.13 | 9304 |
| **USOC** | Life satisfaction | T3 | Shielding status at T1 | shielding | -0.28 | 0.10 | -0.48 | -0.08 | 609 |
| **USOC** | Psych distress | T1 | Loneliness at T1 | not lonely | 0.04 | 0.03 | -0.01 | 0.09 | 11234 |
| **USOC** | Psych distress | T1 | Loneliness at T1 | lonely | -0.06 | 0.10 | -0.27 | 0.14 | 854 |
| **USOC** | Psych distress | T2 | Loneliness at T1 | not lonely | 0.05 | 0.03 | -0.01 | 0.11 | 9258 |
| **USOC** | Psych distress | T2 | Loneliness at T1 | lonely | -0.05 | 0.13 | -0.30 | 0.20 | 647 |
| **USOC** | Psych distress | T3 | Loneliness at T1 | not lonely | 0.03 | 0.03 | -0.03 | 0.09 | 9259 |
| **USOC** | Psych distress | T3 | Loneliness at T1 | lonely | -0.20 | 0.12 | -0.44 | 0.04 | 654 |
| **USOC** | Life satisfaction | T2 | Loneliness at T1 | not lonely | -0.09 | 0.03 | -0.15 | -0.03 | 9258 |
| **USOC** | Life satisfaction | T2 | Loneliness at T1 | lonely | -0.07 | 0.09 | -0.24 | 0.10 | 647 |
| **USOC** | Life satisfaction | T3 | Loneliness at T1 | not lonely | -0.14 | 0.03 | -0.19 | -0.08 | 9259 |
| **USOC** | Life satisfaction | T3 | Loneliness at T1 | lonely | -0.02 | 0.09 | -0.20 | 0.15 | 654 |
| **USOC** | Psych distress | T1 | Sex | male | 0.21 | 0.05 | 0.12 | 0.30 | 5080 |
| **USOC** | Psych distress | T1 | Sex | female | 0.14 | 0.04 | 0.07 | 0.22 | 7008 |
| **USOC** | Psych distress | T2 | Sex | male | 0.20 | 0.05 | 0.10 | 0.30 | 4137 |
| **USOC** | Psych distress | T2 | Sex | female | 0.10 | 0.04 | 0.02 | 0.18 | 5768 |
| **USOC** | Psych distress | T3 | Sex | male | 0.14 | 0.05 | 0.04 | 0.24 | 4123 |
| **USOC** | Psych distress | T3 | Sex | female | 0.11 | 0.04 | 0.03 | 0.19 | 5790 |
| **USOC** | Life satisfaction | T2 | Sex | male | -0.27 | 0.05 | -0.37 | -0.18 | 4137 |
| **USOC** | Life satisfaction | T2 | Sex | female | -0.09 | 0.04 | -0.16 | -0.02 | 5768 |
| **USOC** | Life satisfaction | T3 | Sex | male | -0.32 | 0.05 | -0.41 | -0.23 | 4123 |
| **USOC** | Life satisfaction | T3 | Sex | female | -0.11 | 0.03 | -0.18 | -0.05 | 5790 |
| **USOC** | Psych distress | T1 | Age | 16-24 | -0.23 | 0.36 | -0.93 | 0.48 | 923 |
| **USOC** | Psych distress | T1 | Age | 25-34 | 0.34 | 0.14 | 0.06 | 0.62 | 1223 |

| | | | | | | | | | |
|---|---|---|---|---|---|---|---|---|---|
| **USOC** | Psych distress | T1 | Age | 35-44 | 0.08 | 0.13 | -0.17 | 0.34 | 1831 |
| **USOC** | Psych distress | T1 | Age | 45-54 | 0.21 | 0.08 | 0.05 | 0.37 | 2496 |
| **USOC** | Psych distress | T1 | Age | 55-64 | 0.18 | 0.06 | 0.07 | 0.29 | 2590 |
| **USOC** | Psych distress | T1 | Age | 65-74 | 0.21 | 0.05 | 0.11 | 0.30 | 2183 |
| **USOC** | Psych distress | T1 | Age | 75+ | 0.10 | 0.07 | -0.03 | 0.24 | 842 |
| **USOC** | Psych distress | T2 | Age | 16-24 | -0.31 | 0.29 | -0.88 | 0.26 | 590 |
| **USOC** | Psych distress | T2 | Age | 25-34 | 0.48 | 0.17 | 0.15 | 0.80 | 897 |
| **USOC** | Psych distress | T2 | Age | 35-44 | 0.24 | 0.14 | -0.05 | 0.52 | 1421 |
| **USOC** | Psych distress | T2 | Age | 45-54 | 0.17 | 0.09 | 0.00 | 0.34 | 1994 |
| **USOC** | Psych distress | T2 | Age | 55-64 | 0.15 | 0.06 | 0.04 | 0.27 | 2281 |
| **USOC** | Psych distress | T2 | Age | 65-74 | 0.13 | 0.05 | 0.03 | 0.24 | 1979 |
| **USOC** | Psych distress | T2 | Age | 75+ | -0.03 | 0.07 | -0.16 | 0.11 | 743 |
| **USOC** | Psych distress | T3 | Age | 16-24 | -0.07 | 0.38 | -0.83 | 0.68 | 599 |
| **USOC** | Psych distress | T3 | Age | 25-34 | 0.17 | 0.15 | -0.13 | 0.47 | 891 |
| **USOC** | Psych distress | T3 | Age | 35-44 | 0.15 | 0.13 | -0.10 | 0.40 | 1437 |
| **USOC** | Psych distress | T3 | Age | 45-54 | 0.15 | 0.08 | -0.01 | 0.31 | 2043 |
| **USOC** | Psych distress | T3 | Age | 55-64 | 0.11 | 0.06 | 0.00 | 0.23 | 2281 |
| **USOC** | Psych distress | T3 | Age | 65-74 | 0.14 | 0.05 | 0.04 | 0.25 | 1944 |
| **USOC** | Psych distress | T3 | Age | 75+ | 0.07 | 0.07 | -0.08 | 0.21 | 718 |
| **USOC** | Life satisfaction | T2 | Age | 16-24 | 0.07 | 0.19 | -0.31 | 0.44 | 590 |
| **USOC** | Life satisfaction | T2 | Age | 25-34 | -0.38 | 0.11 | -0.60 | -0.16 | 897 |
| **USOC** | Life satisfaction | T2 | Age | 35-44 | -0.44 | 0.11 | -0.66 | -0.23 | 1421 |
| **USOC** | Life satisfaction | T2 | Age | 45-54 | -0.19 | 0.08 | -0.35 | -0.04 | 1994 |
| **USOC** | Life satisfaction | T2 | Age | 55-64 | -0.25 | 0.06 | -0.37 | -0.14 | 2281 |
| **USOC** | Life satisfaction | T2 | Age | 65-74 | -0.10 | 0.06 | -0.21 | 0.02 | 1979 |
| **USOC** | Life satisfaction | T2 | Age | 75+ | 0.08 | 0.08 | -0.09 | 0.24 | 743 |
| **USOC** | Life satisfaction | T3 | Age | 16-24 | 0.22 | 0.21 | -0.20 | 0.63 | 599 |
| **USOC** | Life satisfaction | T3 | Age | 25-34 | -0.22 | 0.11 | -0.43 | -0.01 | 891 |
| **USOC** | Life satisfaction | T3 | Age | 35-44 | -0.26 | 0.10 | -0.45 | -0.06 | 1437 |
| **USOC** | Life satisfaction | T3 | Age | 45-54 | -0.19 | 0.07 | -0.32 | -0.05 | 2043 |
| **USOC** | Life satisfaction | T3 | Age | 55-64 | -0.26 | 0.06 | -0.37 | -0.16 | 2281 |
| **USOC** | Life satisfaction | T3 | Age | 65-74 | -0.16 | 0.05 | -0.27 | -0.06 | 1944 |
| **USOC** | Life satisfaction | T3 | Age | 75+ | -0.17 | 0.08 | -0.32 | -0.01 | 718 |
| **USOC** | Psych distress | T1 | Prior mental ill-health | good MH | 0.13 | 0.03 | 0.07 | 0.18 | 9887 |

| | | | | | | | | | |
|---|---|---|---|---|---|---|---|---|---|
| **USOC** | Psych distress | T1 | Prior mental ill-health | ill health | 0.21 | 0.08 | 0.05 | 0.37 | 2201 |
| **USOC** | Psych distress | T2 | Prior mental ill-health | good MH | 0.09 | 0.03 | 0.04 | 0.15 | 8164 |
| **USOC** | Psych distress | T2 | Prior mental ill-health | ill health | 0.22 | 0.09 | 0.04 | 0.39 | 1741 |
| **USOC** | Psych distress | T3 | Prior mental ill-health | good MH | 0.08 | 0.03 | 0.02 | 0.14 | 8156 |
| **USOC** | Psych distress | T3 | Prior mental ill-health | ill health | 0.10 | 0.09 | -0.07 | 0.28 | 1757 |
| **USOC** | Life satisfaction | T2 | Prior mental ill-health | good MH | -0.10 | 0.03 | -0.16 | -0.04 | 8164 |
| **USOC** | Life satisfaction | T2 | Prior mental ill-health | ill health | -0.32 | 0.07 | -0.45 | -0.19 | 1741 |
| **USOC** | Life satisfaction | T3 | Prior mental ill-health | good MH | -0.15 | 0.03 | -0.21 | -0.09 | 8156 |
| **USOC** | Life satisfaction | T3 | Prior mental ill-health | ill health | -0.27 | 0.06 | -0.39 | -0.14 | 1757 |
| **ALSPAC-G1** | Psych distress | T0 | Sex | male | 0.04 | 0.15 | -0.25 | 0.32 | 501 |
| **ALSPAC-G1** | Psych distress | T0 | Sex | female | 0.29 | 0.11 | 0.08 | 0.51 | 1308 |
| **ALSPAC-G1** | Psych distress | T1 | Sex | male | 0.21 | 0.13 | -0.04 | 0.46 | 586 |
| **ALSPAC-G1** | Psych distress | T1 | Sex | female | 0.30 | 0.11 | 0.09 | 0.51 | 1478 |
| **ALSPAC-G1** | Psych distress | T2 | Sex | male | 0.07 | 0.14 | -0.20 | 0.34 | 459 |
| **ALSPAC-G1** | Psych distress | T2 | Sex | female | 0.25 | 0.12 | 0.01 | 0.49 | 1151 |
| **ALSPAC-G1** | Psych distress | T3 | Sex | male | 0.18 | 0.14 | -0.10 | 0.47 | 539 |
| **ALSPAC-G1** | Psych distress | T3 | Sex | female | 0.31 | 0.11 | 0.09 | 0.52 | 1380 |
| **ALSPAC-G1** | Psych distress | T0 | Prior mental ill-health | no prior MH | 0.10 | 0.05 | -0.01 | 0.20 | 1404 |
| **ALSPAC-G1** | Psych distress | T0 | Prior mental ill-health | prior MH | -0.09 | 0.12 | -0.32 | 0.14 | 405 |
| **ALSPAC-G1** | Psych distress | T1 | Prior mental ill-health | no prior MH | 0.16 | 0.08 | 0.00 | 0.33 | 1376 |
| **ALSPAC-G1** | Psych distress | T1 | Prior mental ill-health | prior MH | 0.27 | 0.20 | -0.13 | 0.67 | 398 |
| **ALSPAC-G1** | Psych distress | T2 | Prior mental ill-health | no prior MH | 0.05 | 0.09 | -0.14 | 0.24 | 1098 |
| **ALSPAC-G1** | Psych distress | T2 | Prior mental ill-health | prior MH | 0.16 | 0.22 | -0.27 | 0.58 | 315 |
| **ALSPAC-G1** | Psych distress | T3 | Prior mental ill-health | no prior MH | 0.11 | 0.09 | -0.06 | 0.28 | 1300 |
| **ALSPAC-G1** | Psych distress | T3 | Prior mental ill-health | prior MH | 0.38 | 0.20 | -0.02 | 0.78 | 375 |
| **ALSPAC-G1** | Life satisfaction | T0 | Sex | male | -0.33 | 0.17 | -0.66 | 0.00 | 469 |
| **ALSPAC-G1** | Life satisfaction | T0 | Sex | female | -0.25 | 0.11 | -0.47 | -0.03 | 1251 |
| **ALSPAC-G1** | Life satisfaction | T1 | Sex | male | -0.38 | 0.16 | -0.69 | -0.08 | 587 |
| **ALSPAC-G1** | Life satisfaction | T1 | Sex | female | -0.33 | 0.10 | -0.53 | -0.13 | 1488 |
| **ALSPAC-G1** | Life satisfaction | T2 | Sex | male | -0.04 | 0.16 | -0.36 | 0.28 | 456 |
| **ALSPAC-G1** | Life satisfaction | T2 | Sex | female | -0.23 | 0.12 | -0.46 | 0.00 | 1160 |
| **ALSPAC-G1** | Life satisfaction | T3 | Sex | male | -0.23 | 0.16 | -0.55 | 0.09 | 533 |
| **ALSPAC-G1** | Life satisfaction | T3 | Sex | female | -0.33 | 0.11 | -0.53 | -0.12 | 1387 |

| | | | | | | | | | |
|---|---|---|---|---|---|---|---|---|---|
| **ALSPAC-G1** | Life satisfaction | T0 | Prior mental ill-health | no prior MH | -0.20 | 0.08 | -0.35 | -0.06 | 1439 |
| **ALSPAC-G1** | Life satisfaction | T0 | Prior mental ill-health | prior MH | 0.03 | 0.10 | -0.17 | 0.24 | 281 |
| **ALSPAC-G1** | Life satisfaction | T1 | Prior mental ill-health | no prior MH | -0.38 | 0.10 | -0.58 | -0.19 | 1418 |
| **ALSPAC-G1** | Life satisfaction | T1 | Prior mental ill-health | prior MH | 0.00 | 0.21 | -0.41 | 0.40 | 278 |
| **ALSPAC-G1** | Life satisfaction | T2 | Prior mental ill-health | no prior MH | -0.14 | 0.11 | -0.36 | 0.07 | 1123 |
| **ALSPAC-G1** | Life satisfaction | T2 | Prior mental ill-health | prior MH | 0.23 | 0.22 | -0.21 | 0.67 | 228 |
| **ALSPAC-G1** | Life satisfaction | T3 | Prior mental ill-health | no prior MH | -0.23 | 0.10 | -0.42 | -0.03 | 1336 |
| **ALSPAC-G1** | Life satisfaction | T3 | Prior mental ill-health | prior MH | -0.11 | 0.20 | -0.51 | 0.29 | 264 |
| **ALSPAC-G0** | Psych distress | T0 | Sex | male | 0.08 | 0.14 | -0.19 | 0.35 | 755 |
| **ALSPAC-G0** | Psych distress | T0 | Sex | female | 0.25 | 0.08 | 0.09 | 0.42 | 1947 |
| **ALSPAC-G0** | Psych distress | T1 | Sex | male | 0.31 | 0.11 | 0.10 | 0.53 | 843 |
| **ALSPAC-G0** | Psych distress | T1 | Sex | female | 0.54 | 0.08 | 0.38 | 0.70 | 2334 |
| **ALSPAC-G0** | Psych distress | T2 | Sex | male | 0.38 | 0.11 | 0.16 | 0.61 | 743 |
| **ALSPAC-G0** | Psych distress | T2 | Sex | female | 0.48 | 0.08 | 0.32 | 0.65 | 2003 |
| **ALSPAC-G0** | Psych distress | T3 | Sex | male | 0.31 | 0.12 | 0.08 | 0.54 | 739 |
| **ALSPAC-G0** | Psych distress | T3 | Sex | female | 0.37 | 0.08 | 0.20 | 0.53 | 2069 |
| **ALSPAC-G0** | Psych distress | T0 | Prior mental ill-health | no prior MH | 0.02 | 0.05 | -0.09 | 0.12 | 2223 |
| **ALSPAC-G0** | Psych distress | T0 | Prior mental ill-health | prior MH | 0.33 | 0.09 | 0.15 | 0.50 | 479 |
| **ALSPAC-G0** | Psych distress | T1 | Prior mental ill-health | no prior MH | 0.38 | 0.07 | 0.25 | 0.51 | 2121 |
| **ALSPAC-G0** | Psych distress | T1 | Prior mental ill-health | prior MH | 0.61 | 0.21 | 0.19 | 1.03 | 458 |
| **ALSPAC-G0** | Psych distress | T2 | Prior mental ill-health | no prior MH | 0.34 | 0.07 | 0.21 | 0.48 | 1889 |
| **ALSPAC-G0** | Psych distress | T2 | Prior mental ill-health | prior MH | 0.78 | 0.22 | 0.34 | 1.22 | 390 |
| **ALSPAC-G0** | Psych distress | T3 | Prior mental ill-health | no prior MH | 0.21 | 0.07 | 0.07 | 0.34 | 1951 |
| **ALSPAC-G0** | Psych distress | T3 | Prior mental ill-health | prior MH | 0.55 | 0.21 | 0.13 | 0.97 | 398 |
| **ALSPAC-G0** | Psych distress | T0 | Age | 45-54 | 0.32 | 0.26 | -0.18 | 0.83 | 447 |
| **ALSPAC-G0** | Psych distress | T0 | Age | 55-64 | 0.30 | 0.08 | 0.13 | 0.46 | 1954 |
| **ALSPAC-G0** | Psych distress | T0 | Age | 65-74 | -0.29 | 0.15 | -0.59 | 0.01 | 292 |
| **ALSPAC-G0** | Psych distress | T0 | Age | 75+ | -0.91 | 1.92 | -6.23 | 4.41 | 9 |
| **ALSPAC-G0** | Psych distress | T1 | Age | 45-54 | 0.38 | 0.25 | -0.11 | 0.88 | 576 |
| **ALSPAC-G0** | Psych distress | T1 | Age | 55-64 | 0.51 | 0.07 | 0.36 | 0.66 | 2250 |
| **ALSPAC-G0** | Psych distress | T1 | Age | 65-74 | 0.42 | 0.14 | 0.15 | 0.69 | 342 |
| **ALSPAC-G0** | Psych distress | T1 | Age | 75+ | -0.55 | 0.79 | -2.73 | 1.64 | 9 |
| **ALSPAC-G0** | Psych distress | T2 | Age | 45-54 | 0.52 | 0.26 | 0.01 | 1.03 | 442 |

| | | | | | | | | | |
|---|---|---|---|---|---|---|---|---|---|
| **ALSPAC-G0** | Psych distress | T2 | Age | 55-64 | 0.47 | 0.08 | 0.31 | 0.63 | 1976 |
| **ALSPAC-G0** | Psych distress | T2 | Age | 65-74 | 0.41 | 0.13 | 0.16 | 0.66 | 320 |
| **ALSPAC-G0** | Psych distress | T2 | Age | 75+ | -0.17 | 0.72 | -2.45 | 2.11 | 8 |
| **ALSPAC-G0** | Psych distress | T3 | Age | 45-54 | 0.48 | 0.31 | -0.13 | 1.09 | 368 |
| **ALSPAC-G0** | Psych distress | T3 | Age | 55-64 | 0.33 | 0.08 | 0.17 | 0.49 | 2054 |
| **ALSPAC-G0** | Psych distress | T3 | Age | 65-74 | 0.37 | 0.14 | 0.10 | 0.64 | 376 |
| **ALSPAC-G0** | Psych distress | T3 | Age | 75+ | -0.19 | 1.25 | -3.41 | 3.04 | 10 |

**Note: TP = timepoint; coef = coefficient; se = standard error ; l_ci = lower 95% confidence interval ; u_ci = cupper 95% confidence interval. Models adjust for covariates.**

#### Table S7. Regression coefficients from sensitivity analysis (excluding those living in households of larger than ten)

| cohort | outcome | exposure | coef | coef_se | lower_ci | upper_ci | n |
|---|---|---|---|---|---|---|---|
| **NCDS** | T1 Psych distress | T1 living alone alternative coding | 0.07 | 0.05 | -0.03 | 0.16 | 3566 |
| **NCDS** | T2 Psych distress | T1 living alone alternative coding | 0.04 | 0.04 | -0.04 | 0.12 | 3691 |
| **NCDS** | T3 Psych distress | T1 living alone alternative coding | 0.01 | 0.04 | -0.07 | 0.09 | 3723 |
| **NCDS** | T2 Psych distress | T2 living alone | 0.07 | 0.04 | -0.01 | 0.15 | 3682 |
| **NCDS** | T3 Psych distress | T3 living alone | 0.07 | 0.04 | -0.01 | 0.15 | 3719 |
| **NCDS** | T2 Life satisfaction | T2 living alone | -0.30 | 0.04 | -0.38 | -0.21 | 3685 |
| **NCDS** | T3 Life satisfaction | T3 living alone | -0.29 | 0.05 | -0.38 | -0.20 | 3727 |
| **NCDS** | T2 Psych distress (binary) | T2 living alone | 0.27 | 0.12 | 0.04 | 0.50 | 3681 |
| **NCDS** | T3 Psych distress (binary) | T3 living alone | 0.14 | 0.12 | -0.10 | 0.39 | 3716 |
| **BCS** | T1 Psych distress | T1 living alone alternative coding | -0.04 | 0.07 | -0.18 | 0.09 | 2616 |
| **BCS** | T2 Psych distress | T1 living alone alternative coding | 0.04 | 0.06 | -0.08 | 0.17 | 2673 |
| **BCS** | T3 Psych distress | T1 living alone alternative coding | 0.11 | 0.06 | -0.01 | 0.24 | 2725 |
| **BCS** | T2 Psych distress | T2 living alone | 0.05 | 0.07 | -0.08 | 0.17 | 2669 |
| **BCS** | T3 Psych distress | T3 living alone | 0.07 | 0.06 | -0.05 | 0.19 | 2720 |
| **BCS** | T2 Life satisfaction | T2 living alone | -0.35 | 0.06 | -0.47 | -0.24 | 2674 |
| **BCS** | T3 Life satisfaction | T3 living alone | -0.19 | 0.06 | -0.31 | -0.07 | 2726 |
| **BCS** | T2 Psych distress (binary) | T2 living alone | 0.04 | 0.16 | -0.27 | 0.35 | 2668 |
| **BCS** | T3 Psych distress (binary) | T3 living alone | 0.20 | 0.16 | -0.10 | 0.51 | 2719 |
| **NS** | T1 Psych distress | T1 living alone alternative coding | 0.36 | 0.13 | 0.10 | 0.62 | 1131 |
| **NS** | T2 Psych distress | T1 living alone alternative coding | 0.16 | 0.09 | -0.02 | 0.35 | 1178 |
| **NS** | T3 Psych distress | T1 living alone alternative coding | 0.08 | 0.09 | -0.10 | 0.27 | 1244 |
| **NS** | T2 Psych distress | T2 living alone | 0.09 | 0.10 | -0.10 | 0.28 | 1182 |
| **NS** | T3 Psych distress | T3 living alone | -0.08 | 0.09 | -0.24 | 0.09 | 1245 |
| **NS** | T2 Life satisfaction | T2 living alone | -0.12 | 0.06 | -0.25 | 0.00 | 1186 |
| **NS** | T3 Life satisfaction | T3 living alone | -0.01 | 0.07 | -0.15 | 0.13 | 1250 |
| **NS** | T2 Psych distress (binary) | T2 living alone | 0.03 | 0.21 | -0.38 | 0.44 | 1182 |
| **NS** | T3 Psych distress (binary) | T3 living alone | 0.29 | 0.20 | -0.09 | 0.68 | 1248 |

| | | | | | | | |
|---|---|---|---|---|---|---|---|
| **ELSA** | T1 Psych distress | T1 living alone alternative coding | 0.00 | 0.05 | -0.09 | 0.10 | 5470 |
| **ELSA** | T3 Psych distress | T1 living alone alternative coding | -0.04 | 0.05 | -0.13 | 0.06 | 5046 |
| **ELSA** | T3 Psych distress | T3 living alone | 0.21 | 0.04 | 0.12 | 0.29 | 5045 |
| **ELSA** | T3 Life satisfaction | T3 living alone | -0.17 | 0.04 | -0.26 | -0.09 | 5037 |
| **ELSA** | T3 Psych distress (binary) | T3 living alone | 0.09 | 0.02 | 0.05 | 0.12 | 5045 |
| **USOC** | T1 Psych distress | T1 living alone alternative coding | 0.17 | 0.03 | 0.12 | 0.23 | 12,087 |
| **USOC** | T2 Psych distress | T1 living alone alternative coding | 0.15 | 0.03 | 0.09 | 0.21 | 9,904 |
| **USOC** | T3 Psych distress | T1 living alone alternative coding | 0.12 | 0.03 | 0.06 | 0.18 | 9,912 |
| **USOC** | T1 Life satisfaction | T1 living alone alternative coding | -0.26 | 0.04 | -0.35 | -0.18 | 10,132 |
| **USOC** | T2 Life satisfaction | T1 living alone alternative coding | -0.17 | 0.03 | -0.22 | -0.11 | 9,904 |
| **USOC** | T3 life satisfaction (binary) | T1 living alone alternative coding | -0.20 | 0.03 | -0.25 | -0.14 | 9,912 |
| **USOC** | T2 Psych distress | T2 living alone | 0.12 | 0.03 | 0.07 | 0.17 | 9905 |
| **USOC** | T3 Psych distress | T3 living alone | 0.10 | 0.03 | 0.04 | 0.15 | 9913 |
| **USOC** | T2 Life satisfaction | T2 living alone | -0.17 | 0.03 | -0.23 | -0.12 | 9905 |
| **USOC** | T3 Life satisfaction | T3 living alone | -0.16 | 0.03 | -0.21 | -0.11 | 9913 |
| **USOC** | T2 Psych distress (binary) | T2 living alone | 0.30 | 0.05 | 0.19 | 0.40 | 9905 |
| **USOC** | T3 Psych distress (binary) | T3 living alone | 0.19 | 0.05 | 0.10 | 0.28 | 9913 |
| **USOC** | T2 life satisfaction (binary) | T2 living alone | 0.25 | 0.04 | 0.18 | 0.33 | 9905 |
| **USOC** | T3 life satisfaction (binary) | T3 living alone | 0.22 | 0.03 | 0.16 | 0.29 | 9913 |
| **ALSPAC-G1** | T1 Psych distress | T1 living alone alternative coding | 0.27 | 0.08 | 0.10 | 0.43 | 2062 |
| **ALSPAC-G1** | T2 Psych distress | T1 living alone alternative coding | 0.18 | 0.09 | -0.01 | 0.36 | 1609 |
| **ALSPAC-G1** | T3 Psych distress | T1 living alone alternative coding | 0.26 | 0.09 | 0.09 | 0.44 | 1917 |
| **ALSPAC-G1** | T2 Psych distress | T2 living alone | 0.15 | 0.09 | -0.02 | 0.33 | 1618 |
| **ALSPAC-G1** | T3 Psych distress | T3 living alone | 0.20 | 0.07 | 0.05 | 0.35 | 1964 |
| **ALSPAC-G1** | T2 Life satisfaction | T2 living alone | -0.16 | 0.09 | -0.33 | 0.02 | 1624 |
| **ALSPAC-G1** | T3 Life satisfaction | T3 living alone | -0.24 | 0.08 | -0.39 | -0.10 | 1965 |

| | | | | | | | |
|---|---|---|---|---|---|---|---|
| **ALSPAC-G0** | T1 Psych distress | T1 living alone alternative coding | 0.48 | 0.07 | 0.35 | 0.61 | 3175 |
| **ALSPAC-G0** | T2 Psych distress | T1 living alone alternative coding | 0.45 | 0.07 | 0.32 | 0.59 | 2744 |
| **ALSPAC-G0** | T3 Psych distress | T1 living alone alternative coding | 0.35 | 0.07 | 0.21 | 0.48 | 2806 |
| **ALSPAC-G0** | T2 Psych distress | T2 living alone | 0.46 | 0.07 | 0.33 | 0.59 | 2797 |
| **ALSPAC-G0** | T3 Psych distress | T3 living alone | 0.38 | 0.06 | 0.26 | 0.50 | 2873 |

**Note: TP = timepoint; coef = coefficient; se = standard error ; l_ci = lower 95% confidence interval ; u_ci = cupper 95% confidence interval. Models adjust for covariates.**

### Table S8. Results from longitudinal multi-level models (time x lone household interactions)

| cohort | outcome (cont outcomes only) | Model | exposure | TP | coef_interaction | se | l_ci | u_ci | main effect (time) | se | l_ci | u_ci | main effect (Lone HH) | se | L_ci | u_ci | n | |
|---|---|---|---|---|---|---|---|---|---|---|---|---|---|---|---|---|---|---|
| **NCDS** | Psych distress | Unadjusted | living alone | 1 | -0.04 | 0.05 | -0.14 | 0.06 | -0.03 | 0.02 | -0.07 | 0.00 | 0.14 | 0.05 | 0.04 | 0.23 | 3928 | |
| **NCDS** | Psych distress | Unadjusted | living alone | 2 | -0.07 | 0.05 | -0.17 | 0.03 | 0.10 | 0.02 | 0.07 | 0.14 | -- | -- | -- | -- | -- | |
| **NCDS** | Psych distress | Unadjusted | living alone | 3 | -0.10 | 0.05 | -0.21 | 0.00 | 0.07 | 0.02 | 0.04 | 0.11 | -- | -- | -- | -- | -- | |
| **NCDS** | Psych distress | Adjusted | living alone | 1 | -0.03 | 0.05 | -0.13 | 0.07 | -0.03 | 0.02 | -0.06 | 0.01 | 0.09 | 0.05 | -0.01 | 0.18 | 3928 | |
| **NCDS** | Psych distress | Adjusted | living alone | 2 | -0.06 | 0.05 | -0.16 | 0.04 | 0.11 | 0.02 | 0.07 | 0.14 | -- | -- | -- | -- | -- | |
| **NCDS** | Psych distress | Adjusted | living alone | 3 | -0.09 | 0.05 | -0.20 | 0.01 | 0.07 | 0.02 | 0.04 | 0.11 | -- | -- | -- | -- | -- | |
| **BCS** | Psych distress | Unadjusted | living alone | 1 | -0.05 | 0.06 | -0.16 | 0.06 | 0.02 | 0.02 | -0.01 | 0.05 | 0.06 | 0.06 | -0.05 | 0.18 | 3026 | |
| **BCS** | Psych distress | Unadjusted | living alone | 2 | -0.01 | 0.06 | -0.12 | 0.11 | 0.16 | 0.02 | 0.13 | 0.20 | -- | -- | -- | -- | -- | |
| **BCS** | Psych distress | Unadjusted | living alone | 3 | 0.06 | 0.06 | -0.06 | 0.18 | 0.09 | 0.02 | 0.05 | 0.12 | -- | -- | -- | -- | -- | |
| **BCS** | Psych distress | Adjusted | living alone | 1 | -0.06 | 0.06 | -0.17 | 0.06 | 0.02 | 0.02 | -0.01 | 0.05 | 0.03 | 0.06 | -0.08 | 0.15 | 3026 | |
| **BCS** | Psych distress | Adjusted | living alone | 2 | -0.02 | 0.06 | -0.13 | 0.10 | 0.17 | 0.02 | 0.13 | 0.20 | -- | -- | -- | -- | -- | |
| **BCS** | Psych distress | Adjusted | living alone | 3 | 0.05 | 0.06 | -0.07 | 0.17 | 0.09 | 0.02 | 0.05 | 0.12 | -- | -- | -- | -- | -- | |
| **Next Steps** | Psych distress | Unadjusted | living alone | 1 | 0.21 | 0.13 | -0.04 | 0.47 | 0.32 | 0.04 | 0.24 | 0.40 | 0.01 | 0.09 | -0.16 | 0.17 | 4555 | |
| **Next Steps** | Psych distress | Unadjusted | living alone | 2 | 0.14 | 0.12 | -0.10 | 0.37 | 0.28 | 0.04 | 0.20 | 0.36 | -- | -- | -- | -- | -- | |
| **Next Steps** | Psych distress | Unadjusted | living alone | 3 | 0.09 | 0.12 | -0.14 | 0.32 | 0.36 | 0.04 | 0.29 | 0.44 | -- | -- | -- | -- | -- | |
| **Next Steps** | Psych distress | Adjusted | living alone | 1 | 0.19 | 0.12 | -0.05 | 0.43 | 0.34 | 0.04 | 0.27 | 0.42 | 0.06 | 0.09 | -0.12 | 0.24 | 4555 | |
| **Next Steps** | Psych distress | Adjusted | living alone | 2 | 0.05 | 0.11 | -0.16 | 0.26 | 0.27 | 0.04 | 0.19 | 0.34 | -- | -- | -- | -- | -- | |
| **Next Steps** | Psych distress | Adjusted | living alone | 3 | 0.08 | 0.11 | -0.14 | 0.29 | 0.33 | 0.04 | 0.26 | 0.40 | -- | -- | -- | -- | -- | |
| | | | | | | | | | | | | | | | | | *(obs)* | *(groups)* |
| **ELSA** | Psych distress | Unadjusted | living alone | 1 | -0.03 | 0.06 | -0.15 | 0.08 | -0.01 | 0.02 | -0.05 | 0.03 | 0.34 | 0.05 | 0.25 | 0.43 | 15139 | 5047 |
| **ELSA** | Psych distress | Unadjusted | living alone | 3 | -0.10 | 0.05 | -0.21 | 0.00 | 0.02 | 0.02 | -0.02 | 0.07 | -- | -- | -- | -- | -- | |
| **ELSA** | Psych distress | Adjusted | living alone | 1 | -0.03 | 0.06 | -0.14 | 0.08 | 0.00 | 0.02 | -0.05 | 0.04 | 0.26 | 0.04 | 0.17 | 0.34 | 15139 | 5047 |
| **ELSA** | Psych distress | Adjusted | living alone | 3 | -0.10 | 0.05 | -0.20 | 0.00 | 0.03 | 0.02 | -0.01 | 0.07 | -- | -- | -- | -- | -- | |
| **TwinsUK** | Psych distress | Unadjusted | Live Alone | 1 | -0.08 | 0.04 | -0.16 | 0.00 | 0.24 | 0.02 | 0.21 | 0.28 | 0.08 | 0.05 | -0.01 | 0.18 | 2327 | |
| **TwinsUK** | Psych distress | Unadjusted | Live Alone | 2 | -0.10 | 0.04 | -0.19 | -0.02 | 0.49 | 0.02 | 0.45 | 0.52 | -- | -- | -- | -- | -- | |

| | | | | | | | | | | | | | | | | | |
|---|---|---|---|---|---|---|---|---|---|---|---|---|---|---|---|---|---|
| **TwinsUK** | Psych distress | Unadjusted | Live Alone | 3 | -0.11 | 0.04 | -0.19 | -0.03 | 0.54 | 0.02 | 0.50 | 0.57 | | | | | 2327 |
| **TwinsUK** | Psych distress | Adjusted | Live Alone | 1 | -0.08 | 0.04 | -0.16 | 0.00 | 0.24 | 0.02 | 0.21 | 0.28 | 0.10 | 0.05 | 0.01 | 0.20 | 2327 |
| **TwinsUK** | Psych distress | Adjusted | Live Alone | 2 | -0.10 | 0.04 | -0.19 | -0.02 | 0.49 | 0.02 | 0.45 | 0.52 | -- | -- | -- | -- | -- |
| **TwinsUK** | Psych distress | Adjusted | Live Alone | 3 | -0.11 | 0.04 | -0.19 | -0.03 | 0.54 | 0.02 | 0.50 | 0.57 | | | | | 2327 |
| **GS** | Psych distress | Unadjusted | Live Alone | 1 | 0.02 | 0.06 | -0.08 | 0.11 | 0.31 | 0.02 | 0.27 | 0.35 | 0.12 | 0.04 | 0.03 | 0.22 | 2882 |
| **GS** | Psych distress | Unadjusted | Live Alone | 2 | 0.11 | 0.06 | 0.00 | 0.21 | 0.22 | 0.02 | 0.18 | 0.26 | -- | -- | -- | -- | -- |
| **GS** | Psych distress | Unadjusted | Live Alone | 3 | 0.05 | 0.07 | -0.06 | 0.16 | 0.47 | 0.02 | 0.42 | 0.51 | -- | -- | -- | -- | -- |
| **GS** | Psych distress | Adjusted | Live Alone | 1 | 0.02 | 0.06 | -0.08 | 0.11 | 0.31 | 0.02 | 0.27 | 0.35 | 0.12 | 0.04 | 0.02 | 0.21 | 2882 |
| **GS** | Psych distress | Adjusted | Live Alone | 2 | 0.11 | 0.06 | 0.01 | 0.22 | 0.22 | 0.02 | 0.18 | 0.27 | -- | -- | -- | -- | -- |
| **GS** | Psych distress | Adjusted | Live Alone | 3 | 0.05 | 0.07 | -0.06 | 0.15 | 0.47 | 0.02 | 0.43 | 0.51 | -- | -- | -- | -- | -- |
| **USOC** | Psych distress | Unadjusted | living alone | 1 | 0.05 | 0.04 | -0.03 | 0.12 | 0.20 | 0.01 | 0.18 | 0.23 | 0.04 | 0.04 | -0.03 | 0.11 | 12270 |
| **USOC** | Psych distress | Unadjusted | living alone | 2 | 0.05 | 0.04 | -0.03 | 0.12 | 0.10 | 0.01 | 0.08 | 0.12 | -- | -- | -- | -- | -- |
| **USOC** | Psych distress | Unadjusted | living alone | 3 | 0.00 | 0.04 | -0.07 | 0.08 | 0.26 | 0.01 | 0.23 | 0.28 | -- | -- | -- | -- | -- |
| **USOC** | Psych distress | Adjusted | living alone | 1 | 0.05 | 0.04 | -0.02 | 0.12 | 0.20 | 0.01 | 0.18 | 0.23 | 0.04 | 0.04 | -0.02 | 0.11 | 12270 |
| **USOC** | Psych distress | Adjusted | living alone | 2 | 0.05 | 0.04 | -0.02 | 0.12 | 0.10 | 0.01 | 0.08 | 0.13 | -- | -- | -- | -- | -- |
| **USOC** | Psych distress | Adjusted | living alone | 3 | 0.01 | 0.04 | -0.07 | 0.08 | 0.26 | 0.01 | 0.24 | 0.29 | -- | -- | -- | -- | -- |
| **ALSPAC-G1** | Psych distress | Unadjusted | living alone | 1 | 0.04 | 0.08 | -0.12 | 0.19 | -0.10 | 0.02 | -0.14 | -0.07 | 0.26 | 0.09 | 0.08 | 0.44 | 2852 |
| **ALSPAC-G1** | Psych distress | Unadjusted | living alone | 2 | -0.04 | 0.07 | -0.18 | 0.11 | -0.06 | 0.02 | -0.10 | -0.02 | -- | -- | -- | -- | -- |
| **ALSPAC-G1** | Psych distress | Unadjusted | living alone | 3 | 0.04 | 0.08 | -0.12 | 0.20 | -0.06 | 0.02 | -0.10 | -0.01 | -- | -- | -- | -- | -- |
| **ALSPAC-G1** | Psych distress | Adjusted | living alone | 1 | 0.04 | 0.08 | -0.12 | 0.21 | -0.11 | 0.02 | -0.15 | -0.06 | 0.21 | 0.09 | 0.03 | 0.39 | 2102 |
| **ALSPAC-G1** | Psych distress | Adjusted | living alone | 2 | -0.02 | 0.08 | -0.18 | 0.14 | -0.07 | 0.02 | -0.12 | -0.02 | -- | -- | -- | -- | -- |
| **ALSPAC-G1** | Psych distress | Adjusted | living alone | 3 | 0.08 | 0.09 | -0.10 | 0.25 | -0.07 | 0.02 | -0.11 | -0.02 | -- | -- | -- | -- | -- |
| **ALSPAC-G0** | Psych distress | Unadjusted | living alone | 1 | 0.15 | 0.09 | -0.03 | 0.32 | -0.73 | 0.02 | -0.77 | -0.69 | 0.27 | 0.09 | 0.09 | 0.45 | 3434 |
| **ALSPAC-G0** | Psych distress | Unadjusted | living alone | 2 | 0.17 | 0.09 | -0.01 | 0.34 | -0.68 | 0.02 | -0.72 | -0.64 | -- | -- | -- | -- | -- |
| **ALSPAC-G0** | Psych distress | Unadjusted | living alone | 3 | 0.08 | 0.09 | -0.08 | 0.25 | -0.64 | 0.02 | -0.68 | -0.60 | -- | -- | -- | -- | -- |
| **ALSPAC-G0** | Psych distress | Adjusted | living alone | 1 | 0.13 | 0.09 | -0.05 | 0.31 | -0.73 | 0.02 | -0.77 | -0.69 | 0.23 | 0.09 | 0.05 | 0.41 | 3326 |
| **ALSPAC-G0** | Psych distress | Adjusted | living alone | 2 | 0.16 | 0.09 | -0.02 | 0.34 | -0.69 | 0.02 | -0.73 | -0.65 | -- | -- | -- | -- | -- |
| **ALSPAC-G0** | Psych distress | Adjusted | living alone | 3 | 0.09 | 0.09 | -0.08 | 0.26 | -0.65 | 0.02 | -0.69 | -0.61 | -- | -- | -- | -- | -- |

#### Table S9. Results from the meta-analysis in LPS

| | Time point | SMD | Lower CI | Upper CI | $I^2$ |
|---|---|---|---|---|---|
| Continuous Psychological Distress | 0 | 0.093 | 0.044 | 0.143 | 58.54 |
| | 1 | 0.191 | 0.092 | 0.289 | 87.21 |
| | 2 | 0.153 | 0.06 | 0.246 | 82.77 |
| | 3 | 0.146 | 0.074 | 0.218 | 76.04 |
| Binary Psychological Distress | 0 | 1.251 | 1.116 | 1.386 | 85.39 |
| | 1 | 1.312 | 1.03 | 1.593 | 96.75 |
| | 2 | 1.262 | 1.008 | 1.516 | 95.84 |
| | 3 | 1.307 | 1.148 | 1.466 | 91.89 |
| Continuous Life Satisfaction | 0 | -0.251 | -0.458 | -0.044 | 97.85 |
| | 1 | -0.223 | -0.296 | -0.15 | 66.26 |
| | 2 | -0.222 | -0.29 | -0.155 | 66.61 |
| | 3 | -0.191 | -0.236 | -0.146 | 35.38 |
| Binary Life Satisfaction | 0 | 1.056 | 0.779 | 1.333 | 97.76 |
| | 1 | 1.04 | 0.882 | 1.198 | 93.5 |
| | 2 | 1.05 | 0.806 | 1.293 | 97.48 |
| | 3 | 1.071 | 0.906 | 1.236 | 95.18 |

### Table S10. Main effects from EHR analyses

| | Anxiety | | | | Depression | | | | Eating disorder | | | | OCD | | | | Self harm | | | | Severe mental illness | | | |
|---|---|---|---|---|---|---|---|---|---|---|---|---|---|---|---|---|---|---|---|---|---|---|---|---|
| | Coefficient | SE | 95% CI | P value | Coefficient | SE | 95% CI | P value | Coefficient | SE | 95% CI | P value | Coefficient | SE | 95% CI | P value | Coefficient | SE | 95% CI | P value | Coefficient | SE | 95% CI | P value |
| **Effect of not living alone pre-pandemic** | -13.63 | 3.67 | (-20.93, -6.33) | <0.001 | -25.56 | 3.89 | (-33.30, -17.82) | <0.001 | -2.88 | 0.19 | (-3.27, -2.49) | <0.001 | -0.31 | 0.11 | (-0.53, -0.08) | 0.009 | -25.83 | 0.85 | (-27.53, -24.14) | <0.001 | -57.96 | 1.97 | (-61.87, -54.06) | <0.001 |
| **Effect of the pandemic in those living alone** | -48.07 | 5.06 | (-58.12,-38.02) | <0.001 | -66.12 | 4.96 | (-75.99,-56.25) | <0.001 | 0.1 | 0.4 | (-0.70,0.91) | 0.798 | -0.61 | 0.14 | (-0.89,-0.34) | <0.001 | -6.99 | 1.44 | (-9.85,-4.13) | <0.001 | -7.48 | 2.79 | (-13.03,-1.92) | 0.009 |
| **Effect of the pandemic among those not living** | 5.67 | 7.55 | (-9.34, 20.68) | 0.455 | 13.42 | 6.89 | (-0.29, 27.12) | 0.055 | 0.49 | 0.52 | (-0.55, 1.52) | 0.352 | 0.05 | 0.17 | (-0.30, 0.39) | 0.795 | 7.95 | 1.68 | (4.62, 11.29) | <0.001 | 4.40 | 3.00 | (-1.57, 10.37) | 0.147 |
| **Summer** | 7.06 | 6.01 | (-4.89, 19.00) | 0.244 | 8.81 | 6.05 | (-3.22, 20.83) | 0.149 | 0.43 | 0.4 | (-0.37, 1.22) | 0.291 | -0.01 | 0.14 | (-0.28, 0.27) | 0.966 | 3.02 | 1.06 | (0.93, 5.12) | 0.005 | 0.33 | 1.97 | (-3.57, 4.24) | 0.866 |
| **Autumn** | 22.19 | 6.11 | (10.05, 34.33) | <0.001 | 24.78 | 6.38 | (12.09, 37.47) | <0.001 | 0.25 | 0.38 | (-0.51, 1.00) | 0.521 | 0.06 | 0.14 | (-0.22, 0.34) | 0.654 | -0.36 | 0.95 | (-2.24, 1.53) | 0.708 | 2.49 | 1.92 | (-1.33, 6.32) | 0.199 |
| **Winter** | 1.97 | 6.83 | (-11.60, 15.55) | 0.773 | 8.36 | 6.5 | (-4.55, 21.27) | 0.201 | 0.05 | 0.4 | (-0.75, 0.85) | 0.906 | 0.00 | 0.14 | (-0.27, 0.28) | 0.989 | -2.08 | 1.14 | (-4.34, 0.18) | 0.071 | 3.74 | 2.54 | (-1.31, 8.80) | 0.144 |
| **Constant** | 244.48 | 5.08 | (234.39, 254.57) | <0.001 | 212.59 | 5.53 | (201.60, 223.58) | <0.001 | 9.49 | 0.29 | (8.91, 10.06) | <0.001 | 3.25 | 0.12 | (3.01, 3.49) | <0.001 | 52.32 | 0.94 | (50.46, 54.18) | <0.001 | 88.36 | 2.1 | (84.19, 92.53) | <0.001 |

### Table S11. Sex-stratified estimates from EHR analyses

| | Anxiety | | | | Depression | | | | Eating disorder | | | | OCD | | | | Self harm | | | | Severe mental illness | | | |
|---|---|---|---|---|---|---|---|---|---|---|---|---|---|---|---|---|---|---|---|---|---|---|---|---|
| | Coefficient | SE | 95% CI | P value | Coefficient | SE | 95% CI | P value | Coefficient | SE | 95% CI | P value | Coefficient | SE | 95% CI | P value | Coefficient | SE | 95% CI | P value | Coefficient | SE | 95% CI | P value |
| **Pre-pandemic effects** | | | | | | | | | | | | | | | | | | | | | | | | |
| Females not living alone | 16.4 | 4.48 | (7.56, 25.24) | <0.001 | 2.72 | 4.75 | (-6.67, 12.10) | 0.568 | -4.24 | 0.36 | (-4.94, -3.53) | <0.001 | 0.24 | 0.14 | (-0.02, 0.51) | 0.074 | -26.29 | 1.02 | (-28.31, -24.27) | <0.001 | -49.45 | 1.98 | (-53.35, -45.56) | <0.001 |
| Males living alone | -88.63 | 3.8 | (-96.12, -81.14) | <0.001 | -34.16 | 4.51 | (-43.05, -25.26) | <0.001 | -12.88 | 0.34 | (-13.54, -12.21) | <0.001 | -0.18 | 0.16 | (-0.50, 0.14) | 0.263 | -11.17 | 0.99 | (-13.12, -9.22) | <0.001 | 8.64 | 2.61 | (3.49, 13.80) | 0.001 |
| Males not living alone | -135.15 | 3.47 | (-142.01, -128.29 | <0.001 | -88.65 | 3.95 | (-96.45, -80.85) | <0.001 | -14.26 | 0.33 | (-14.92, -13.61) | <0.001 | -1.06 | 0.12 | (-1.31, -0.82) | <0.001 | -36.64 | 0.98 | (-38.57, -34.71) | <0.001 | -58.29 | 2 | (-62.23, -54.34) | <0.001 |
| **Effect of the pandemic in those living a** | -37.89 | 6.08 | (-49.88,-25.89) | <0.001 | -65.09 | 5.45 | (-75.86,-54.33) | <0.001 | 0.6 | 0.7 | (-0.79,1.99) | 0.395 | -0.48 | 0.16 | (-0.79,-0.17) | 0.002 | -8.2 | 1.58 | (-11.33,-5.08) | <0.001 | -4.4 | 2.93 | (-10.18,1.38) | 0.134 |
| **Interaction effects during pandemic** | | | | | | | | | | | | | | | | | | | | | | | | |
| Females not living alone | -3.09 | 9.82 | (-22.48, 16.29) | 0.753 | 3.09 | 8.18 | (-13.05, 19.23) | 0.706 | 0.62 | 0.91 | (-1.19, 2.42) | 0.5 | -0.1 | 0.21 | (-0.52, 0.32) | 0.641 | 9.4 | 1.88 | (5.69, 13.11) | <0.001 | 0.79 | 3.17 | (-5.48, 7.06) | 0.804 |
| Males living alone | -8.43 | 7.52 | (-23.27, 6.41) | 0.264 | -2.23 | 6.98 | (-16.00, 11.54) | 0.75 | -0.97 | 0.71 | (-2.38, 0.43) | 0.173 | -0.27 | 0.22 | (-0.70, 0.15) | 0.209 | 2.47 | 2.11 | (-1.69, 6.63) | 0.243 | -6.38 | 3.94 | (-14.16, 1.40) | 0.107 |
| Males not living alone | 7.11 | 7.61 | (-7.91, 22.14) | 0.352 | 22.02 | 6.67 | (8.86, 35.18) | 0.001 | -0.62 | 0.7 | (-2.01, 0.77) | 0.379 | -0.06 | 0.17 | (-0.41, 0.28) | 0.717 | 8.91 | 1.75 | (5.46, 12.35) | <0.001 | 1.86 | 3.11 | (-4.28, 7.99) | 0.551 |
| **Summer** | 6.99 | 4.49 | (-1.87, 15.85) | 0.121 | 8.81 | 4.37 | (0.18, 17.44) | 0.045 | 0.42 | 0.36 | (-0.30, 1.14) | 0.249 | 0 | 0.11 | (-0.21, 0.20) | 0.963 | 3.04 | 0.77 | (1.52, 4.57) | <0.001 | 0.33 | 1.41 | (-2.44, 3.11) | 0.813 |
| **Autumn** | 23.85 | 4.59 | (14.79, 32.91) | <0.001 | 24.85 | 4.66 | (15.66, 34.04) | <0.001 | 0.24 | 0.35 | (-0.45, 0.93) | 0.489 | 0.07 | 0.11 | (-0.15, 0.29) | 0.536 | -0.34 | 0.71 | (-1.74, 1.07) | 0.638 | 2.5 | 1.37 | (-0.22, 5.21) | 0.071 |
| **Winter** | 3.44 | 5.08 | (-6.58, 13.46) | 0.499 | 8.45 | 4.68 | (-0.80, 17.69) | 0.073 | 0.06 | 0.37 | (-0.67, 0.79) | 0.878 | 0 | 0.11 | (-0.21, 0.21) | 0.987 | -2.05 | 0.84 | (-3.71, -0.40) | 0.015 | 3.76 | 1.8 | (0.21, 7.32) | 0.038 |
| **Constant** | 289.5 | 4.07 | (281.46, 297.54) | <0.001 | 229.65 | 4.77 | (220.24, 239.07) | <0.001 | 15.8 | 0.37 | (15.08, 16.53) | <0.001 | 3.34 | 0.12 | (3.10, 3.59) | <0.001 | 57.94 | 0.99 | (55.99, 59.89) | <0.001 | 84.27 | 1.98 | (80.36, 88.17) | <0.001 |

**NB: Except where specified, coefficients relate to effects in the baseline categories of all variables (pre-pandemic, spring, living alone, females)**

### Table S12. Age band-stratified estimates from EHR analyses

| | Anxiety | | | | Depression | | | | Eating disorder | | | | OCD | | | | Self harm | | | | Severe mental illness | | | |
|---|---|---|---|---|---|---|---|---|---|---|---|---|---|---|---|---|---|---|---|---|---|---|---|---|
| | Coefficient | SE | 95% CI | P value | Coefficient | SE | 95% CI | P value | Coefficient | SE | 95% CI | P value | Coefficient | SE | 95% CI | P value | Coefficient | SE | 95% CI | P value | Coefficient | SE | 95% CI | P value |
| **Pre-pandemic effects** | | | | | | | | | | | | | | | | | | | | | | | | |
| Age 18-39 not living alone | -266.13 | 5.29 | (-276.55, -255.72 | <0.001 | -215.01 | 5.25 | (-225.34, -204.6 | <0.001 | -18.37 | 0.54 | (-19.42, -17.32) | <0.001 | -5.49 | 0.32 | (-6.12, -4.87) | <0.001 | -90.82 | 1.54 | (-93.85, -87.80) | <0.001 | 6.97 | 3.34 | (0.40, 13.53) | 0.038 |
| Age 40-59 living alone | -308.98 | 5.27 | (-319.34, -298.62 | <0.001 | -262.97 | 4.85 | (-272.51, -253.4 | <0.001 | -19.59 | 0.52 | (-20.61, -18.56) | <0.001 | -5.93 | 0.31 | (-6.54, -5.32) | <0.001 | -100.56 | 1.51 | (-103.52, -97.5 | <0.001 | -46.86 | 1.49 | (-49.80, -43.93) | <0.001 |
| Age 40-59 not living alone | -298.61 | 5.87 | (-310.14, -287.07 | <0.001 | -269.19 | 4.75 | (-278.54, -259.8 | <0.001 | -18.45 | 0.54 | (-19.50, -17.39) | <0.001 | -6.43 | 0.31 | (-7.05, -5.82) | <0.001 | -98.63 | 1.56 | (-101.70, -95.5 | <0.001 | -35.79 | 2.82 | (-41.35, -30.24) | <0.001 |
| Age 60-79 living alone | -318.99 | 5.92 | (-330.63, -307.36 | <0.001 | -282.19 | 4.94 | (-291.92, -272.4 | <0.001 | -18.11 | 0.54 | (-19.18, -17.05) | <0.001 | -6.4 | 0.31 | (-7.01, -5.78) | <0.001 | -101.43 | 1.5 | (-104.37, -98.4 | <0.001 | -49.06 | 1.67 | (-52.34, -45.77) | <0.001 |
| Age 60-79 not living alone | -58.01 | 6.8 | (-71.40, -44.63) | <0.001 | -59.24 | 5.69 | (-70.43, -48.06) | <0.001 | -9.22 | 0.55 | (-10.30, -8.14) | <0.001 | -1.22 | 0.32 | (-1.85, -0.58) | <0.001 | -58.58 | 1.65 | (-61.82, -55.34) | <0.001 | -45.75 | 1.33 | (-48.37, -43.12) | <0.001 |
| Age 80+ living alone | -136.01 | 5.86 | (-147.54, -124.47 | <0.001 | -69.54 | 7.08 | (-83.46, -55.62) | <0.001 | -12.9 | 0.55 | (-13.97, -11.82) | <0.001 | -2.85 | 0.33 | (-3.51, -2.20) | <0.001 | -40.07 | 1.74 | (-43.49, -36.65) | <0.001 | 55.06 | 2.72 | (49.70, 60.42) | <0.001 |
| Age 80+ not living alone | -209.38 | 5.33 | (-219.87, -198.89 | <0.001 | -160.58 | 5.26 | (-170.93, -150.2 | <0.001 | -17.62 | 0.52 | (-18.65, -16.60) | <0.001 | -4.77 | 0.31 | (-5.39, -4.16) | <0.001 | -86.08 | 1.53 | (-89.09, -83.07) | <0.001 | -42.59 | 1.45 | (-45.44, -39.74) | <0.001 |
| **Effect of the pandemic in those living alo** | -66.76 | 10.6 | (-87.61,-45.90) | <0.001 | -79.77 | 9.25 | (-97.96,-61.58) | <0.001 | 0.87 | 1.22 | (-1.53,3.27) | 0.477 | -0.65 | 0.44 | (-1.51,0.21) | 0.138 | -20.46 | 3.02 | (-26.40,-14.52) | <0.001 | -16.36 | 1.9 | (-20.10,-12.61) | <0.001 |
| **Interaction effects during pandemic** | | | | | | | | | | | | | | | | | | | | | | | | |
| Age 18-39 not living alone | 10.27 | 14.6 | (-18.44, 38.98) | 0.482 | 10.88 | 12.19 | (-13.10, 34.86) | 0.373 | 1.27 | 1.42 | (-1.51, 4.05) | 0.37 | -0.05 | 0.5 | (-1.03, 0.92) | 0.913 | 22.24 | 3.32 | (15.71, 28.77) | <0.001 | 13.12 | 2.2 | (8.81, 17.44) | <0.001 |
| Age 40-59 living alone | 5.44 | 11.82 | (-17.80, 28.67) | 0.646 | -23.08 | 11.41 | (-45.52, -0.63) | 0.044 | -1.47 | 1.25 | (-3.93, 0.99) | 0.242 | -0.69 | 0.46 | (-1.59, 0.21) | 0.133 | 12.26 | 3.66 | (5.06, 19.46) | 0.001 | -1.06 | 4.16 | (-9.25, 7.13) | 0.8 |
| Age 40-59 not living alone | 37.8 | 11.65 | (14.90, 60.70) | 0.001 | 24.54 | 10.38 | (4.12, 44.96) | 0.019 | -1.04 | 1.23 | (-3.46, 1.38) | 0.399 | 0.1 | 0.44 | (-0.77, 0.96) | 0.824 | 21.46 | 3.1 | (15.36, 27.55) | <0.001 | 11.83 | 2.3 | (7.31, 16.35) | <0.001 |
| Age 60-79 living alone | 48.69 | 11.06 | (26.93, 70.45) | <0.001 | 37.36 | 9.7 | (18.28, 56.44) | <0.001 | -0.8 | 1.23 | (-3.22, 1.62) | 0.516 | 0.45 | 0.44 | (-0.42, 1.33) | 0.308 | 21.47 | 3.09 | (15.39, 27.54) | <0.001 | 18.82 | 4.5 | (9.98, 27.66) | <0.001 |
| Age 60-79 not living alone | 59.89 | 11.2 | (37.87, 81.91) | <0.001 | 59.17 | 9.62 | (40.25, 78.08) | <0.001 | -0.96 | 1.22 | (-3.35, 1.43) | 0.43 | 0.44 | 0.44 | (-0.42, 1.30) | 0.315 | 21.81 | 3.08 | (15.76, 27.86) | <0.001 | 15.04 | 2.31 | (10.50, 19.58) | <0.001 |
| Age 80+ living alone | 58.36 | 11.22 | (36.28, 80.43) | <0.001 | 64.72 | 9.51 | (46.01, 83.42) | <0.001 | -0.85 | 1.23 | (-3.27, 1.56) | 0.488 | 0.64 | 0.44 | (-0.23, 1.50) | 0.148 | 23.92 | 3.13 | (17.76, 30.07) | <0.001 | 26.57 | 3.5 | (19.69, 33.45) | <0.001 |
| Age 80+ not living alone | 59.53 | 11.51 | (36.88, 82.17) | <0.001 | 70.83 | 9.63 | (51.89, 89.77) | <0.001 | -0.99 | 1.24 | (-3.42, 1.45) | 0.426 | 0.54 | 0.44 | (-0.32, 1.40) | 0.219 | 22.2 | 3.07 | (16.16, 28.24) | <0.001 | 17.56 | 2.38 | (12.87, 22.25) | <0.001 |
| **Summer** | 7.06 | 3.43 | (0.32, 13.81) | 0.04 | 8.47 | 3.24 | (2.09, 14.85) | 0.009 | 0.32 | 0.3 | (-0.26, 0.91) | 0.279 | 0.02 | 0.1 | (-0.19, 0.22) | 0.87 | 2.81 | 0.71 | (1.43, 4.20) | <0.001 | 0 | 1.12 | (-2.19, 2.20) | 0.999 |
| **Autumn** | 21.36 | 3.58 | (14.32, 28.40) | <0.001 | 21.67 | 3.59 | (14.60, 28.73) | <0.001 | 0.17 | 0.27 | (-0.37, 0.70) | 0.54 | 0.08 | 0.11 | (-0.13, 0.29) | 0.46 | -0.11 | 0.59 | (-1.27, 1.05) | 0.851 | 2.28 | 1.09 | (0.13, 4.43) | 0.038 |
| **Winter** | 3.57 | 3.88 | (-4.06, 11.20) | 0.359 | 7.49 | 3.53 | (0.55, 14.43) | 0.035 | 0.08 | 0.29 | (-0.48, 0.65) | 0.77 | 0.03 | 0.1 | (-0.16, 0.22) | 0.738 | -1.69 | 0.72 | (-3.11, -0.26) | 0.02 | 4 | 1.43 | (1.19, 6.81) | 0.005 |
| **Constant** | 409.92 | 5.24 | (399.61, 420.23) | <0.001 | 338.14 | 5.06 | (328.19, 348.08) | <0.001 | 21.52 | 0.51 | (20.51, 22.53) | <0.001 | 6.66 | 0.3 | (6.08, 7.24) | <0.001 | 105.9 | 1.48 | (103.00, 108.81 | <0.001 | 75.54 | 1.42 | (72.75, 78.34) | <0.001 |

**NB: Except where specified, coefficients relate to effects in the baseline categories of all variables (pre-pandemic, spring, living alone, age 18-40)**

### Table S13. Ethnicity-stratified estimates from EHR analyses

| | Anxiety | | | | Depression | | | | Eating disorder | | | | OCD | | | | Self harm | | | | Severe mental illness | | | |
|---|---|---|---|---|---|---|---|---|---|---|---|---|---|---|---|---|---|---|---|---|---|---|---|---|
| | Coefficien | SE | 95% CI | P value | Coefficien | SE | 95% CI | P value | Coefficien | SE | 95% CI | P value | Coefficien | SE | 95% CI | P value | Coefficien | SE | 95% CI | P value | Coefficien | SE | 95% CI | P value |
| **Pre-pandemic effects** | | | | | | | | | | | | | | | | | | | | | | | | |
| White not living alone | 85.28 | 9.86 | (65.91, 104.65) | <0.001 | 92.1 | 9.29 | (73.85, 110.34) | <0.001 | 7.25 | 1.36 | (4.59, 9.92) | <0.001 | 1.7 | 0.76 | (0.21, 3.19) | 0.026 | 27.63 | 2.84 | (22.05, 33.21) | <0.001 | 96.47 | 4.49 | (87.66, 105.29) | <0.001 |
| Mixed living alone | 46.66 | 4.4 | (38.02, 55.31) | <0.001 | 37.2 | 4.37 | (28.61, 45.79) | <0.001 | 2.42 | 0.76 | (0.93, 3.91) | 0.002 | 0.46 | 0.4 | (-0.34, 1.25) | 0.259 | 0.76 | 1.66 | (-2.50, 4.03) | 0.646 | -11.69 | 2.51 | (-16.62, -6.75) | <0.001 |
| Mixed not living alone | -15.34 | 3.99 | (-23.19, -7.50) | <0.001 | -5.22 | 4.11 | (-13.29, 2.86) | 0.205 | -3.29 | 0.45 | (-4.18, -2.40) | <0.001 | -0.08 | 0.3 | (-0.68, 0.52) | 0.798 | -12.81 | 1.15 | (-15.07, -10.56) | <0.001 | 36.7 | 2.88 | (31.05, 42.36) | <0.001 |
| Asian living alone | -61.52 | 3.09 | (-67.58, -55.46) | <0.001 | -63.63 | 2.94 | (-69.40, -57.85) | <0.001 | -4.31 | 0.32 | (-4.94, -3.68) | <0.001 | -0.33 | 0.24 | (-0.79, 0.14) | 0.165 | -25.01 | 0.74 | (-26.46, -23.55) | <0.001 | -22.64 | 2.15 | (-26.86, -18.42) | <0.001 |
| Asian not living alone | -19.49 | 3.84 | (-27.04, -11.94) | <0.001 | 0.64 | 6.44 | (-12.01, 13.30) | 0.921 | -1.65 | 0.59 | (-2.81, -0.49) | 0.005 | -1.01 | 0.38 | (-1.75, -0.26) | 0.008 | -7.61 | 1.43 | (-10.42, -4.80) | <0.001 | 116.59 | 3.83 | (109.06, 124.12 | <0.001 |
| Black living alone | -75.11 | 3.78 | (-82.54, -67.69) | <0.001 | -62.95 | 3.1 | (-69.04, -56.86) | <0.001 | -3.45 | 0.38 | (-4.20, -2.70) | <0.001 | -1.72 | 0.25 | (-2.22, -1.22) | <0.001 | -23.51 | 0.89 | (-25.25, -21.76) | <0.001 | -12.7 | 2.42 | (-17.46, -7.95) | <0.001 |
| Black not living alone | -50.57 | 4.29 | (-59.01, -42.14) | <0.001 | -36.62 | 3.89 | (-44.26, -28.98) | <0.001 | -3.14 | 0.59 | (-4.29, -1.98) | <0.001 | -0.62 | 0.33 | (-1.26, 0.03) | 0.063 | -15.21 | 1.13 | (-17.43, -12.98) | <0.001 | -3.84 | 3.46 | (-10.64, 2.96) | 0.268 |
| Other living alone | -73.71 | 3.49 | (-80.57, -66.85) | <0.001 | -71.15 | 2.92 | (-76.88, -65.41) | <0.001 | -4.58 | 0.43 | (-5.43, -3.73) | <0.001 | -1.31 | 0.27 | (-1.84, -0.79) | <0.001 | -24.23 | 0.85 | (-25.89, -22.56) | <0.001 | -36.5 | 2.14 | (-40.71, -32.30) | <0.001 |
| Other not living alone | 18.28 | 4.18 | (10.06, 26.50) | <0.001 | 5.06 | 3.04 | (-0.91, 11.02) | 0.096 | -1.09 | 0.31 | (-1.69, -0.49) | <0.001 | 0.48 | 0.22 | (0.05, 0.91) | 0.027 | -9.22 | 0.89 | (-10.98, -7.47) | <0.001 | -37.9 | 2.14 | (-42.11, -33.69) | <0.001 |
| Unknown living alone | 71.73 | 3.89 | (64.10, 79.36) | <0.001 | 63.81 | 4.25 | (55.46, 72.16) | <0.001 | 3.08 | 0.34 | (2.41, 3.75) | <0.001 | 0.76 | 0.24 | (0.30, 1.23) | 0.001 | 25 | 0.97 | (23.10, 26.91) | <0.001 | 34.07 | 2.83 | (28.51, 39.64) | <0.001 |
| Unknown not living alone | 57.48 | 3.92 | (49.78, 65.19) | <0.001 | 38.83 | 3.48 | (31.98, 45.67) | <0.001 | -0.17 | 0.3 | (-0.77, 0.42) | 0.571 | 0.31 | 0.22 | (-0.13, 0.75) | 0.163 | -5.5 | 0.82 | (-7.12, -3.88) | <0.001 | -27.15 | 2.18 | (-31.44, -22.85) | <0.001 |
| **Effect of the pandemic in those living alone and w** | -30.26 | 4.82 | (-39.73,-20.80) | <0.001 | -45.63 | 3.93 | (-53.35,-37.90) | <0.001 | 0.96 | 0.4 | (0.18,1.74) | 0.015 | -0.68 | 0.26 | (-1.19,-0.16) | 0.01 | -2.11 | 1.03 | (-4.14,-0.08) | 0.042 | -3.76 | 2.45 | (-8.56,1.04) | 0.125 |
| **Interaction effects during pandemic** | | | | | | | | | | | | | | | | | | | | | | | | |
| White not living alone | -0.56 | 8.31 | (-16.89, 15.78) | 0.947 | 1.3 | 6.43 | (-11.33, 13.93) | 0.84 | 0.6 | 0.63 | (-0.64, 1.84) | 0.34 | 0.12 | 0.3 | (-0.47, 0.70) | 0.692 | 4.24 | 1.37 | (1.56, 6.93) | 0.002 | 2.28 | 2.62 | (-2.88, 7.44) | 0.386 |
| Mixed living alone | -17.47 | 7.17 | (-31.57, -3.38) | 0.015 | -29.04 | 6.42 | (-41.64, -16.44) | <0.001 | -1.17 | 0.6 | (-2.36, 0.01) | 0.053 | 0.01 | 0.3 | (-0.58, 0.59) | 0.986 | -6.43 | 2.07 | (-10.50, -2.37) | 0.002 | -3.55 | 3.84 | (-11.11, 4.00) | 0.356 |
| Mixed not living alone | -9.99 | 8.06 | (-25.81, 5.84) | 0.216 | -13.29 | 6.47 | (-26.01, -0.57) | 0.041 | -0.54 | 0.51 | (-1.54, 0.46) | 0.289 | 0.06 | 0.29 | (-0.50, 0.62) | 0.83 | 2.81 | 1.32 | (0.21, 5.41) | 0.034 | 0.28 | 2.74 | (-5.10, 5.66) | 0.919 |
| Asian living alone | 21.59 | 13.06 | (-4.06, 47.25) | 0.099 | 10.57 | 12.9 | (-14.77, 35.91) | 0.413 | 2.59 | 2.11 | (-1.56, 6.74) | 0.221 | -0.46 | 0.95 | (-2.32, 1.40) | 0.627 | -8.46 | 3.54 | (-15.41, -1.51) | 0.017 | -23.3 | 7.59 | (-38.20, -8.39) | 0.002 |
| Asian not living alone | 1.49 | 9.48 | (-17.13, 20.11) | 0.875 | -1.01 | 8.65 | (-18.00, 15.98) | 0.907 | -0.02 | 1.11 | (-2.20, 2.16) | 0.986 | 0.7 | 0.57 | (-0.43, 1.82) | 0.225 | 4.46 | 2.55 | (-0.55, 9.47) | 0.081 | 0.56 | 3.4 | (-6.11, 7.23) | 0.868 |
| Black living alone | 13.67 | 7.7 | (-1.45, 28.80) | 0.076 | 6.49 | 6.61 | (-6.49, 19.46) | 0.326 | 0.23 | 0.67 | (-1.08, 1.54) | 0.729 | 1.29 | 0.46 | (0.39, 2.18) | 0.005 | 3 | 1.76 | (-0.45, 6.46) | 0.088 | -12.35 | 3.76 | (-19.73, -4.96) | 0.001 |
| Black not living alone | 18.97 | 5.97 | (7.25, 30.70) | 0.002 | 20.53 | 5.28 | (10.17, 30.90) | <0.001 | -0.95 | 0.47 | (-1.87, -0.02) | 0.044 | 0.28 | 0.3 | (-0.31, 0.88) | 0.351 | 3.33 | 1.21 | (0.96, 5.70) | 0.006 | -0.02 | 2.74 | (-5.41, 5.36) | 0.993 |
| Other living alone | 15.22 | 8.33 | (-1.14, 31.59) | 0.068 | 6.1 | 9.76 | (-13.07, 25.27) | 0.532 | -0.37 | 1.12 | (-2.57, 1.82) | 0.74 | 0.43 | 0.51 | (-0.58, 1.44) | 0.404 | -1.65 | 2.28 | (-6.13, 2.84) | 0.472 | -27.43 | 6.26 | (-39.74, -15.12) | <0.001 |
| Other not living alone | 24.12 | 6.28 | (11.78, 36.47) | <0.001 | 22.56 | 5.6 | (11.55, 33.56) | <0.001 | -1.16 | 0.63 | (-2.40, 0.09) | 0.068 | 0.4 | 0.31 | (-0.21, 1.01) | 0.201 | 2.58 | 1.39 | (-0.15, 5.30) | 0.064 | 0.15 | 3 | (-5.75, 6.05) | 0.96 |
| Unknown living alone | 22.9 | 7.58 | (8.00, 37.80) | 0.003 | 20.97 | 6.96 | (7.30, 34.64) | 0.003 | -0.66 | 0.81 | (-2.25, 0.93) | 0.417 | 0.38 | 0.46 | (-0.52, 1.29) | 0.405 | -1.53 | 1.73 | (-4.92, 1.86) | 0.375 | -4.34 | 4.21 | (-12.61, 3.92) | 0.303 |
| Unknown not living alone | 15.62 | 6.2 | (3.44, 27.80) | 0.012 | 25.13 | 5.03 | (15.25, 35.02) | <0.001 | -0.08 | 0.58 | (-1.22, 1.07) | 0.894 | 0.59 | 0.34 | (-0.07, 1.25) | 0.079 | 2.75 | 1.29 | (0.20, 5.29) | 0.034 | 1.56 | 2.69 | (-3.72, 6.84) | 0.561 |
| **Summer** | 3.07 | 2.78 | (-2.39, 8.53) | 0.27 | 9.11 | 2.95 | (3.31, 14.90) | 0.002 | 0.6 | 0.29 | (0.03, 1.18) | 0.04 | 0.01 | 0.15 | (-0.29, 0.31) | 0.956 | 3.14 | 0.64 | (1.88, 4.40) | <0.001 | 0.53 | 1.45 | (-2.31, 3.37) | 0.714 |
| **Autumn** | 13.92 | 2.75 | (8.51, 19.33) | <0.001 | 19.9 | 2.95 | (14.11, 25.69) | <0.001 | 0.13 | 0.3 | (-0.47, 0.73) | 0.676 | 0.22 | 0.16 | (-0.10, 0.54) | 0.17 | 0.05 | 0.62 | (-1.17, 1.27) | 0.937 | 2.28 | 1.41 | (-0.48, 5.05) | 0.105 |
| **Winter** | 1.86 | 3.03 | (-4.11, 7.82) | 0.541 | 11.49 | 3.23 | (5.15, 17.83) | <0.001 | 0.33 | 0.37 | (-0.40, 1.05) | 0.38 | 0.11 | 0.16 | (-0.20, 0.42) | 0.48 | -1.01 | 0.7 | (-2.38, 0.37) | 0.151 | 3.87 | 1.49 | (0.95, 6.79) | 0.009 |
| **Constant** | 198.33 | 3.16 | (192.12, 204.53) | <0.001 | 166.56 | 2.96 | (160.74, 172.38 | <0.001 | 7.29 | 0.34 | (6.62, 7.95) | <0.001 | 2.65 | 0.23 | (2.20, 3.10) | <0.001 | 34.4 | 0.73 | (32.96, 35.85) | <0.001 | 59.04 | 2.19 | (54.75, 63.33) | <0.001 |

**NB: Except where specified, coefficients relate to effects in the baseline categories of all variables (pre-pandemic, spring, living alone, White ethnicity)**

### Table S14. IMD-stratified estimates from EHR analyses

| | Anxiety | | | | Depression | | | | Eating disorder | | | | OCD | | | | Self harm | | | | Severe mental illness | | | |
|---|---|---|---|---|---|---|---|---|---|---|---|---|---|---|---|---|---|---|---|---|---|---|---|---|
| | Coefficient | SE | 95% CI | P value | Coefficient | SE | 95% CI | P value | Coefficient | SE | 95% CI | P value | Coefficient | SE | 95% CI | P value | Coefficient | SE | 95% CI | P value | Coefficient | SE | 95% CI | P value |
| **Pre-pandemic effects, by quintile of deprivation** | | | | | | | | | | | | | | | | | | | | | | | | |
| IMD 1 (least deprived), not living alone | -86.84 | 5.23 | (-97.11, -76.57) | <0.001 | -101.47 | 5.98 | (-113.23, -89.70 | <0.001 | -1.18 | 0.37 | (-1.90, -0.46) | 0.001 | -0.45 | 0.26 | (-0.97, 0.07) | 0.09 | -54.51 | 1.5 | (-57.46, -51.56) | <0.001 | -73.14 | 2.86 | (-78.76, -67.51) | <0.001 |
| IMD 2, living alone | -95.64 | 5.4 | (-106.25, -85.03) | <0.001 | -117.42 | 5.91 | (-129.03, -105.8 | <0.001 | -3.76 | 0.27 | (-4.30, -3.23) | <0.001 | -0.74 | 0.22 | (-1.17, -0.31) | 0.001 | -73.71 | 1.49 | (-76.65, -70.78) | <0.001 | -120.29 | 2.29 | (-124.80, -115. | <0.001 |
| IMD 2, not living alone | -105.07 | 5.19 | (-115.28, -94.87) | <0.001 | -120.73 | 6.03 | (-132.58, -108.8 | <0.001 | -2.27 | 0.35 | (-2.95, -1.58) | <0.001 | -0.75 | 0.26 | (-1.26, -0.24) | 0.004 | -63.79 | 1.5 | (-66.75, -60.84) | <0.001 | -88.57 | 2.84 | (-94.14, -82.99) | <0.001 |
| IMD 3, living alone | -110.56 | 5.25 | (-120.88, -100.24 | <0.001 | -135.75 | 5.94 | (-147.42, -124.0 | <0.001 | -4.33 | 0.27 | (-4.87, -3.79) | <0.001 | -0.9 | 0.21 | (-1.31, -0.49) | <0.001 | -78.23 | 1.47 | (-81.11, -75.35) | <0.001 | -125.56 | 2.26 | (-130.01, -121. | <0.001 |
| IMD 3, not living alone | -116.84 | 5.04 | (-126.75, -106.93 | <0.001 | -137.25 | 6.31 | (-149.64, -124.8 | <0.001 | -2.63 | 0.36 | (-3.34, -1.91) | <0.001 | -0.61 | 0.25 | (-1.10, -0.12) | 0.016 | -72.47 | 1.49 | (-75.39, -69.55) | <0.001 | -101.03 | 2.67 | (-106.29, -95.7 | <0.001 |
| IMD 4, living alone | -125.41 | 5.16 | (-135.55, -115.27 | <0.001 | -153.96 | 5.82 | (-165.40, -142.5 | <0.001 | -4.35 | 0.28 | (-4.90, -3.80) | <0.001 | -0.82 | 0.21 | (-1.24, -0.41) | <0.001 | -81.84 | 1.5 | (-84.79, -78.89) | <0.001 | -129.33 | 2.26 | (-133.77, -124. | <0.001 |
| IMD 4, not living alone | -22.04 | 5.97 | (-33.76, -10.31) | <0.001 | -39.18 | 6.51 | (-51.97, -26.39) | <0.001 | -3.23 | 0.3 | (-3.83, -2.64) | <0.001 | -0.37 | 0.22 | (-0.81, 0.06) | 0.093 | -52.07 | 1.64 | (-55.30, -48.84) | <0.001 | -100.69 | 2.31 | (-105.23, -96.1 | <0.001 |
| IMD 5 (most deprived), living alone | -54.79 | 5.59 | (-65.77, -43.80) | <0.001 | -62.49 | 6.28 | (-74.84, -50.15) | <0.001 | 0.65 | 0.46 | (-0.24, 1.55) | 0.152 | -0.15 | 0.23 | (-0.61, 0.31) | 0.514 | -38.45 | 1.62 | (-41.64, -35.27) | <0.001 | -45.22 | 2.88 | (-50.89, -39.56) | <0.001 |
| IMD 5 (most deprived), not living alone | -73.54 | 5.25 | (-83.86, -63.21) | <0.001 | -90.4 | 5.94 | (-102.07, -78.73 | <0.001 | -3.71 | 0.29 | (-4.28, -3.13) | <0.001 | -0.62 | 0.21 | (-1.04, -0.20) | 0.004 | -66.33 | 1.44 | (-69.17, -63.49) | <0.001 | -112.49 | 2.3 | (-117.00, -107. | <0.001 |
| **Effect of the pandemic in those living alo** | -69.05 | 6.69 | (-82.20,-55.90) | <0.001 | -103.46 | 6.86 | (-116.93,-89.98) | <0.001 | 0.43 | 0.37 | (-0.31,1.16) | 0.252 | -0.84 | 0.24 | (-1.30,-0.37) | <0.001 | -14.9 | 3.23 | (-21.25,-8.56) | <0.001 | -11.96 | 4.22 | (-20.26,-3.67) | 0.005 |
| **Interaction effects during pandemic** | | | | | | | | | | | | | | | | | | | | | | | | |
| IMD 1 (least deprived), not living alone | 25.82 | 10.32 | (5.54, 46.10) | 0.013 | 34.88 | 10.11 | (15.01, 54.75) | 0.001 | 0.38 | 0.55 | (-0.70, 1.46) | 0.494 | 0.18 | 0.28 | (-0.37, 0.74) | 0.522 | 15.64 | 3.51 | (8.74, 22.55) | <0.001 | 6.91 | 4.47 | (-1.88, 15.69) | 0.123 |
| IMD 2, living alone | 34.9 | 8.94 | (17.32, 52.47) | <0.001 | 38.98 | 8.68 | (21.92, 56.05) | <0.001 | -0.23 | 0.79 | (-1.78, 1.32) | 0.771 | 0.35 | 0.31 | (-0.26, 0.95) | 0.261 | 8.76 | 3.67 | (1.54, 15.98) | 0.018 | 4.21 | 5.14 | (-5.89, 14.30) | 0.413 |
| IMD 2, not living alone | 32.18 | 9.1 | (14.30, 50.06) | <0.001 | 46.97 | 8.5 | (30.26, 63.68) | <0.001 | 0.47 | 0.53 | (-0.56, 1.51) | 0.369 | 0.19 | 0.27 | (-0.34, 0.72) | 0.488 | 16.31 | 3.36 | (9.72, 22.91) | <0.001 | 8.56 | 4.4 | (-0.09, 17.21) | 0.052 |
| IMD 3, living alone | 32.12 | 8.27 | (15.86, 48.39) | <0.001 | 47.38 | 8.34 | (30.98, 63.78) | <0.001 | -0.05 | 0.69 | (-1.41, 1.31) | 0.939 | 0.34 | 0.32 | (-0.30, 0.98) | 0.293 | 9.08 | 3.46 | (2.28, 15.89) | 0.009 | 4.29 | 4.97 | (-5.48, 14.05) | 0.389 |
| IMD 3, not living alone | 34.1 | 8.71 | (16.98, 51.22) | <0.001 | 53.26 | 8.12 | (37.31, 69.21) | <0.001 | -0.02 | 0.52 | (-1.04, 0.99) | 0.965 | 0.15 | 0.28 | (-0.40, 0.70) | 0.589 | 15.99 | 3.34 | (9.43, 22.54) | <0.001 | 9.03 | 4.38 | (0.42, 17.63) | 0.04 |
| IMD 4, living alone | 34.7 | 8.31 | (18.37, 51.03) | <0.001 | 48.59 | 8.1 | (32.67, 64.51) | <0.001 | -0.36 | 0.54 | (-1.42, 0.69) | 0.499 | 0.39 | 0.33 | (-0.27, 1.04) | 0.246 | 10.92 | 3.41 | (4.22, 17.62) | 0.001 | 7.25 | 4.84 | (-2.26, 16.76) | 0.135 |
| IMD 4, not living alone | 36.2 | 8.75 | (19.00, 53.40) | <0.001 | 56.93 | 8.08 | (41.05, 72.81) | <0.001 | 0.16 | 0.52 | (-0.85, 1.18) | 0.752 | 0.47 | 0.28 | (-0.08, 1.01) | 0.093 | 15.57 | 3.33 | (9.03, 22.11) | <0.001 | 9.34 | 4.32 | (0.84, 17.83) | 0.031 |
| IMD 5 (most deprived), living alone | 33.47 | 8.11 | (17.54, 49.41) | <0.001 | 54.35 | 7.98 | (38.67, 70.03) | <0.001 | -1.28 | 0.64 | (-2.55, -0.02) | 0.047 | -0.05 | 0.31 | (-0.67, 0.56) | 0.867 | 11.5 | 3.34 | (4.94, 18.06) | 0.001 | 7.11 | 4.72 | (-2.17, 16.39) | 0.133 |
| IMD 5 (most deprived), not living alone | 36.13 | 8.61 | (19.21, 53.05) | <0.001 | 61.04 | 7.96 | (45.39, 76.69) | <0.001 | -0.16 | 0.51 | (-1.15, 0.84) | 0.76 | 0.37 | 0.27 | (-0.16, 0.91) | 0.171 | 15.58 | 3.34 | (9.02, 22.14) | <0.001 | 10.4 | 4.33 | (1.89, 18.90) | 0.017 |
| **Summer** | 6.99 | 2.82 | (1.46, 12.52) | 0.013 | 8.7 | 2.81 | (3.19, 14.22) | 0.002 | 0.42 | 0.21 | (0.00, 0.84) | 0.049 | 0 | 0.09 | (-0.18, 0.18) | 0.982 | 3 | 0.59 | (1.84, 4.16) | <0.001 | 0.34 | 0.98 | (-1.58, 2.27) | 0.725 |
| **Autumn** | 23.93 | 2.81 | (18.40, 29.46) | <0.001 | 24.69 | 3 | (18.79, 30.58) | <0.001 | 0.26 | 0.21 | (-0.15, 0.66) | 0.222 | 0.08 | 0.09 | (-0.09, 0.25) | 0.367 | -0.39 | 0.51 | (-1.40, 0.62) | 0.448 | 2.45 | 0.95 | (0.59, 4.31) | 0.01 |
| **Winter** | 3.43 | 3.2 | (-2.86, 9.71) | 0.284 | 8.29 | 3.03 | (2.34, 14.24) | 0.006 | 0.05 | 0.22 | (-0.38, 0.47) | 0.828 | 0 | 0.09 | (-0.18, 0.17) | 0.992 | -2.04 | 0.62 | (-3.26, -0.83) | 0.001 | 3.76 | 1.2 | (1.41, 6.12) | 0.002 |
| **Constant** | 316.6 | 4.95 | (306.86, 326.33) | <0.001 | 294.85 | 5.82 | (283.40, 306.29 | <0.001 | 10.48 | 0.29 | (9.91, 11.06) | <0.001 | 3.63 | 0.2 | (3.24, 4.02) | <0.001 | 97.07 | 1.41 | (94.31, 99.83) | <0.001 | 148.19 | 2.26 | (143.74, 152.6 | <0.001 |

**NB: Except where specified, coefficients relate to effects in the baseline categories of all variables (pre-pandemic, spring, living alone, IMD quintile 1)**

### Table S15. Region-stratified estimates from EHR analyses

| | Anxiety | | | | Depression | | | | Eating disorder | | | | OCD | | | | Self harm | | | | Severe mental illness | | | |
|---|---|---|---|---|---|---|---|---|---|---|---|---|---|---|---|---|---|---|---|---|---|---|---|---|
| | Coefficient | SE | 95% CI | P value | Coefficient | SE | 95% CI | P value | Coefficient | SE | 95% CI | P value | Coefficient | SE | 95% CI | P value | Coefficient | SE | 95% CI | P value | Coefficient | SE | 95% CI | P value |
| **Pre-pandemic effects** | | | | | | | | | | | | | | | | | | | | | | | | |
| East midlands, not living alone | -67.3 | 12.38 | (-91.60, -43.00) | <0.001 | -60.55 | 14.8 | (-89.61, -31.50) | <0.001 | 3.3 | 0.58 | (2.15, 4.44) | <0.001 | -0.04 | 0.35 | (-0.72, 0.64) | 0.916 | -36.67 | 1.08 | (-38.79, -34.56) | <0.001 | 6.96 | 2.88 | (1.31, 12.60) | 0.016 |
| East, living alone | -135.7 | 5.33 | (-146.15, -125.25 | <0.001 | -132.77 | 6.46 | (-145.46, -120.0 | <0.001 | -1.26 | 0.35 | (-1.95, -0.57) | <0.001 | -1.18 | 0.27 | (-1.70, -0.66) | <0.001 | -49.06 | 1 | (-51.02, -47.09) | <0.001 | -57.05 | 2.01 | (-61.01, -53.10) | <0.001 |
| East, not living alone | 28.72 | 6.45 | (16.06, 41.39) | <0.001 | -3.41 | 8.78 | (-20.65, 13.83) | 0.698 | 4.34 | 0.59 | (3.19, 5.50) | <0.001 | -0.11 | 0.34 | (-0.78, 0.57) | 0.757 | 2.03 | 1.64 | (-1.18, 5.25) | 0.215 | -12.31 | 3.45 | (-19.08, -5.54) | <0.001 |
| London, living alone | 6.36 | 5.36 | (-4.16, 16.88) | 0.236 | -38.16 | 6.69 | (-51.29, -25.03) | <0.001 | -1.06 | 0.41 | (-1.85, -0.26) | 0.01 | 0 | 0.3 | (-0.59, 0.59) | 0.997 | -30.02 | 1.14 | (-32.25, -27.78) | <0.001 | -62.74 | 1.97 | (-66.61, -58.87) | <0.001 |
| London, not living alone | 48.82 | 5.76 | (37.51, 60.13) | <0.001 | 33.24 | 8.13 | (17.28, 49.19) | <0.001 | -0.32 | 0.49 | (-1.29, 0.64) | 0.511 | -0.23 | 0.29 | (-0.79, 0.33) | 0.422 | 9.89 | 1.57 | (6.81, 12.98) | <0.001 | 0.73 | 2.99 | (-5.13, 6.59) | 0.807 |
| North East, living alone | 55.74 | 6.09 | (43.80, 67.69) | <0.001 | 5.77 | 7.01 | (-7.99, 19.53) | 0.411 | -2.3 | 0.34 | (-2.96, -1.63) | <0.001 | -0.17 | 0.28 | (-0.72, 0.37) | 0.533 | -24.43 | 1.12 | (-26.64, -22.23) | <0.001 | -59.25 | 2.04 | (-63.26, -55.24) | <0.001 |
| North East, not living alone | -37.22 | 5.04 | (-47.11, -27.33) | <0.001 | -59.83 | 6.67 | (-72.93, -46.73) | <0.001 | 3.37 | 0.46 | (2.47, 4.27) | <0.001 | 0.01 | 0.34 | (-0.67, 0.69) | 0.979 | -4.64 | 1.62 | (-7.81, -1.46) | 0.004 | -4.82 | 3 | (-10.70, 1.07) | 0.109 |
| North West, living alone | -54.61 | 5.27 | (-64.95, -44.27) | <0.001 | -72 | 6.46 | (-84.68, -59.32) | <0.001 | -0.23 | 0.37 | (-0.95, 0.49) | 0.525 | -0.04 | 0.27 | (-0.57, 0.50) | 0.89 | -31.82 | 1.21 | (-34.20, -29.45) | <0.001 | -60.07 | 2.04 | (-64.07, -56.07) | <0.001 |
| North West, not living alone | -76.31 | 4.93 | (-85.99, -66.62) | <0.001 | -69.5 | 7.3 | (-83.84, -55.17) | <0.001 | 0.79 | 0.43 | (-0.06, 1.64) | 0.069 | -0.68 | 0.32 | (-1.30, -0.06) | 0.032 | -13.44 | 1.09 | (-15.59, -11.29) | <0.001 | -14.02 | 3.05 | (-20.01, -8.03) | <0.001 |
| South East, living alone | -81.61 | 4.66 | (-90.76, -72.46) | <0.001 | -79.48 | 6.66 | (-92.56, -66.40) | <0.001 | -0.91 | 0.35 | (-1.59, -0.23) | 0.008 | -0.96 | 0.26 | (-1.46, -0.45) | <0.001 | -33.75 | 1.04 | (-35.78, -31.71) | <0.001 | -63.85 | 2.11 | (-67.98, -59.71) | <0.001 |
| South East, not living alone | -4.37 | 7.57 | (-19.22, 10.48) | 0.564 | -14.56 | 10 | (-34.19, 5.07) | 0.146 | 2.89 | 0.72 | (1.48, 4.31) | <0.001 | -0.55 | 0.4 | (-1.34, 0.23) | 0.167 | -16.52 | 1.53 | (-19.52, -13.53) | <0.001 | 6.68 | 2.87 | (1.06, 12.31) | 0.02 |
| South West, living alone | -2.14 | 5.4 | (-12.74, 8.45) | 0.692 | -42.45 | 7.62 | (-57.40, -27.50) | <0.001 | 0.07 | 0.59 | (-1.08, 1.22) | 0.902 | -0.22 | 0.32 | (-0.85, 0.42) | 0.502 | -33.18 | 1.27 | (-35.68, -30.68) | <0.001 | -50.85 | 2.04 | (-54.86, -46.84) | <0.001 |
| South West, not living alone | 17.85 | 5.74 | (6.58, 29.12) | 0.002 | 6.88 | 7.06 | (-6.98, 20.75) | 0.33 | -0.17 | 0.47 | (-1.09, 0.74) | 0.711 | -0.85 | 0.28 | (-1.40, -0.29) | 0.003 | -1.73 | 1.19 | (-4.07, 0.61) | 0.147 | 0.09 | 2.93 | (-5.67, 5.84) | 0.977 |
| West Midlands, living alone | 2.01 | 5.26 | (-8.31, 12.32) | 0.703 | -27.66 | 6.5 | (-40.43, -14.89) | <0.001 | -2.86 | 0.34 | (-3.53, -2.19) | <0.001 | -0.7 | 0.26 | (-1.20, -0.20) | 0.007 | -31.4 | 0.96 | (-33.29, -29.51) | <0.001 | -58.61 | 2.09 | (-62.73, -54.50) | <0.001 |
| West Midlands, not living alone | -10.58 | 4.92 | (-20.23, -0.92) | 0.032 | -17.99 | 7.04 | (-31.81, -4.16) | 0.011 | -2.41 | 0.33 | (-3.06, -1.75) | <0.001 | -0.39 | 0.26 | (-0.90, 0.11) | 0.128 | -29.1 | 1.11 | (-31.28, -26.92) | <0.001 | -59.06 | 1.95 | (-62.90, -55.23) | <0.001 |
| Yorkshire and The Humber, living alone | -24.35 | 5.3 | (-34.75, -13.95) | <0.001 | -41.75 | 7.11 | (-55.71, -27.79) | <0.001 | 1.03 | 0.39 | (0.27, 1.79) | 0.008 | 0.48 | 0.31 | (-0.13, 1.09) | 0.126 | -7.03 | 1.11 | (-9.21, -4.85) | <0.001 | 3.14 | 3.02 | (-2.79, 9.08) | 0.299 |
| Yorkshire and The Humber, not living alor | -48.11 | 5.28 | (-58.47, -37.74) | <0.001 | -68.8 | 6.39 | (-81.34, -56.25) | <0.001 | -1.82 | 0.32 | (-2.45, -1.19) | <0.001 | -0.37 | 0.26 | (-0.87, 0.13) | 0.151 | -35.17 | 1.08 | (-37.29, -33.06) | <0.001 | -59.65 | 2.11 | (-63.78, -55.51) | <0.001 |
| **Effect of the pandemic in those living a** | -70.19 | 5.78 | (-81.54,-58.84) | <0.001 | -100.93 | 6.43 | (-113.56,-88.31) | <0.001 | 0.1 | 0.55 | (-0.98,1.17) | 0.861 | -1.21 | 0.27 | (-1.74,-0.68) | <0.001 | -3.87 | 2.23 | (-8.25,0.51) | 0.083 | -7.06 | 2.75 | (-12.46,-1.66) | 0.01 |
| **Interaction effects during pandemic** | | | | | | | | | | | | | | | | | | | | | | | | |
| East midlands, not living alone | 16.09 | 8.18 | (0.03, 32.14) | 0.05 | 24.24 | 8.49 | (7.57, 40.91) | 0.004 | 0.13 | 0.62 | (-1.08, 1.34) | 0.833 | 0.44 | 0.32 | (-0.18, 1.06) | 0.167 | 6.75 | 2.54 | (1.76, 11.73) | 0.008 | 4.06 | 2.96 | (-1.76, 9.87) | 0.171 |
| East, living alone | 12.71 | 7.68 | (-2.37, 27.79) | 0.098 | 24.45 | 8.13 | (8.48, 40.42) | 0.003 | -0.43 | 0.67 | (-1.74, 0.87) | 0.515 | -0.21 | 0.35 | (-0.89, 0.48) | 0.552 | -4.79 | 2.71 | (-10.11, 0.54) | 0.078 | -1.45 | 4.31 | (-9.90, 7.00) | 0.737 |
| East, not living alone | 32.56 | 8.52 | (15.84, 49.27) | <0.001 | 51.47 | 7.89 | (35.98, 66.96) | <0.001 | 0.31 | 0.65 | (-0.98, 1.59) | 0.64 | 0.65 | 0.3 | (0.06, 1.23) | 0.031 | 4.43 | 2.41 | (-0.31, 9.16) | 0.067 | 3.55 | 3.07 | (-2.48, 9.58) | 0.248 |
| London, living alone | 169.32 | 17.14 | (135.69, 202.96) | <0.001 | 194.41 | 19.07 | (156.98, 231.84 | <0.001 | 2.12 | 1.05 | (0.07, 4.18) | 0.043 | 3.27 | 0.53 | (2.23, 4.32) | <0.001 | 3.3 | 2.56 | (-1.71, 8.32) | 0.197 | -5.1 | 4.09 | (-13.13, 2.94) | 0.213 |
| London, not living alone | 81.42 | 8.31 | (65.12, 97.72) | <0.001 | 106.16 | 8.36 | (89.74, 122.57) | <0.001 | 0.75 | 0.74 | (-0.70, 2.21) | 0.31 | 1.45 | 0.32 | (0.83, 2.07) | <0.001 | 4.57 | 2.37 | (-0.08, 9.22) | 0.054 | 4.38 | 3.09 | (-1.68, 10.45) | 0.156 |
| North East, living alone | -4.56 | 8.66 | (-21.56, 12.44) | 0.599 | 6.03 | 9.92 | (-13.43, 25.50) | 0.543 | -0.74 | 1.23 | (-3.15, 1.68) | 0.549 | 0.24 | 0.41 | (-0.57, 1.05) | 0.561 | -8.67 | 3.27 | (-15.10, -2.25) | 0.008 | 11.63 | 4.67 | (2.47, 20.79) | 0.013 |
| North East, not living alone | 28.12 | 8.82 | (10.82, 45.43) | 0.001 | 50.67 | 8.92 | (33.16, 68.17) | <0.001 | -0.25 | 0.77 | (-1.76, 1.26) | 0.748 | 0.25 | 0.38 | (-0.49, 0.99) | 0.509 | 4.43 | 2.46 | (-0.41, 9.27) | 0.073 | 6.04 | 3.03 | (0.09, 11.99) | 0.047 |
| North West, living alone | -0.65 | 9.39 | (-19.08, 17.79) | 0.945 | -3.07 | 9.72 | (-22.15, 16.00) | 0.752 | -0.16 | 0.79 | (-1.70, 1.38) | 0.84 | 0.78 | 0.38 | (0.03, 1.53) | 0.042 | -5.19 | 3.03 | (-11.14, 0.76) | 0.087 | -2.66 | 4.48 | (-11.46, 6.13) | 0.552 |
| North West, not living alone | 21.19 | 10.68 | (0.24, 42.15) | 0.047 | 35.16 | 9.8 | (15.93, 54.39) | <0.001 | 0.72 | 0.67 | (-0.59, 2.03) | 0.282 | 0.51 | 0.34 | (-0.16, 1.19) | 0.134 | 6.56 | 2.51 | (1.63, 11.50) | 0.009 | 3.31 | 3.04 | (-2.67, 9.28) | 0.278 |
| South East, living alone | 25.83 | 8.99 | (8.17, 43.48) | 0.004 | 40.27 | 8.39 | (23.81, 56.73) | <0.001 | -1.22 | 0.87 | (-2.94, 0.49) | 0.162 | 0.68 | 0.44 | (-0.19, 1.54) | 0.125 | 0.47 | 3.38 | (-6.16, 7.11) | 0.889 | -1.3 | 4.18 | (-9.51, 6.92) | 0.757 |
| South East, not living alone | 42.22 | 9.31 | (23.95, 60.50) | <0.001 | 55.1 | 8 | (39.41, 70.80) | <0.001 | 0.91 | 0.73 | (-0.52, 2.34) | 0.21 | 0.7 | 0.33 | (0.05, 1.34) | 0.034 | 6.04 | 2.44 | (1.25, 10.83) | 0.014 | 3.33 | 2.97 | (-2.50, 9.17) | 0.263 |
| South West, living alone | 35.69 | 7.33 | (21.30, 50.08) | <0.001 | 41.17 | 7.97 | (25.53, 56.81) | <0.001 | -0.08 | 0.8 | (-1.65, 1.49) | 0.922 | 0.26 | 0.37 | (-0.47, 0.98) | 0.486 | -2.58 | 2.67 | (-7.83, 2.67) | 0.335 | 1.17 | 4.12 | (-6.92, 9.27) | 0.776 |
| South West, not living alone | 44.96 | 7.85 | (29.56, 60.36) | <0.001 | 54.19 | 7.77 | (38.94, 69.44) | <0.001 | 0.72 | 0.69 | (-0.63, 2.08) | 0.295 | 0.7 | 0.31 | (0.08, 1.31) | 0.026 | 4.58 | 2.37 | (-0.08, 9.23) | 0.054 | 5.12 | 3.04 | (-0.84, 11.08) | 0.092 |
| West Midlands, living alone | 7.28 | 9.61 | (-11.59, 26.14) | 0.449 | 14.24 | 10.62 | (-6.61, 35.09) | 0.18 | -0.35 | 1.2 | (-2.71, 2.01) | 0.77 | 0.63 | 0.5 | (-0.34, 1.61) | 0.204 | -0.99 | 3.01 | (-6.89, 4.91) | 0.742 | 1 | 5.4 | (-9.60, 11.60) | 0.853 |
| West Midlands, not living alone | 13.81 | 8.04 | (-1.98, 29.60) | 0.087 | 39.22 | 8.68 | (22.18, 56.27) | <0.001 | 0.33 | 0.81 | (-1.26, 1.91) | 0.687 | 0.66 | 0.36 | (-0.05, 1.37) | 0.067 | 3.25 | 2.53 | (-1.71, 8.21) | 0.198 | 3.82 | 3.12 | (-2.30, 9.95) | 0.221 |
| Yorkshire and The Humber, living alone | 19.89 | 8.47 | (3.26, 36.52) | 0.019 | 14.94 | 8.56 | (-1.86, 31.74) | 0.081 | 0.28 | 0.82 | (-1.32, 1.88) | 0.732 | 0.93 | 0.34 | (0.27, 1.60) | 0.006 | -6.47 | 2.68 | (-11.74, -1.20) | 0.016 | -1.62 | 4.44 | (-10.35, 7.10) | 0.715 |
| Yorkshire and The Humber, not living alor | 40.87 | 9.11 | (22.98, 58.75) | <0.001 | 47.3 | 8.66 | (30.30, 64.30) | <0.001 | 1.02 | 0.7 | (-0.36, 2.40) | 0.147 | 0.71 | 0.31 | (0.11, 1.31) | 0.021 | 2.57 | 2.34 | (-2.02, 7.16) | 0.272 | 3.31 | 3.02 | (-2.61, 9.24) | 0.273 |
| **Summer** | 6.42 | 2.42 | (1.67, 11.17) | 0.008 | 9.14 | 2.46 | (4.31, 13.97) | <0.001 | 0.21 | 0.22 | (-0.22, 0.65) | 0.343 | -0.04 | 0.09 | (-0.22, 0.13) | 0.62 | 2.81 | 0.48 | (1.87, 3.75) | <0.001 | 0.07 | 0.83 | (-1.55, 1.70) | 0.929 |
| **Autumn** | 22.97 | 2.54 | (17.98, 27.96) | <0.001 | 25.23 | 2.68 | (19.97, 30.49) | <0.001 | 0.15 | 0.21 | (-0.27, 0.57) | 0.486 | 0.03 | 0.09 | (-0.14, 0.21) | 0.699 | -0.74 | 0.46 | (-1.64, 0.16) | 0.108 | 2.16 | 0.84 | (0.51, 3.80) | 0.01 |
| **Winter** | 2.87 | 2.78 | (-2.60, 8.33) | 0.304 | 8.33 | 2.77 | (2.89, 13.76) | 0.003 | -0.04 | 0.22 | (-0.46, 0.39) | 0.866 | -0.06 | 0.09 | (-0.24, 0.12) | 0.511 | -2.21 | 0.51 | (-3.22, -1.20) | <0.001 | 3.07 | 1 | (1.10, 5.04) | 0.002 |
| **Constant** | 263.1 | 4.5 | (254.27, 271.94) | <0.001 | 237.86 | 6.27 | (225.55, 250.18 | <0.001 | 8.44 | 0.33 | (7.79, 9.09) | <0.001 | 3.46 | 0.25 | (2.98, 3.94) | <0.001 | 59.53 | 0.91 | (57.75, 61.31) | <0.001 | 90.35 | 1.94 | (86.55, 94.15) | <0.001 |
| **NB: Except where specified, coefficients relate to effects in the baseline categories of all variables (pre-pandemic, spring, living alone, East)** | | | | | | | | | | | | | | | | | | | | | | | | |

### Table S16. Shielded-stratified estimates from EHR analyses

| | Anxiety | | | | Depression | | | | Eating disorder | | | | OCD | | | | Self harm | | | | Severe mental illness | | | |
|---|---|---|---|---|---|---|---|---|---|---|---|---|---|---|---|---|---|---|---|---|---|---|---|---|
| | Coefficient | SE | 95% CI | P value | Coefficient | SE | 95% CI | P value | Coefficient | SE | 95% CI | P value | Coefficient | SE | 95% CI | P value | Coefficient | SE | 95% CI | P value | Coefficient | SE | 95% CI | P value |
| **Pre-pandemic effects** | | | | | | | | | | | | | | | | | | | | | | | | |
| Not shielding, not living alone | -17.97 | 3.66 | (-25.20, -10.75) | <0.001 | -28.01 | 3.71 | (-35.33, -20.69) | <0.001 | -2.84 | 0.2 | (-3.23, -2.44) | <0.001 | -0.44 | 0.12 | (-0.68, -0.19) | <0.001 | -24.88 | 0.89 | (-26.64, -23.12) | <0.001 | -49.6 | 1.88 | (-53.30, -45.90) | <0.001 |
| Shielding, living alone | -38.19 | 3.7 | (-45.49, -30.89) | <0.001 | -11 | 5.62 | (-22.10, 0.10) | 0.052 | 3.65 | 0.48 | (2.71, 4.60) | <0.001 | -1.52 | 0.16 | (-1.82, -1.21) | <0.001 | 13.64 | 1.22 | (11.24, 16.04) | <0.001 | 110.49 | 5.29 | (100.05, 120.93 | <0.001 |
| Shielding, not living alone | -41.05 | 3.71 | (-48.37, -33.73) | <0.001 | -15.79 | 4.71 | (-25.09, -6.50) | 0.001 | 2.32 | 0.36 | (1.61, 3.03) | <0.001 | -1.26 | 0.13 | (-1.52, -1.01) | <0.001 | -16.94 | 0.8 | (-18.52, -15.36) | <0.001 | 15.12 | 2.82 | (9.56, 20.68) | <0.001 |
| **Effect of the pandemic in tho** | -45.57 | 5.34 | (-56.10,-35.03) | <0.001 | -66.38 | 4.89 | (-76.03,-56.73) | <0.001 | -0.15 | 0.42 | (-0.99,0.68) | 0.721 | -0.67 | 0.15 | (-0.96,-0.38) | <0.001 | -7.28 | 1.39 | (-10.03,-4.53) | <0.001 | -8.58 | 2.46 | (-13.44,-3.71) | 0.001 |
| **Interaction effects during pandemic** | | | | | | | | | | | | | | | | | | | | | | | | |
| Not shielding, not living alone | 7.66 | 8.13 | (-8.39, 23.72) | 0.347 | 13.99 | 6.96 | (0.25, 27.73) | 0.046 | 0.76 | 0.54 | (-0.31, 1.83) | 0.163 | 0.09 | 0.18 | (-0.27, 0.45) | 0.629 | 8.22 | 1.66 | (4.94, 11.51) | <0.001 | 5.63 | 2.82 | (0.07, 11.19) | 0.047 |
| Shielding, living alone | 26.48 | 6.63 | (13.40, 39.56) | <0.001 | -0.35 | 7.42 | (-14.99, 14.30) | 0.963 | 2.33 | 0.73 | (0.89, 3.76) | 0.002 | 0.32 | 0.22 | (-0.12, 0.75) | 0.152 | 3.29 | 2.39 | (-1.43, 8.01) | 0.17 | 17.48 | 7.83 | (2.03, 32.92) | 0.027 |
| Shielding, not living alone | 31.99 | 7.48 | (17.24, 46.75) | <0.001 | 9.99 | 6.9 | (-3.63, 23.60) | 0.15 | 0.72 | 0.62 | (-0.51, 1.95) | 0.249 | 0.23 | 0.19 | (-0.14, 0.60) | 0.227 | 8.36 | 1.76 | (4.88, 11.83) | <0.001 | 5.78 | 4.27 | (-2.66, 14.21) | 0.178 |
| **Summer** | 7.76 | 3.94 | (-0.02, 15.53) | 0.05 | 11.14 | 4.28 | (2.69, 19.59) | 0.01 | 0.75 | 0.37 | (0.03, 1.47) | 0.041 | 0.05 | 0.1 | (-0.15, 0.24) | 0.637 | 3.88 | 0.98 | (1.94, 5.82) | <0.001 | 3.07 | 2.99 | (-2.84, 8.97) | 0.307 |
| **Autumn** | 22.08 | 3.92 | (14.34, 29.82) | <0.001 | 24.33 | 4.66 | (15.13, 33.52) | <0.001 | 0.62 | 0.37 | (-0.10, 1.35) | 0.091 | 0.08 | 0.1 | (-0.12, 0.28) | 0.423 | 0.08 | 0.91 | (-1.71, 1.87) | 0.929 | 7.19 | 2.96 | (1.35, 13.03) | 0.016 |
| **Winter** | 1.59 | 4.58 | (-7.46, 10.63) | 0.729 | 7.9 | 4.48 | (-0.94, 16.74) | 0.08 | 0.23 | 0.39 | (-0.54, 1.01) | 0.554 | 0.05 | 0.11 | (-0.16, 0.27) | 0.629 | -2.19 | 1.01 | (-4.19, -0.19) | 0.032 | 7.9 | 3.51 | (0.96, 14.83) | 0.026 |
| **Constant** | 251.41 | 3.65 | (244.21, 258.61) | <0.001 | 213.94 | 4.28 | (205.48, 222.39 | <0.001 | 8.83 | 0.3 | (8.25, 9.41) | <0.001 | 3.41 | 0.12 | (3.18, 3.64) | <0.001 | 50.53 | 0.87 | (48.81, 52.24) | <0.001 | 72.23 | 2.55 | (67.21, 77.26) | <0.001 |
| **NB: Except where specified, coefficients relate to effects in the baseline categories of all variables (pre-pandemic, spring, living alone, not shielding)** | | | | | | | | | | | | | | | | | | | | | | | | |

### Table S17. Previous mental illness-stratified estimates from EHR analyses

| | Anxiety | | | | Depression | | | | Eating disorder | | | | OCD | | | | Self harm | | | | Severe mental illness | | | |
|---|---|---|---|---|---|---|---|---|---|---|---|---|---|---|---|---|---|---|---|---|---|---|---|---|
| | Coefficient | SE | 95% CI | P value | Coefficient | SE | 95% CI | P value | Coefficient | SE | 95% CI | P value | Coefficient | SE | 95% CI | P value | Coefficient | SE | 95% CI | P value | Coefficient | SE | 95% CI | P value |
| **Pre-pandemic effects** | | | | | | | | | | | | | | | | | | | | | | | | |
| No previous mental illness, not living alone | 2.24 | 5.98 | (-9.56, 14.05) | 0.708 | -3.95 | 5.46 | (-14.72, 6.82) | 0.47 | -0.43 | 0.24 | (-0.91, 0.05) | 0.078 | -0.03 | 0.06 | (-0.15, 0.10) | 0.683 | -3.55 | 1.95 | (-7.39, 0.29) | 0.07 | -9.96 | 1.44 | (-12.81, -7.11) | <0.001 |
| Previous mental illness, living alone | 907.96 | 13.31 | (881.69, 934.23) | <0.001 | 809.36 | 16.47 | (776.86, 841.85 | <0.001 | 46.6 | 1.1 | (44.43, 48.78) | <0.001 | 14.41 | 0.46 | (13.51, 15.31 | <0.001 | 264.23 | 4.25 | (255.83, 272.63 | <0.001 | 483.38 | 8.53 | (466.54, 500.21 | <0.001 |
| Previous mental illness, not living alone | 943.07 | 12.03 | (919.32, 966.82) | <0.001 | 785.88 | 14.91 | (756.46, 815.29 | <0.001 | 35.75 | 0.59 | (34.59, 36.91) | <0.001 | 14.82 | 0.35 | (14.12, 15.51 | <0.001 | 135.55 | 2.46 | (130.68, 140.41 | <0.001 | 189.25 | 3.74 | (181.86, 196.64 | <0.001 |
| **Effect of the pandemic in those living alone** | -15.48 | 6.43 | (-28.18,-2.78) | 0.017 | -31.69 | 6.01 | (-43.55,-19.83) | <0.001 | -0.08 | 0.27 | (-0.62,0.45) | 0.766 | -0.22 | 0.08 | (-0.38,-0.06) | 0.006 | -1.66 | 1.92 | (-5.46,2.13) | 0.388 | -2.66 | 1.52 | (-5.67,0.35) | 0.083 |
| **Interaction effects during pandemic** | | | | | | | | | | | | | | | | | | | | | | | | |
| No previous mental illness, not living alone | 2.34 | 9.18 | (-15.79, 20.46) | 0.799 | 8.2 | 8.37 | (-8.31, 24.70) | 0.329 | 0.14 | 0.37 | (-0.60, 0.87) | 0.716 | 0.08 | 0.09 | (-0.11, 0.27) | 0.406 | 1.58 | 2.77 | (-3.88, 7.03) | 0.57 | 1.61 | 2.1 | (-2.53, 5.76) | 0.444 |
| Previous mental illness, living alone | -188.7 | 20.92 | (-229.98, -147.42 | <0.001 | -238.8 | 22.13 | (-282.47, -195.1 | <0.001 | 0.46 | 2.14 | (-3.76, 4.68) | 0.831 | -2.69 | 0.63 | (-3.94, -1.43) | <0.001 | -38.59 | 8.33 | (-55.03, -22.16) | <0.001 | -36.73 | 14.39 | (-65.13, -8.32) | 0.012 |
| Previous mental illness, not living alone | -203.31 | 23.87 | (-250.42, -156.20 | <0.001 | -241.42 | 22.62 | (-286.07, -196.7 | <0.001 | 2.99 | 1.86 | (-0.68, 6.65) | 0.11 | -3.49 | 0.55 | (-4.57, -2.40) | <0.001 | 4.3 | 4.28 | (-4.16, 12.75) | 0.317 | -20.27 | 6.67 | (-33.44, -7.10) | 0.003 |
| **Summer** | 14.67 | 12.41 | (-9.82, 39.15) | 0.239 | 16.56 | 12.65 | (-8.41, 41.52) | 0.192 | 1.41 | 1.12 | (-0.79, 3.61) | 0.208 | -0.02 | 0.34 | (-0.68, 0.64) | 0.953 | 8.46 | 3.13 | (2.29, 14.64) | 0.007 | 2.49 | 5.78 | (-8.92, 13.90) | 0.667 |
| **Autumn** | 47.36 | 13.56 | (20.60, 74.12) | 0.001 | 49.27 | 14.58 | (20.50, 78.03) | 0.001 | 0.54 | 1.03 | (-1.50, 2.57) | 0.605 | 0.05 | 0.34 | (-0.62, 0.71) | 0.89 | -1.13 | 2.59 | (-6.23, 3.97) | 0.663 | 8.44 | 5.64 | (-2.69, 19.56) | 0.136 |
| **Winter** | 1.32 | 13.8 | (-25.92, 28.55) | 0.924 | 10.99 | 13.11 | (-14.88, 36.85) | 0.403 | -0.13 | 1.11 | (-2.33, 2.07) | 0.908 | -0.04 | 0.33 | (-0.69, 0.61) | 0.905 | -6.72 | 3.26 | (-13.15, -0.29) | 0.04 | 9.65 | 6.91 | (-3.98, 23.28) | 0.164 |
| **Constant** | 100.63 | 9.66 | (81.57, 119.70) | <0.001 | 81.2 | 9.71 | (62.03, 100.36) | <0.001 | 2.12 | 0.73 | (0.67, 3.57) | 0.004 | 1.08 | 0.22 | (0.64, 1.51) | <0.001 | 12.17 | 2.09 | (8.05, 16.29) | <0.001 | 11.38 | 4.17 | (3.16, 19.60) | 0.007 |
| **NB: Except where specified, coefficients relate to effects in the baseline categories of all variables (pre-pandemic, spring, living alone, no previous mental illness)** | | | | | | | | | | | | | | | | | | | | | | | | |

#### Table S18. Heterogeneity between estimates of psychological distress explained by cohort differences (meta-regression analyses)

| Measure | Timepoint | Heterogeneity ($I^2$) | Heterogeneity Explained (%) |
|---|---|---|---|
| **Time between pre-pandemic and first pandemic measure** | 0 | 65.7 | -7.2 |
| | 1 | 89.2 | -2.0 |
| | 2 | 85.9 | -3.1 |
| | 3 | 80.2 | -4.2 |
| **General population representativeness** | 0 | 58.0 | 0.6 |
| | 1 | 88.0 | -0.8 |
| | 2 | 82.7 | 0.1 |
| | 3 | 77.7 | -1.6 |
| **Type of measure for mental health used** | 0 | 19.4 | 39.1 |
| | 1 | 82.0 | 5.2 |
| | 2 | 79.9 | 2.9 |
| | 3 | 71.8 | 4.2 |

### Table S19 Leave-One-Out Analysis

Meta-analysed standardised mean differences for psychological distress when named cohort is removed from analysis.

| Time Period 0 | | | |
|---|---|---|---|
| Cohort | SMD | Lower CI | Upper CI |
| All Included | 0.09 | 0.04 | 0.14 |
| ALSPAC-G0 | 0.08 | 0.03 | 0.14 |
| ALSPAC-G1 | 0.09 | 0.04 | 0.14 |
| BCS | 0.10 | 0.05 | 0.15 |
| ELSA | 0.07 | 0.03 | 0.12 |
| GS | 0.09 | 0.04 | 0.15 |
| NCDS | 0.09 | 0.04 | 0.15 |
| NS | 0.10 | 0.04 | 0.15 |
| NSHD | 0.10 | 0.05 | 0.16 |
| TwinsUK | 0.10 | 0.04 | 0.15 |
| USOC | 0.11 | 0.06 | 0.16 |

| Time Period 1 | | | |
|---|---|---|---|
| Cohort | SMD | Lower CI | Upper CI |
| All Included | 0.19 | 0.09 | 0.29 |
| ALSPAC-G0 | 0.15 | 0.07 | 0.23 |
| ALSPAC-G1 | 0.18 | 0.08 | 0.29 |
| BCS | 0.22 | 0.12 | 0.31 |
| ELSA | 0.18 | 0.07 | 0.29 |
| GS | 0.20 | 0.09 | 0.31 |
| NCDS | 0.21 | 0.10 | 0.31 |
| NS | 0.18 | 0.08 | 0.28 |
| NSHD | 0.18 | 0.08 | 0.29 |
| TwinsUK | 0.21 | 0.11 | 0.31 |
| USOC | 0.19 | 0.08 | 0.31 |

| Time Period 2 | | | |
|---|---|---|---|
| Cohort | SMD | Lower CI | Upper CI |
| All Included | 0.15 | 0.06 | 0.25 |
| ALSPAC-G0 | 0.11 | 0.05 | 0.18 |
| ALSPAC-G1 | 0.15 | 0.05 | 0.25 |
| BCS | 0.17 | 0.07 | 0.27 |
| ELSA* | 0.15 | 0.06 | 0.25 |
| GS | 0.15 | 0.04 | 0.25 |
| NCDS | 0.17 | 0.07 | 0.27 |
| NS | 0.15 | 0.05 | 0.26 |
| NSHD | 0.15 | 0.05 | 0.25 |
| TwinsUK | 0.18 | 0.08 | 0.27 |
| USOC | 0.16 | 0.05 | 0.26 |

| Time Period 3 | | | |
|---|---|---|---|
| Cohort | SMD | Lower CI | Upper CI |
| All Included | 0.15 | 0.07 | 0.22 |
| ALSPAC-G0 | 0.12 | 0.06 | 0.19 |
| ALSPAC-G1 | 0.14 | 0.06 | 0.21 |
| BCS | 0.15 | 0.07 | 0.23 |
| ELSA | 0.14 | 0.06 | 0.21 |
| GS | 0.14 | 0.07 | 0.22 |
| NCDS | 0.17 | 0.09 | 0.24 |
| NS | 0.15 | 0.07 | 0.23 |
| NSHD | 0.15 | 0.07 | 0.23 |
| TwinsUK | 0.17 | 0.10 | 0.24 |
| USOC | 0.15 | 0.07 | 0.23 |

* ELSA does not contain information for Time Period 2

#### Figure S1. Results of meta-analyses in LPS

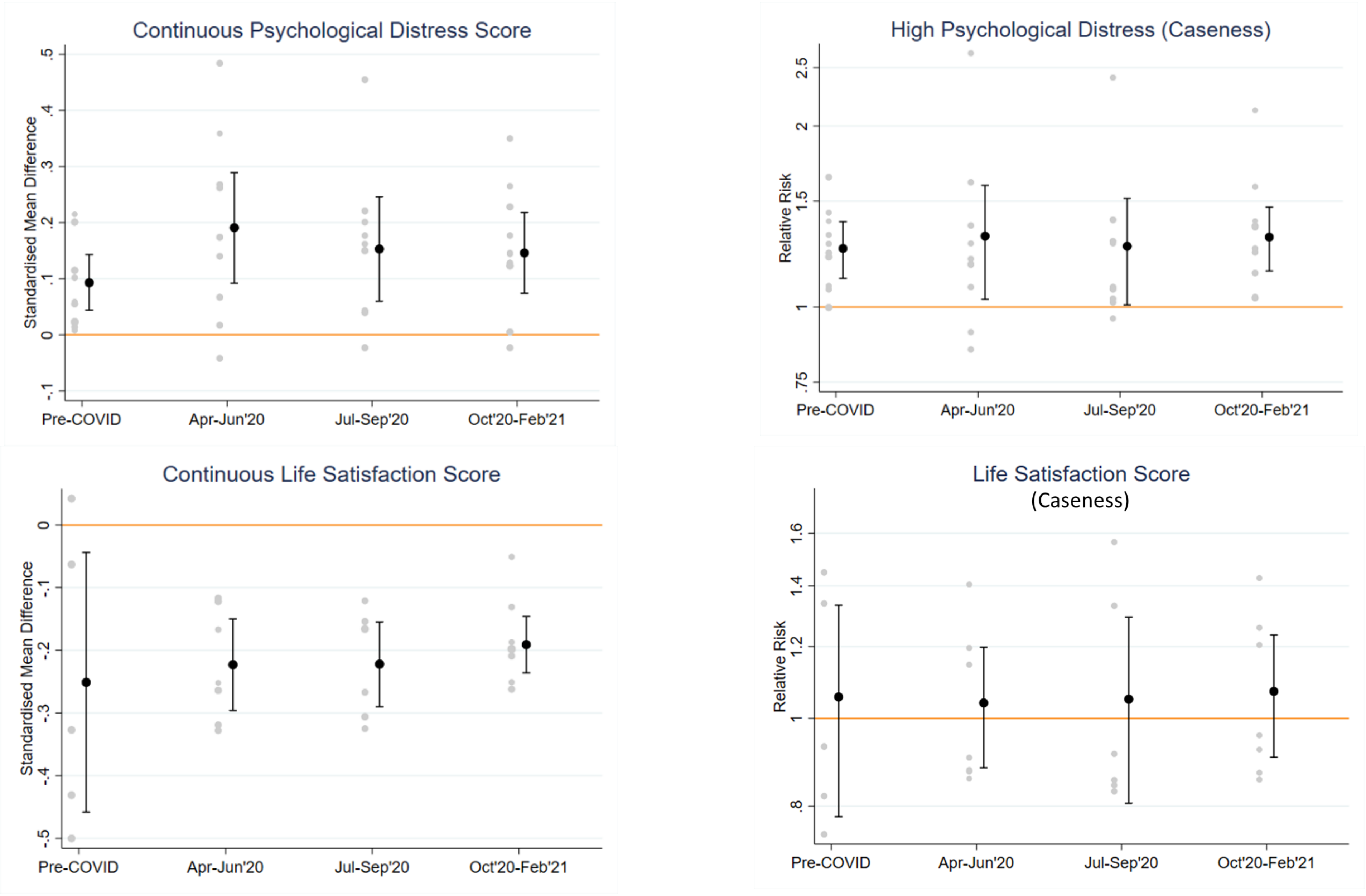
